# Rare Non-coding Variation Identified by Large Scale Whole Genome Sequencing Reveals Unexplained Heritability of Type 2 Diabetes

**DOI:** 10.1101/2020.11.13.20221812

**Authors:** Jennifer Wessel, Timothy D Majarian, Heather M Highland, Sridharan Raghavan, Mindy D Szeto, Natalie R Hasbani, Paul S de Vries, Jennifer A Brody, Chloé Sarnowski, Daniel DiCorpo, Xianyong Yin, Bertha Hidalgo, Xiuqing Guo, James Perry, Jeffrey R O’Connell, Samantha Lent, May E Montasser, Brian E Cade, Deepti Jain, Heming Wang, Peitao Wu, Silvia Bonàs-Guarch, Ricardo D’Oliveira Albanus, Aaron Leong, Irene Miguel-Escalada, Arushi Varshney, NHLBI Trans-Omics for Precision Medicine (TOPMed) Consortium, TOPMed Anthropometry, DIAMANTE, Gregory L Kinney, Lisa R Yanek, Leslie Lange, Marcio Almeida, Juan M Peralta, Stella Aslibekyan, Abigail S Baldridge, Alain G Bertoni, Lawrence F Bielak, Donald W Bowden, Chung-Shiuan Chen, Yii-Der Ida Chen, Seung Hoan Choi, Won Jung Choi, Dawood Darbar, James S Floyd, Barry I Freedman, Mark O Goodarzi, Ryan Irvin, Rita R Kalyani, Tanika Kelly, Seonwook Lee, Ching-Ti Liu, Douglas Loesch, JoAnn E Manson, Rami Nassir, Nicholette D Palmer, James S Pankow, Laura J Rasmussen-Torvik, Alexander P Reiner, Elizabeth Selvin, Aladdin H Shadyab, Jennifer A Smith, Daniel E Weeks, Lu-Chen Weng, Huichun Xu, Jie Yao, Zachary Yoneda, Wei Zhao, Jorge Ferrer, Anubha Mahajan, Mark I McCarthy, Stephen Parker, Alvaro Alonso, Donna K Arnett, John Blangero, Eric Boerwinkle, Michael H Cho, Adolfo Correa, L. Adrienne Cupples, Joanne E Curran, Ravindranath Duggirala, Patrick T Ellinor, Jiang He, Susan R Heckbert, Sharon LR Kardia, Ryan W Kim, Charles Kooperberg, Simin Liu, Steven A Lubitz, Rasika A Mathias, Stephen McGarvey, Braxton D Mitchell, Alanna C Morrison, Patricia A Peyser, Bruce M Psaty, Susan Redline, Dan Roden, M. Benjamin Shoemaker, Nicholas L Smith, Kent D Taylor, Ramachandran S Vasan, Karine A Viaud-Martinez, Jose C Florez, James G Wilson, Robert Sladek, Josée Dupuis, Stephen S Rich, Jerome I Rotter, James B Meigs, Alisa K Manning

## Abstract

Type 2 diabetes is increasing in all ancestry groups^1^. Part of its genetic basis may reside among the rare (minor allele frequency <0.1%) variants that make up the vast majority of human genetic variation^2^. We analyzed high-coverage (mean depth 38.2x) whole genome sequencing from 9,639 individuals with T2D and 34,994 controls in the NHLBI’s Trans-Omics for Precision Medicine (TOPMed) program^2^ to show that rare, non-coding variants that are poorly captured by genotyping arrays or imputation panels contribute h^2^=53% (P=4.2×10^−5^) to the genetic component of risk in the largest (European) ancestry subset. We coupled sequence variation with islet epigenomic signatures^3^ to annotate and group rare variants with respect to gene expression^4^, chromatin state^5^ and three-dimensional chromatin architecture^6^, and show that pancreatic islet regulatory elements contribute to T2D genetic risk (h^2^=8%, P=2.4×10^−3^). We used islet annotation to create a non-coding framework for rare variant aggregation testing. This approach identified five loci containing rare alleles in islet regulatory elements that suggest novel biological mechanisms readily linked to hypotheses about variant-to-function. Large scale whole genome sequence analysis reveals the substantial contribution of rare, non-coding variation to the genetic architecture of T2D and highlights the value of tissue-specific regulatory annotation for variant-to-function discovery.

---

Type 2 diabetes prevalence has exploded globally in all continental ancestry groups^1^. Large-scale genome-wide association studies (GWAS) have identified hundreds of T2D-associated genetic variants, most of which lie in the non-coding genome^7,8^. Twin studies in Europeans suggest genetic contributions to T2D risk (heritability) estimates of up to 72%^9^, but the discovered variants from GWAS in the same ancestry account for just 18% of T2D heritability^10^ some of which localizes to specific classes of pancreatic islet-specific enhancers^6^. “Missing heritability” may reside among the rare variants that make up the vast majority of human genetic variation and that are not interrogated by GWAS, despite initial suggestions from small samples of exome and low-pass whole genome sequencing^11-16^ that rare variants make a limited contribution to T2D heritability^13^. Here, we analyzed large-scale, multi-ancestry, high-coverage whole genome sequencing (WGS) from the NHLBI’s Trans-Omics for Precision Medicine (TOPMed) program^2^ and show that rare, non-coding variation contributes to T2D heritability. The non-coding genome provides no obvious framework for aggregating rare variants for association tests, so we used islet epigenomic signatures to annotate and group variants with respect to gene expression, chromatin state and three-dimensional chromatin architecture^4-6,17^. With this approach, we determined the global contribution of rare variation to T2D heritability, refined heritability estimates in the context of islet epigenomic signatures and used islet epigenomic annotation to frame genome-wide rare variant testing for novel discovery in the non-coding genome.

## Phenotypes and genotypes in the NHLBI TOPMed Program

The TOPMed program aims to understand genetic risk factors for complex cardio-metabolic diseases by combining WGS data with existing studies with deep phenotyping^2^. For this analysis, we included 44,633 individuals from 24 separate studies, representing a broad range of genetic diversity. For most analyses, we used genome-wide principal components (PCs) to assign individuals to one of five groups based on shared genetic background and used labels informed by the most common self-reported ancestry or population among each groups’ members: African, Asian, European, Hispanic/Latinx, and the Samoan Study (**Supplementary Table 1**). Individuals were given a single label to represent their particular genetic ancestry, but each group contained a diverse cross-section of race, culture, and admixture. Such labeling does not imply homogeneity, as ancestry does not exist on a discrete scale. There were 9,639 individuals with T2D and 34,994 controls (See **Methods** and **Supplementary Note** for disease definitions). The prevalence of T2D varied from 14-29% across the five groups (**Supplementary Table 2**).

Whole genome sequencing and joint genotype calling performed by TOPMed identified 373.3M variants in this sample that passed quality control (Freeze5b data release, average sequence depth 38.2x), which represents two to seven times more variants compared with recent GWAS^7,8,10^, exome-wide^15^, or WGS studies^13,14^ for T2D. In total, 92.8% of the variants were rare (minor allele frequency (MAF)<0.1%), 5.3% were low-frequency (MAF≥0.1% and <5%) and 1.8% were common (MAF≥5%; **Figure 1, Supplementary Figure 1, Supplementary Tables 3-5**). The vast majority of variants (95.4%, 360M) were located outside exons.

**Figure 1.**
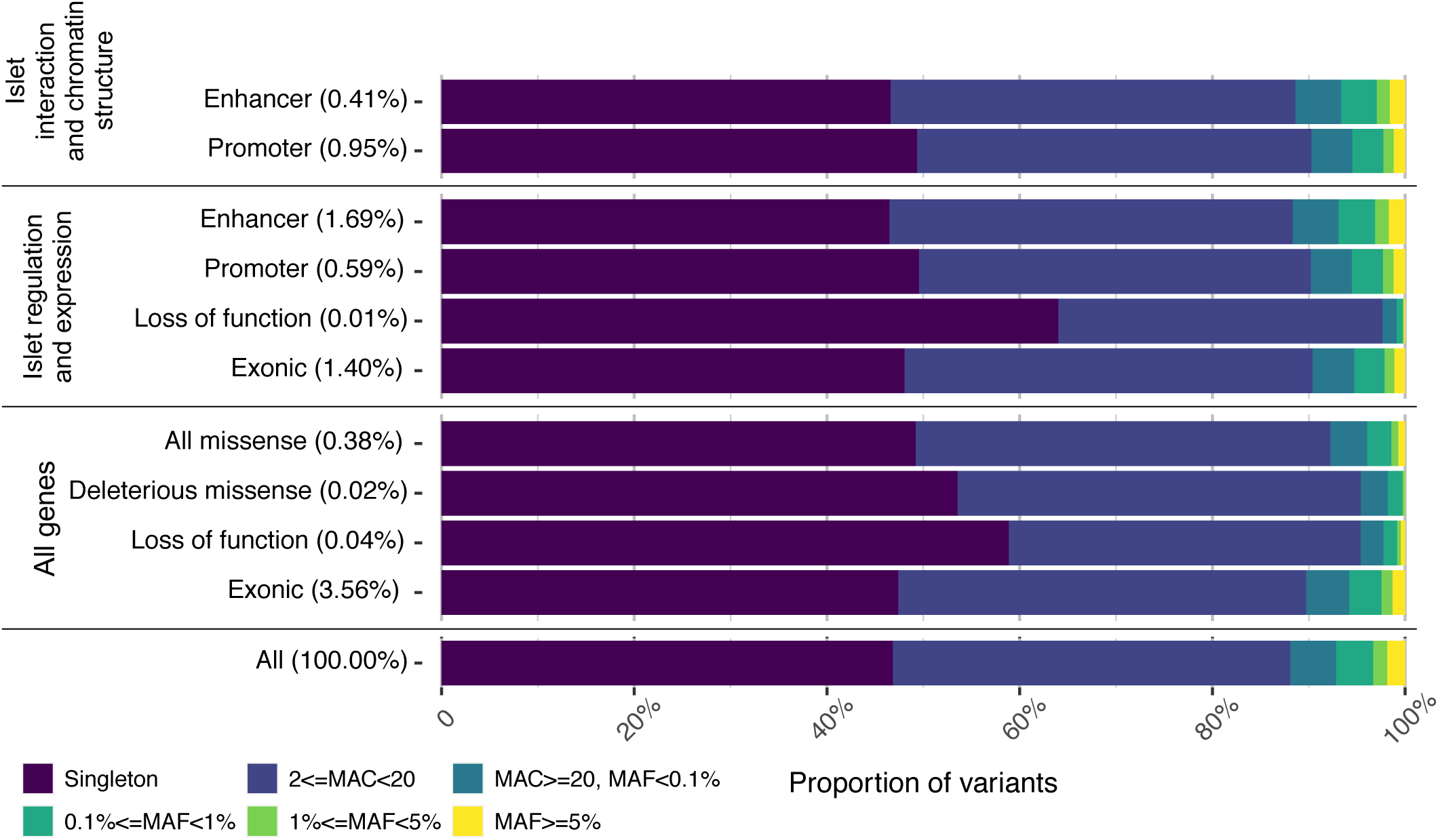
Allele frequency spectrum from whole genome sequencing. Variants were aggregated by minor allele count (MAC) or frequency (MAF) into six, non-overlapping ranges depicted by bar color. Pancreatic islet-specific, non-exonic, functional annotations and protein-coding annotations were used to further subdivide variant classes. Annotations were broadly grouped into three types with potentially overlapping variants. *‘Islet interaction and chromatin structure’* annotations relate to active regulatory regions involved in 3D chromatin interactions within islet cells. ‘*Islet regulation and expression’* annotations capture the regulatory regions of genes expressed in pancreatic islets. *All genes* annotations relate to variants falling within protein coding, exonic regions, partitioned by predicted effect on protein function. The variant frequency spectrum was dominated by singleton and extremely uncommon variants (darkest blue bars). Protein coding variants followed previously observed trends, with average frequency decreasing with increasing predicted severity on protein function (*All genes*), especially in *‘Islet regulation and expression’*. Annotations used in rare variant aggregation and association testing make up between 0.01% and 1.69% of total variation (‘*Islet regulation and expression’* and *‘Islet interaction and chromatin structure’*).

We identified 13.3M variants within exons of protein-coding genes, including 1.4M (0.38% of all variants) annotated as missense and 135K (0.04%) annotated as loss of function. A higher proportion of the loss of function variants were concentrated at the lower end of the frequency spectrum with singletons (58%) and variants with minor allele count (MAC) between 2 and 20 (37%) making up 95% of variants compared to all variants within the genome (46% and 41% for singleton variants and variants with MAC between 2 and 20, respectively, for a total of 87%). This is consistent with models of purifying selection on loss of function variants^18^. This difference was more pronounced for variants in the exons of genes expressed in pancreatic islets, where 98% of 35K loss of function variants had a MAC below 20 (64% singleton, 34% with MAC between 2 and 20).

## Islet-specific functional annotation of the non-coding genome

To identify potential variant function and to refine our search space for rare variants associated with T2D risk, we developed strategies to define functional units of the non-coding genome. T2D GWAS show a clear enrichment of common T2D risk variants mapping to genes expressed in pancreatic islets^19^ and to active promoter and enhancer regions in pancreatic islets^5,6,10,17^. We used two distinct strategies, referred to as ‘*Islet regulation and expression’* and ‘*Islet interaction and chromatin structure’*, to define regulatory regions in pancreatic islets on the basis of shared targets or coordinated function (see **Methods**). First, we used promoter and enhancer regions linked to islet-expressed genes to define the ‘*Islet regulation and expression’* annotation sets, creating groups of variants with possible coordinated regulatory function influencing the expression of a given gene^3,5,20^. Second, we defined variant sets based on the ‘*Islet interaction and chromatin structure’* annotations. These annotations describe large stretches of the genome that organize into 3-dimensional complexes, referred to as “hubs”^6^. These complexes form through physical interactions among enhancers and promoters, yielding loops of DNA tied together by regulatory elements that may contain one or more genes regulated by the “hub”.

In total, we identified 8.5M variants (2.28% of all variants) within promoters and enhancers using the ‘*Islet regulation and expression’* annotation set and 5.1M variants (1.36% of all variants) in the ‘*Islet interaction and chromatin structure*’ regions (**Figure 1, Supplementary Tables 3-5**). The overlap set of these two annotation strategies had 4.3M unique variants that were annotated as ‘promoter’ variants in either annotation set, with an overlap of 1.4M (32.7% of 4.3K) variants. Less overlap existed in the enhancer regions: 6.5M unique variants were annotated as ‘enhancer’ variants in either annotation set, with an overlap of 0.6M (9.2% of 6.5M) variants. No substantive difference in overlap was observed when variants were partitioned by frequency.

## T2D heritability from rare and common variants

We implemented a variant-based heritability analysis with the GCTA software to partition T2D risk into environmental and genetic components using the TOPMed WGS dataset. A major advantage of using WGS data to estimate variant-based heritability is that causal variants are directly ascertained in the sample. We applied multi-component heritability estimation to a subset of 15,109 unrelated individuals of European ancestry (2,215 with T2D; 12,894 controls), the largest ancestry subset, restricting to variants with a MAC greater than 5 and adjusting for BMI (**Supplementary Table 6**; See **Supplementary Methods** for more details). After observing differences in LD patterns across allele frequencies **(Supplementary Figure 2)**, we partitioned variants by MAF bins and LD score quartiles. We used these variants sets of 16 components (defined by four MAF bins and four LD score bins) to obtain heritability estimates which we considered statistically significant when P values were less than 0.05/16=0.003. In building our final models, we removed components with non-significant heritability estimates.

**Figure 2.**
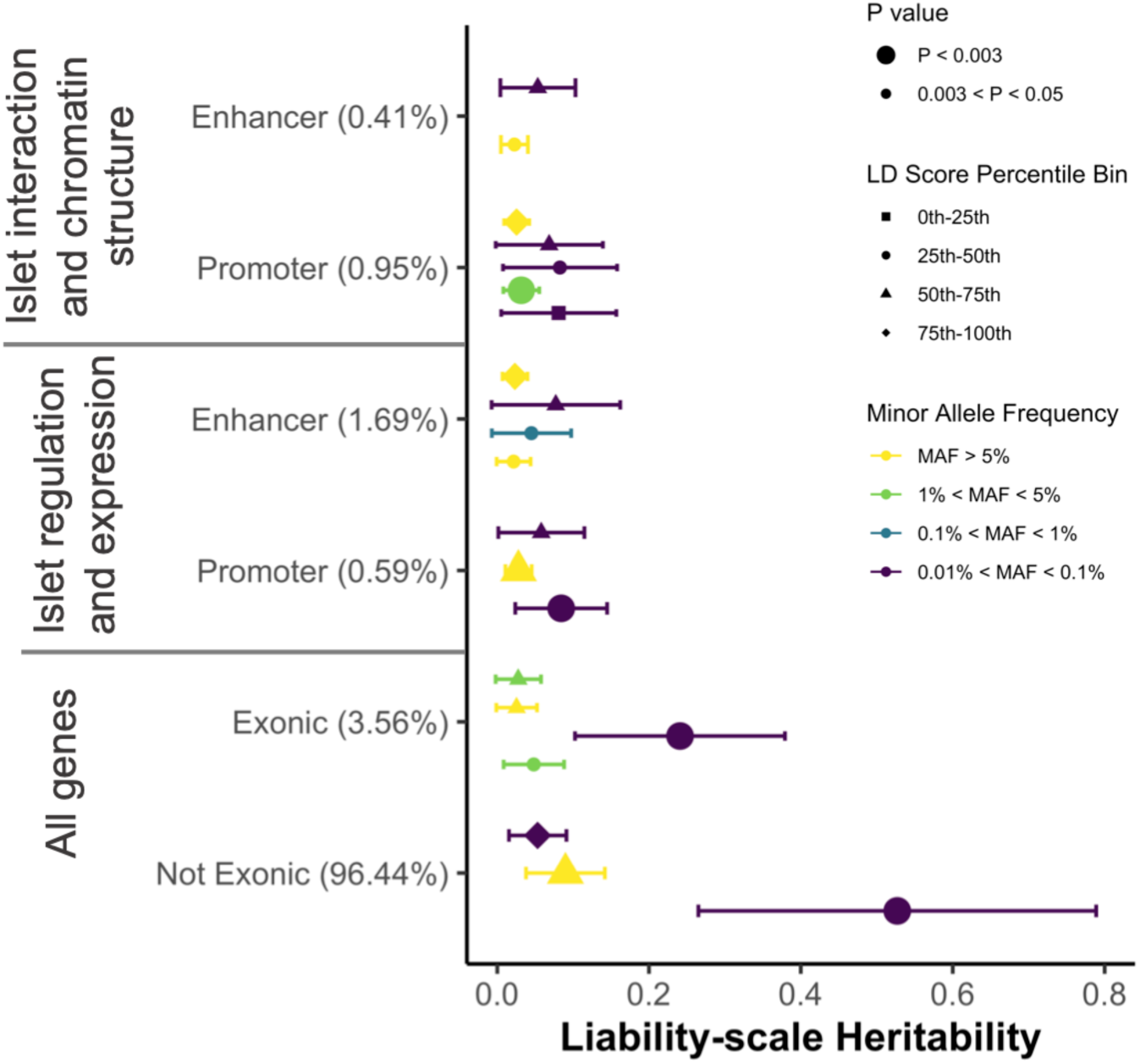
Variant-based heritability of type 2 diabetes, genome-wide and within islet-specific regulatory regions. Within each annotation, we quantified variant-based liability-scale heritability for T2D using genetic relationship matrices, with variants subdivided by variant frequency (colors), and LD Score quartiles (symbols), which measure the amount of variation tagged by a given variant (see **Methods**). The 95% confidence interval of each estimate is provided. Models were adjusted for age, sex, TOPMed project and BMI. We display the variant subsets with P value < 0.05 in our final models (**Supplementary Table 7**).

We compared T2D heritability estimates for the exons of protein-coding genes, into which 3.56% of all variants fall, to the ‘non-exonic’ portion of the genome (**Figure 2, Supplementary Table 7**). Estimated heritability of variants with moderate-to-low LD (in the 2^nd^ LD score quartile) were the largest. For rare, exonic variants estimated heritability was 24% (95% confidence interval [CI] 10%-38%, P=2.9×10^−4^) while for rare, non-exonic variants, the heritability estimate was 53% (95% CI 27%-79%, P=4.2×10^−5^). Interestingly, only the non-exonic estimate was changed upon BMI adjustment with a 9% decrease in estimated heritability from 59% (**Supplementary Table 7)**. For rare non-exonic variants in high LD (4^th^ LD score quartile) the heritability estimate was relatively small (5%, 95% CI 2%-9%, P=1.3×10^−5^). For common, non-exonic variants in the 3^rd^ LD score quartile (representing modest LD) heritability was 9% (95% CI 4%-14%, P=2.4×10^−4^), consistent with previous common variant GWAS^10^.

We further subset non-exonic variants using the previously described functional annotation sets to determine the degree to which these variants that fall into islet regulatory regions contribute to T2D risk. We observed significant heritability estimates in variant subsets (defined by MAF and LD score quartiles) that differed across annotations. In models with ‘*islet regulation and expression’* promoter variants, we observed a significant heritability estimate with rare variants with moderate-to-low LD (2^nd^ LD score quartile) of 8% (95% CI 2%-14%, P=2.4×10^−3^); and among common variants with moderate-to-high LD (3^rd^ LD quartile) heritability was 3% (95% CI 1%-5%, P=2.6×10^−4^). On the other hand, with ‘*islet regulation and expression’* enhancer variants, we observed a significant estimate with only one variant set: common variants with high LD (4^th^ LD score quartile) with an estimate of 2% (95% CI 1%-4%, P=6.6×10^−4^). With the ‘*islet interaction and chromatin structure*’ promoter variants, two variant sets contained a significant estimate: low-frequency variants with moderate-to-low LD (2^nd^ LD quartile) with a heritability estimate of 3% (95% CI 1%-6%, P=3×10^−3^) and common variants with high LD (4^th^ LD score quartile) with a heritability estimate of 3% (95% CI 1%-4%, P=3×10^−4^). The common, annotated variant heritability estimates are consistent with previous observations^6,21^. These results suggest that a substantial fraction of T2D risk is explained by the effects of rare variants with moderate-to-low LD and that promoter regions linked to islet gene regulation capture some, but not all, of this heritability.

## Rare variant association tests using islet annotation aggregation

Association tests that aggregate rare variants, like SKAT and Burden tests^22^, depend on gene bodies to frame variant aggregations. To frame variant aggregation in the non-coding genome, we used the two functional annotation sets to define three types of aggregation units for rare variant tests of associations with T2D: First, for each gene expressed in pancreatic islets, we created aggregation units based on the ‘*Islet regulation and expression’* annotation set using enhancers, promoters, and included predicted loss of function variants within the gene transcript. The second and third types of aggregation units, ‘*whole hub*’ and ‘*hub-components*’ used the ‘*Islet interaction and chromatin structure’* annotation set. *‘Whole hub*’ aggregation units included variants within all promoters and enhancers shown to interact to form each hub. ‘*Hub-components*’ were aggregation units for each individual promoter and enhancer component of each hub. In this strategy, each regulatory region was tested individually, outside of its 3-dimensional context. We also generated three commonly used coding variant aggregation strategies (loss of function, deleterious missense, and all missense) to complement the non-coding islet-specific aggregation strategies.

For each aggregation strategy, we used mixed models that accounted for family relationships and population structure^23-25^ to test groups of variants for association with T2D using SKAT and Burden tests^22^. Association analyses were performed within each ancestry group or study population; since most rare alleles included in aggregation units were observed in only a subset of the ancestry groups or study population (see **Methods** for statistical significance thresholds for each analysis). Although our primary model used BMI adjustment, several loci showed significant associations without BMI adjustment. Such observations guide our biological interpretation of these loci and suggest mechanisms related to obesity and insulin resistance.

Eight ancestry- or study population-specific rare variant aggregation units were significantly associated with T2D (**Table 1, Supplementary Table 8, Supplementary Figure 3)**. We defined ‘driver variants’ as the set of variants contributing substantially to the aggregation unit test statistic (see **Methods**). The significant rare variant aggregation units had between one and four variants driving the observed associations, representing 11 total variants, none of which were associated with T2D in single variant analyses (**Table 2, Supplementary Tables 9-10**). No variant contributed 100% to the test statistic, though, and for all our tests, the aggregate test P value was lower than the P values of individual variants (when allele count was high enough for a single variant test; **Supplementary Table 9**). This demonstrates that these observations were only possible through multi-variant testing, pointing to the critical value of tissue-specific annotation to frame aggregation tests in the non-coding space. The two missense variants driving the chromosome 10 associations linked to *FO681492*.1, a poorly characterized reverse strand transcript, were not present in previous genome builds and were not reported in previous whole exome sequence association studies of T2D^13,15^. Five associations were from the ‘*hub-components’* aggregation units, representing four loci on chromosomes 2, 3 and 15. At the chromosome 2 locus spanning *NR4A2* and *GPD2*, four variants were identified: rs530551407, rs200945165, rs200622604, and rs559881272. Of these, rs200945165 contributed most to the rare variant test statistic (19.24%), was predicted to be a non-coding deleterious variant, and had the highest CADD score. All four of these driver variants fall within an active transcriptional start site, identified in pancreatic islet tissue, suggesting a regulatory mechanism through which the T2D association occurs. On chromosome 15, three aggregate associations were driven by the same three variants which contribute nearly equally to the test statistics: rs145197571, rs79569357, and rs28427880. These included two associations from the ‘*islet regulation and expression*’ aggregation units which contained the same enhancers but were linked to two different genes and overlapped a ‘*whole hub’* signal linked to *MRPL46, MRPS11* and other genes. Each variant was identified in active transcriptional start site in islets, again suggesting a common regulatory mechanism.

**Table 1:**
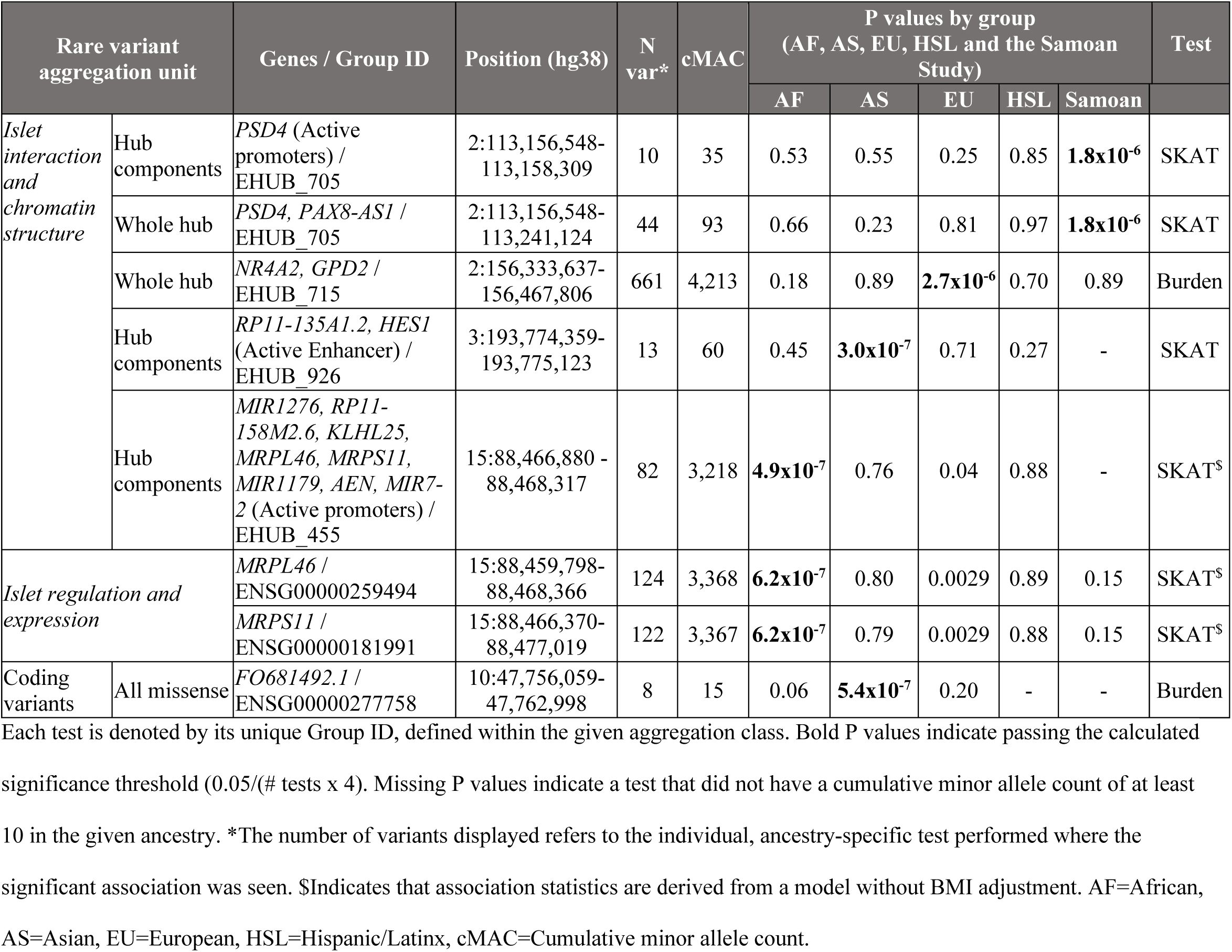
Rare variant associations with type 2 diabetes in 44,633 TOPMed multi-ancestry whole genome sequences.

**Table 2:**
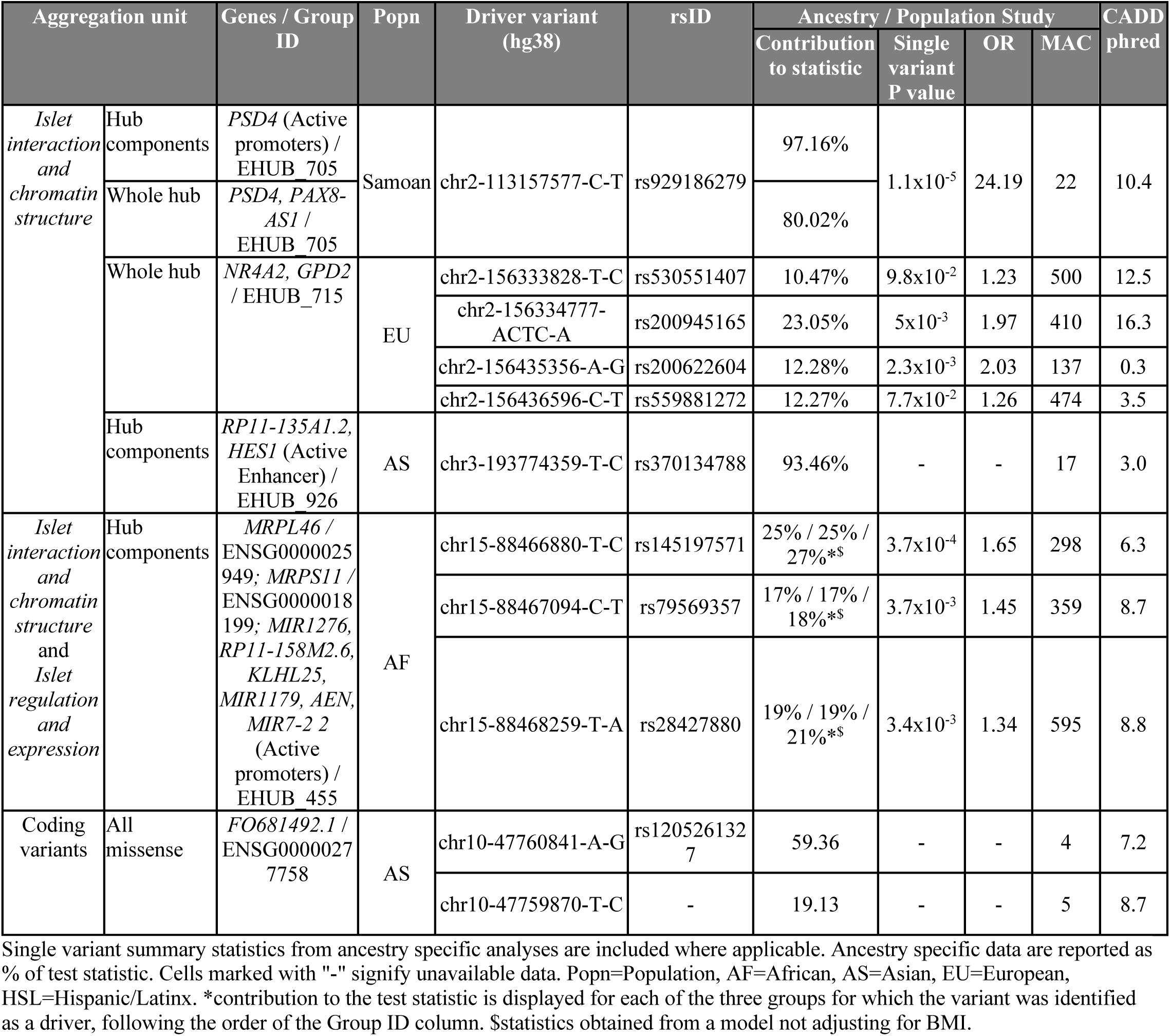
Rare variants driving associations with type 2 diabetes in 44,633 TOPMed multi-ancestry whole genome sequences.

We also tested the coding genome to understand if islet expressed genes were enriched for rare variant associations with T2D but only observed enrichment in one set of rare variant tests: loss of function aggregation tests within the Hispanic/Latinx ancestry group (P = 0.0054; see **Supplementary Methods** and **Supplementary Results**; **Supplementary Table 11**).

## Common variant association tests

We also conducted single variant association analyses with variants having a minor allele count greater than 20 in our TOPMed WGS data. We used mixed models that accounted for family relationships and population structure^23-25^ to test individual common variants for association with T2D in ancestry-specific and pooled (i.e. entire sample) analyses. We identified seven variants at six loci at WGS-wide level of significance (P<4×10^−9^, Bonferroni corrected for the effective number of independent regions from WGS across chromosomes 1-23, see **Methods**) or locus-wide significance (P<1×10^−5^; **Supplementary Table 12, Supplementary Figure 4**). These loci (labelled by their nearest gene: *CDKN2B*-*AS1, TCF7L2, KCNQ1, CCND2, FTO* and *DUSP9*) have been previously reported^10^. We did identify a novel secondary signal at the *CDKN2B*-*AS1* locus: rs150046492 (MAF=0.01, odds ratio [OR] = 0.67, conditional P=8.2×10^−6^; **Supplementary Figure 5**). We explored different modeling strategies by collating genome-wide significant associations (where P values are greater than 5×10^−8^ but less than our WGS significance threshold), examining associations with related cardiometabolic traits (see **Supplementary Note**) and refining the definition of controls with high glycemia and designating them as cases (referred to as T2D+). By lowering the significance threshold to GWAS level (P value > 5×10^−8^) we identified 2 known loci (*ADCY5* and *SLC30A8*) and 6 possibly novel loci (*ODF2L, LMAN2, NKX2-5, KCNV1, VLDLR-AS1 and LINC01052*) where alleles are low-frequency, rare (MAF<2%) or ancestry-specific (**Supplementary Table 13, Supplementary Note**). We found an additional known locus (*MTNR1B*) and a potentially novel locus (*NWD2*) associated with the T2D+ outcome (**Supplementary Table 14, Supplementary Note**).

We generated 95% credible sets to refine the likely causal variants in these regions, and as recently reported^7^ found that the inclusion of diverse ancestries improves upon previously reported credible sets for T2D. At five loci, the 95% posterior probability for a single variant exceeded 0.9 (**Supplementary Table 15**). For *TCF7L2*, the European analysis credible set consists of the same three variants as seen in the DIAMANTE European GWAS^10^; however, our pooled and African American credible sets contain only one variant, rs7903146, which had the highest posterior probability in the European credible set (0.46)^10^ and has been characterized as the causal variant at the locus^26^. Comparing our credible sets with credible sets reported in the DIAMANTE European GWAS, we observe consistency in the variants within the sets and the variant with the highest posterior probability is often the same in both sets (**Supplementary Figure 6, Supplementary Table 15)**. The most notable differences between credible sets occurs where neither set has high posterior probability for any variant (e.g. *SLC30A8* and *CDKN2B-AS1*).

Power to detect single variant associations in our current study is modest (80% power to detect ORs of 1.25 and 3.5 for variants with MAF of 5% and 0.1%, respectively, and 50% power to detect an OR of 5.5 for variants with MAF of 0.02%; **Supplementary Figure 7**); particularly in comparison to recent GWAS^10^. Therefore, we examined the results of 380 of the 403 variants from the DIAMANTE consortium analysis of European ancestry^10^ with imputed GWAS of T2D as a metric of utilizing currently available WGS that provides 14-fold (373M vs. 27M) more variants but has 20-fold smaller sample size (44,633 vs. 898,130). We found 113 (30%) of the index variants were nominally significant in our WGS sample (P<0.05) and at 7 of these loci, the previously reported index variant was the most significant variant at the locus (*TCF7L2, KCNQ1, ADCY5, CCND2, ATP1B2, JAZF1, BCL2A*). We examined associations of other variants around the 239 loci containing the 380 index variants, and we found for 251 (66%) variants, there was at least one variant in the region with a lower P value than the index variant (**Supplementary Table 16**). Notably, 184 (73%) of these “smaller P value” variants had a MAF < 1%. However, when we examined QQ plots of variants in these regions stratified by allele frequency, we did not observe an inflation in P values in the set of variants with MAF < 1% (**Supplementary Figure 8**).

## Novel variants and whether they were missed by imputation

We next asked whether newly observed associations identified in this study (by single variant, conditional, or aggregation analyses) would have been seen in previous common or rare variant studies of T2D. We examined the association and imputation quality of our variants in the previous GWAS performed by the DIAMANTE consortium^10^ which used the human reference consortium (HRC) imputation reference panel in a European-ancestry meta-analysis of 74,124 T2D cases and 824,006 controls (**Supplementary Table 17**). Of the twenty-eight variants we identified through single variant or conditional analyses (**Supplementary Table 17a**), fourteen were not identified as significant in the previous GWAS and represent potentially novel loci. Seven of these fourteen variants were not included in the GWAS because they were not part of the HRC reference panel. The remaining seven novel variants were previously analyzed but were not significant at P<5×10^−8^. For these variants, imputation quality was generally high (>0.60) but were identified here through different analytical methods or different ancestry groups than those utilized in the DIAMANTE GWAS. Eleven variants were classified as ‘driver’ variants in our aggregation analyses (**Supplementary Table 17b**) and eight of these variants were not included in the GWAS because they were not part of the HRC reference panel. The remaining three variants had high imputation quality and the P values from our single variant analysis were similar to those in the GWAS. Taken together this suggests WGS data generated on diverse populations will uncover additional variants associated with T2D susceptibility.

## The contribution of high-coverage WGS to the genetic architecture of T2D

We characterized the contribution of genome sequence to the genetic architecture of T2D by cross-tabulating allele frequency, annotation, and ORs for association among variants that achieve a sub-significant association (P<5×10^−5^; “subthreshold variants”), revealing a few qualitative patterns in our data (**Supplementary Table 18**): an enrichment of subthreshold association signals in the non-coding functional annotations, indicating that additional islet regulatory signals could be found as our sample size increases; and a difference in the magnitude of ORs in these annotations compared to exonic annotations, indicating a genetic architecture for these genomic regions that are potentially unique.

First, we used a simple hypergeometric test to determine if the number of subthreshold variants was enriched across the annotation categories and found an enrichment in the ‘*islet interaction and chromatin structure*’ annotation class (31 variants; Enrichment P=7.1×10^−6^), which persisted when examining variants with allele frequency less than 0.1% (10 variants; Enrichment P=0.002). This test ignores pairwise LD, which does exist in the set of subthreshold variants, mostly among the common variants. We accounted for LD by clumping the variants with r^2^>0.2 and observed that the 22 independent signals represented by the 31 variants still show an enrichment signal (P=0.008), indicating that additional T2D risk variation may yet be found in the ‘*islet interaction and chromatin structure*’ regions of the genome as we increase our sample sizes.

We expected that as variant allele frequency decreased, effect size would increase. We compared the ORs for association with T2D relative to allele frequency and annotation among variants with P<5×10^−5^ (**Supplementary Figure 9, Supplementary Table 19)**. We observed that coding variants, including missense variants and missense variants in genes expressed in islets, and non-coding variants classified as *‘islet regulation and expression’* or ‘*islet interaction and chromatin structure*,’ were represented across the distribution of observed minor allele frequencies and ORs. Among all coding variants tested, the mean OR of ‘islet-expressed missense variants’ was higher compared to ‘all missense variants’ or ‘all coding variants’ (mean OR±SD = 6.1±8.8 vs 3.5±5.1 vs 3.7±6.8, respectively). However, the mean OR was much higher in rare variants (allele frequency <0.1%); and in ‘islet-expressed missense variants’ compared to ‘all missense variants’ or ‘all coding variants’ (14.3±13.6 vs 12.8±11.5 vs 9.3±10.9, respectively). In contrast, variants in the *‘islet regulation and expression’* or *‘islet interaction and chromatin structure’* annotation sets had a similar mean OR for association with T2D (3.0±3.6 or 3.6±5.5, respectively) compared to all variants with P<5×10^−5^ (3.7±6.8). The attenuated effects from the pancreatic islet regulatory annotations compared to the exonic variants suggest that these variants will require larger sample sizes to have statistically significant effect estimates compared to the exonic variants. Finally, we did not observe any rare variants with large protective effects, and in general, there were fewer T2D protective than T2D risk alleles across the observed allele frequency spectrum.

## Discussion

WGS interrogates the >97% of the genome that is non-coding, dramatically increasing the number of disease or health-related variant associations discoverable in individuals and populations. TOPMed high coverage, jointly called WGS data offers many millions of rare, common and population-specific variants that allow us to redefine the genetic architecture of T2D; even in a sample size smaller than current T2D GWAS. In this large-scale multi-ancestry association study within TOPMed, we observed common variant heritability estimates that were consistent with common variant GWAS^10^, but identified a remarkably larger contribution of rare, non-coding variation to T2D heritability estimates. These data revise prior T2D heritability estimates that used just a few thousand low-pass sequences to model heritability and concluded that rare variation contributes relatively little to T2D heritability^13,14,16^. Although the large amount of rare variant heritability for T2D seems outsized, this has now been seen in TOPMed WGS for height and BMI^27^, and may be a genetic architecture characteristic of polygenic traits and phenotypes in general. Notably, this class of variants would not be captured by prior array-based genetic studies nor be imputable with current reference panels, consistent with similar evidence from exome sequencing studies of the additional value of sequencing over array-based imputation^15^. We now identify T2D to be a genetic disorder whose architecture spans the spectrum from common to rare alleles, and from polygenic to monogenic, with rare alleles contributing far more than previously thought to the heritability of the common polygenic form of T2D.

Islet-specific annotation provided frames that refined our search space for rare variant tests. By integrating annotation from islet regulatory function, we found a higher proportion of loss-of-function alleles in the sets of genes expressed in islets compared to all genes. Tissue-specific annotation data also permit a framework to aggregate rare variants into functional elements and variants sets for burden testing WGS-wide. Despite the lower power of our analysis versus current T2D GWAS, our approach identified five ancestry-specific rare variant association signals for further follow-up. Our revised heritability estimates suggest there are many more T2D pathobiology variants to be found in the rare-allele, non-coding genome. As TOPMed and annotation resources grow^3^ these approaches will be valuable to identify T2D causal variants in a framework that points directly to specific functional hypotheses for mechanistic follow up of T2D pathobiology.

## Supporting information

Funding

Parent Study Ethics Statements

## Data Availability

Data will be available on the Accelerating Medicines Partnership Type 2 Diabetes Knowledge Portal website.

http://t2d.hugeamp.org/

## METHODS

### Genome Sequencing

The National Heart, Lung and Blood Institute’s TOPMed (nhlbiwgs.org) Freeze5b data were used in these analyses. The samples were sequenced at an average depth of >30x coverage at the Baylor College of Medicine, the Broad Institute, Illumina, Macrogen, the New York Genome Center, and the University of Washington^2^. All samples from a given study were sequenced at the same center. Sequencing reads were aligned to human genome build GRCh38. Quality control was performed at each stage of the process and is described in detail (https://www.ncbi.nlm.nih.gov/projects/gap/cgi-bin/GetPdf.cgi?id=phd007493.1). Individual genetic variations across the genome were identified in a joint variant calling framework, utilizing all samples collected and conducted by the TOPMed Informatics Resource Center (University of Michigan), which also performed centralized read mapping and genotype calling. These analyses produced variant quality metrics which were used in variant filtering and reporting of samples that failed to meet quality control (QC) thresholds. Data management and QC to ensure correct sample identification, and general study coordination were provided by the TOPMed Data Coordinating Center (University of Washington). For duplicate individuals that participated in multiple studies (e.g. both ARIC and JHS), the TOPMed Diabetes Working Group developed an algorithm to retain only a single set of genotypes and phenotypes for each duplicated individual based on sequencing quality, type of study, and availability of phenotype data (see **Supplementary Note**). For all T2D associated variants, genotyping quality was further checked by inspecting plots of sequencing depth and genotype quality by carrier distribution; and examining alignment of raw sequences on BRAVO (https://bravo.sph.umich.edu/freeze5/hg38/). All variants reported passed this final visual examination.

### Cloud Computing Platforms

Analyses of WGS data were carried out on cloud platforms^28^: Analysis Commons, ENCORE and Terra. These platforms provide data sharing mechanisms that allows for the pooling of both genotypic and phenotypic data across multiple studies. This enables the kinds of analyses that WGS data requires, due to the size of the WGS data and pooling of phenotypic data for rare variant analyses.

### Trait Harmonization

A T2D trait and phenotype harmonization protocol was developed that defined T2D status and covariates needed for analyses. This protocol was shared with studies willing to harmonize their data and study investigators created a file utilizing their in-depth understanding of the study; for other studies, we downloaded data and data dictionaries from the database of Genotypes and Phenotypes (dbGaP) to create a standardized, harmonized study level dataset (see **Supplementary Table 1**). Study level participant characteristics are provided in **Supplementary Table 2**. Studies that provided measures of fasting glucose (FG) and/or hemoglobin A1C (HbA1c) were also used to define an additional T2D phenotype (referred to as T2D+ outcome, see **Supplementary Note**). Previous work has demonstrated that individuals whose FG (≥6.1mmol/L) and HbA1c (≥6.0%) levels are in the ‘pre-diabetic’ range have a 68% absolute risk of being diagnosed with T2D in the next 20 years^21^. We used these definitions to refine our control group and to identify individuals who are likely to develop T2D. Analyses with T2D+ are described in the **Supplementary Note**.

### Ancestry Definition

Genome-wide principal components (PCs) were derived from common genetic variants using PC-Air^29^ and made available to the TOPMed consortium (https://www.nhlbiwgs.org/topmed-whole-genome-sequencing-project-freeze-5b-phases-1-and-2). We used Ward’s hierarchical agglomerative clustering method (R hclust, Ward.D2 method) using the PCs scaled to their Eigen values to derive genetic ancestry group assignments for each individual, resulting in five groups. Each group was labeled by the most common self-reported ancestry or population among its members: African, Asian, European and Hispanic/Latinx ancestry, and one population-based study, the Samoan Study (**Supplementary Table 2**). Fourteen individuals whose self-reported ancestry or population was not represented by the five groups were excluded from analysis. An additional 136 individuals from 12 TOPMed projects were excluded because fewer than 5 individuals from their respective projects were assigned to a genetic ancestry group.

### Variant annotation

We partitioned genetic variants across the genome into ten, non-exclusive classes based on general and T2D-related, functional annotations (see below). We also partitioned the genome into six frequency-based classes: (1) singletons, (2) doubletons to minor allele count (MAC) < 20, (3) MAC ≥ 20 and minor allele frequency (MAF) < 0.1%, (4) 0.1% ≤ MAF < 1%, (5) 1% ≤ MAF < 5%, and (6) MAF≥ 5%. The ten annotation-based classes and the six frequency-based classes were using to define 60 possibly overlapping variant classes. Variant annotation and partitioning were performed using the Hail software^30^, a suite of highly parallelizable computational methods leveraging distributed cloud computing resources.

### Functional annotation of the non-coding genome

Multiple sources of functional, non-coding genomic annotations were used to characterize genetic variants. Annotations were derived from assays of pancreatic islet tissue, including gene expression^4^, chromatin accessibility^4^, and 3D contact maps^6^. Annotations were grouped into two distinct sets: ‘*Islet regulation and expression’* and ‘*Islet interaction and chromatin structure*.’ We used the Whole Genome Sequence Annotator (WGSA) to annotate SNPs and indels^31^.

#### Islet regulation and expression

We determined the gene transcripts expressed in pancreatic islets from RNA-seq data of 89 individual donors^3,4^ made available in the Diabetes Epigenome Atlas^3^. We restricted to transcripts where the average (FPKM) was greater than 2 among all samples. Non-exonic regions were annotated by islet chromatin state maps^5^ with particular focus on regions denoted as active enhancers and active transcriptional start sites (active promoters), both of which have been implicated previously in T2D predisposition^26^. Active promoter regions were assigned to islet-expressed genes by distance, creating a promoter-gene link if a gene’s first exon was within 5KB of the promoter. Promoters with no islet-specific link were removed. Enhancer regions were linked to genes using the GeneHancer database^20,32^, a tissue-agnostic enhancer-gene map^20^. We retained the islet-specific enhancers which overlapped the GeneHancer enhancer regions with at least 1 base pair overlap. The remaining regions were then filtered by gene target to retain those enhancers linked to islet-expressed genes.

#### Islet interaction and chromatin structure

A separate set of annotations^6^ was also used to characterize non-exonic function, termed the *Islet interaction and chromatin structure*. Here, complexes were defined through multiple sequencing-based methods designed to interrogate chromatin-chromatin contact, chromatin accessibility, and protein-DNA binding. We obtained available data for contact maps that were generated using promoter-capture Hi-C^33^ and accessibility and protein binding that were assayed through ATAC-seq^34^ and ChIP-seq^35^. These annotations are comprised of two parts (1) predicted regulatory function within a small region and (2) long-range, 3-dimentional (3D) interactions. Similar to chromatin states, observed pancreatic islet ATAC-seq peaks were classified into discrete categories based on histone modification, of which we used regions annotated as active promoters or active enhancers (class 1). The long-range, 3D interaction annotations were used to define “hubs” and predicted gene targets of the active promoters and enhancers within these ‘hubs’. These identified both regions with large 3D structures built from looping and interacting chromatin and the enhancers and promoters within these physically interacting regions.

### Exonic annotations

We used common bioinformatics tools^31,36-40^ to determine coding variant impact on protein function and create coding variant aggregation units. Within the exons of protein-coding genes, we focused on three classes of variation with differing predicted impact on protein function: ‘loss of function,’ ‘deleterious missense,’ and ‘all missense.’ The Variant Effect Predictor (VEP) was used to determine the functional consequence of a given variant, categorized broadly as high, moderate, or low impact^36^. ‘Loss of function’ variants were defined by high-impact variant effect predictions including the annotations: frame-shift, stop gain, stop loss, start loss, and transcript ablation^36^. Five common bioinformatics tools were used to define ‘deleterious missense’ variants, predicted to have non-tolerated impact on protein function by all methods: LRT^37^, Mutation Taster^38^, PolyPhen2-HumDiv^39^, PolyPhen2-HumVar^39^, and SIFT^40^. Finally, ‘all missense’ variants include those variants annotated as “missense” per VEP^36^. We also considered a subset of coding variant aggregation units from genes expressed within pancreatic islet cells^3,4^.

### Heritability estimation

Heritability refers to the proportion of disease risk conferred by genetics. To understand the contribution of variant subsets defined by predicted function to the heritability of T2D risk, we estimated variant-based heritability (h^2^) for all exonic variants, all non-exonic variants, variants annotated as promoters or enhancers from the ‘*Islet regulation and expression’* annotations, and variants annotated as promoters or enhancers from the ‘*Islet interaction and chromatin structure’* annotations. We used plink v1.9 and GCTA v1.91.7-1.93.0 and followed the procedure for estimating SNP-based heritability in imputed or whole genome sequence data described in Yang *et al*.^41^ and Evans *et al*.^42^ (See **Supplementary Methods** for more details). We used an unrelated subset of European-ancestry individuals from eight TOPMed projects (N=15,109): AFGen, CFS, COPD, FHS, GeneSTAR, GOLDN, MESA, VTE. We partitioned variants by minor allele frequency into 4 classes: 0.01% < MAF < 0.1%, 0.1% < MAF < 1%, 1% < MAF < 5%, and 5% < MAF < 50%. We calculated the LD Score^43^, a measure describing the amount of variation tagged by a given variant, for each variant, and further partitioned variants within each MAF class using the 25th, 50th and 75th percentiles of the LD Score distribution, resulting in 16 sets of variants from which we constructed genetic relationship matrices (GRM) for h^2^ estimation. These variant sets were then restricted to each annotation to estimate the liability scale h^2^ for T2D (with population prevalence = 0.08) using variance components models adjusting for age, sex, and TOPMed project. We adjusted for BMI in an additional model. In each analysis, we first estimated h^2^ with each GRM alone. Then, we jointly tested only the GRMs with h^2^ < 0.05 or variance > 0.001 from the single-GRM model; and report the liability-scale h^2^ estimate, h^2^ variance and P value of each GRM from the joint model. Of note, we have used the method implemented in the GCTA software to estimate the proportion of variation in disease liability^44^ to provide heritability estimates that are more interpretable compared to the 0-1 scale used to code binary disease outcomes. By definition, liability of disease is assumed to be the sum of environmental and additive genetic components from independent normal distributions. As described in Lee *et al*.^44^, there are several advantages to working under the liability scale, mainly that heritability is independent of prevalence. There is some concern that liability scale estimates could be biased by population substructure^45^ and we performed sensitivity analyses that used genetic principle components as covariates.

### Rare variant aggregation and association analyses

Rare and low-frequency genetic variants were grouped into aggregation units with six aggregation strategies that we used in rare-variant test for T2D associations. Our aggregation strategy assigns variants to groups based on gene targets or higher order 3-dimensional chromatin complexes linked to the co-regulation of sets of genes. Three strategies predominantly focused on non-coding variation while the remaining three focus on coding variation.

#### Islet regulation and expression

The ‘*Islet regulation and expression’* annotation set was used to generate aggregation units with islet-expressed genes as the basic organizing unit. Variants with MAF ≤ 1% were mapped to islet-expressed genes by the promoter and enhancer chromatin state predictions described above. If present, loss of function variants falling within an islet expressed exon of each gene were also included in the aggregation unit.

#### Islet interaction and chromatin structure

(*1*) *Hub-components:* Variants with MAF ≤ 1% within individual regulatory regions, enhancers and promoters, from the ‘*Islet interaction and chromatin structure’* annotation set were tested for cumulative association with T2D. Each regulatory interval served as the organizing unit for this aggregation strategy. (*2*) *Whole hub:* We used the same genomic regions and variants as in the ‘*hub-components’* aggregation units to create aggregation units that correspond to individual enhancer hubs shown to spatially interact to form 3D chromatin complexes. Each group, organized by physical interaction from islet promoter-capture Hi-C, consisted of multiple promoters and enhancers. These aggregation units contain the exact same genetic variants as the hub-components, gathered into larger groups of interacting promoter and enhancer regions.

#### Coding aggregation units

A gene-centric approach was taken for exonic variant aggregation and association testing. We followed similar variant annotation class definitions as defined by Fuchsberger et al^13^, creating three annotation classes: loss of function, deleterious missense, and all missense, as described above in the **Exonic annotations** subsection. While the loss of function annotation set includes variants across the frequency spectrum, both deleterious missense and all missense aggregations limit variants to MAF < 1%.

### Aggregate association analyses

SNP-set associations were evaluated using both SKAT and Burden tests per the GMMAT method^22^ (implemented in the GENESIS R/Bioconductor package) assuming an additive genetic model. GMMAT fits a logistic mixed model and performs Score (or Wald for effect estimates) tests under the null hypothesis of no association between a binary trait and each genetic variant to control for population structure and relatedness within the sample. Analysis of each ancestry group or study population was performed separately, using the same set of covariates (age, sex, TOPMed project), adjusted for population structure using a genetic relatedness matrix^25^. Meta-analysis was not performed since many of the rare alleles included in an aggregation unit were only observed in a subset of the ancestry groups or study population. Four analyses were performed for each aggregation strategy and ancestry group or study population using two statistical tests (SKAT, Burden) and BMI adjustments (BMI adjusted, BMI unadjusted). In all cases, association results were filtered to those aggregation units with a cumulative minor allele count greater than 10. The workflows utilized in this analysis are available in a public github repository (version 1.4.1; https://dockstore.org/workflows/github.com/AnalysisCommons/genesis_wdl/genesis_GWAS:v1_4_1?tab=info) from the Analysis Commons consortium^28^.

Aggregation units that passed a Bonferroni corrected significance threshold (between 2.97×10^−5^ and 1.94×10^−6^ accounting for the number of tests and the four analyses performed within each aggregation strategy and ancestry group/study population pair; **Supplementary Table 19**) were further explored to understand which variants were driving the observed association. Both SKAT and Burden analyses rely on generating a per-group of SNP’s test statistic through an additive model of individual variant score statistics, allowing for back-calculation of the contribution of each variant to the observed association. Starting with the variant with the highest contribution to the test statistic and proceeding in order of decreasing contribution, the driver variants for a particular aggregation unit was defined as the minimal set of variants with a combined contribution of at least 50%. Any remaining variants with a contribution greater than 10% were also considered ‘driver variants’.

### Single Variant Analysis and Functional Fine-Mapping Association testing

We performed single variant association tests with T2D using the Scalable and Accurate Implementation of Generalized mixed model (SAIGE) method assuming an additive genetic model. Our single variant association analyses were performed on variants with minor allele count > 20. SAIGE implements an accurate generalized mixed model association test that computes accurate P values in the presence of extreme case-control imbalance^23^. An empirical kinship matrix was used in SAIGE to control for population structure and relatedness within the sample. Both pooled analyses (i.e. entire sample) and ancestry/population-specific analyses were performed. In a meta-analysis across ancestries/populations, heterogeneity was assessed with Cochran’s Q statistic. Covariates used in ancestry-specific analyses included age, sex, TOPMed project (where TOPMed project represents either a subset of individuals from a single study selected based on certain criteria identified the study’s investigators or a consortium of studies that each contribute to a particular phenotype of interest); and covariates used in pooled analyses included age, sex, ancestry/population group, TOPMed project and the first 7 PCs. We also evaluated BMI as a confounder by performing analyses with and without BMI as a covariate. Statistical significance thresholds for single variant associations were determined by assessing the effective number of independent regions across chromosomes 1-23^46,47^ in the entire study sample and then performing a Bonferroni correction (**Supplementary Table 20**), which resulted in a threshold of 4×10^−9^.

### Fine-mapping of potential causal variants within T2D susceptibility loci

We performed association tests while conditioning on the variants identified in the pooled and ancestry-specific results to determine if additional common or low-frequency variants showed distinct associations with T2D. Within a 500 KB region around the index variant, variants were considered to be distinct from the index variant if they remained significant in conditional analyses at locus-wide significance (p<1×10^−5^ and MAC>20). To identify potentially causal variants underlying each T2D association signal, we created a 95% credible set that had a 95% posterior probability of containing the causal variant. Within each region, we calculated a Bayes factor (BF) for each variant^48^. For genetic loci with multiple distinct association signals, the association P values were derived from conditional analyses. We thereafter calculated a posterior probability for each variant that drives the association signal through dividing its BF by the sum of BFs in the region. The 95% credible set for each association signal was eventually constructed by sorting the variants’ posterior probability in descending order and including variants until their cumulative posterior probability for association was over 95%^10^.

### Power calculations

Power was calculated using the genetic association study power calculator^49^. The following assumptions were used in power calculations: disease prevalence of 8%, significance level of 4×10-9, additive model, case n=9,639 and control n=34,494. Power is presented at levels of MAF commensurate with WGS single-variant association analyses (MAF=0.02%, 0.04%, 0.1%, 1% and %5); and OR from 1.0 - 6.0 (**Supplementary Figure 7**).

### Known variants

Known variants (n=403) representing 243 loci and genetic credible set regions at these loci were curated from DIAMANTE European, a large genome-wide, genotype and imputation-based T2D association study^10^. Twenty-three TOPMed variants did not pass genotyping quality, had MAC<20 or their genomic locations could not be updated to build 38 and were excluded from analyses. A known region was defined for each of the 380 available variants (239 loci) as the published genetic credible sets +/- 500kb around the variant (**Supplementary Table 16**).

## DATA AND CODE AVAILABILITY

The datasets generated during and/or analyzed during the current study are available in the AMP- T2D Portal, http://t2d.hugeamp.org/.

## TOPMed Acknowledgments

Molecular data for the Trans-Omics in Precision Medicine (TOPMed) program was supported by the National Heart, Lung and Blood Institute (NHLBI). See the TOPMed Omics Support Table for study specific omics support information (Supplementary Table). Core support including centralized genomic read mapping and genotype calling, along with variant quality metrics and filtering were provided by the TOPMed Informatics Research Center (3R01HL-117626-02S1; contract HHSN268201800002I). Core support including phenotype harmonization, data management, sample-identity QC, and general program coordination were provided by the TOPMed Data Coordinating Center (R01HL-120393; U01HL-120393; contract HHSN268201800001I). We gratefully acknowledge the studies and participants who provided biological samples and data for TOPMed.

## Study-Specific, Resource and Individual Acknowledgements

See Supplementary Table for study’s, individuals and resources funding support.

## AUTHOR CONTRIBUTIONS

J.W., and T.D.M. contributed equally to this work. J.W., T.D.M., H.M.H., S.R., M.D.S., N.R.H., P.S.D., X.Y., J.R., S.R., J.D., R.S., J.M. and A.K.M contributed to writing. J.W., T.D.M., S.R., J.A.B., M.M., A.K.M. analyzed data. J.W., T.D.M., H.M.H., S.R., M.D.S., N.R.H., P.S.D., J.A.B., J.P. J.R.O, M.M., S.L., P.W., A.K.M. contributed to analyses. A.K.M., J.W., L.L. contributed to the development of the trait harmonization protocol. Islet interaction and chromatin structure functional data: S.B-G., I.M-E., J.F. Islet regulation and expression functional data: R.D’O.A., A.V., S.P. J.A.B., C.S., D.D. provided results for cardiometabolic-related traits. A.M., M.I.Mc provided imputation results from the DIAMANTE Consortium. J.W., R.S., J.D., S.S.R., J.I.R., J.B.M., A.K.M. provided leadership.

## COMPETING INTEREST DECLARATION

|  |  |
| --- | --- |
| Anubha Mahajan | As of January 2020, A.M. is an employee of Genentech, and a holder of Roche stock. |
| Patrick T. Ellinor | Dr. Ellinor is supported by a grant from Bayer AG to the Broad Institute focused on the genetics and therapeutics of cardiovascular disease. Dr. Ellinor has consulted for Bayer AG, Novartis and Quest Diagnostics. |
| Jose Florez | Dr. Florez has received a consulting honorarium from Goldfinch Bio. and speaker fees from Novo Nordisk |
| James S. Floyd | Dr. Floyd has consulted for Shionogi Inc |
| Barry I. Freedman | Dr. Freedman is a consultant for RenalytixAI and AstraZeneca Pharmaceuticals |
| Steven A. Lubitz | Dr. Lubitz receives sponsored research support from Bristol Myers Squibb / Pfizer, Bayer HealthCare, and Boehringer Ingelheim, and has consulted for Abbott, Quest Diagnostics, Bristol Myers Squibb / Pfizer. |
| Michael H. Cho | Grant support from GSK; consulting fees from Genentech |
| Bruce M. Psaty | Psaty serves on the Steering Committee of the Yale Open Data Access Project funded by Johnson & Johnson. |
| Stella Aslibekyan | SA holds equity in 23andMe, Inc. |
| Mark I McCarthy | The views expressed in this article are those of the author(s) and not necessarily those of the NHS, the NIHR, or the Department of Health. M.I.Mc. has served on advisory panels for Pfizer, NovoNordisk and Zoe Global, has received honoraria from Merck, Pfizer, Novo Nordisk and Eli Lilly, and research funding from Abbvie, Astra Zeneca, Boehringer Ingelheim, Eli Lilly, Janssen, Merck, NovoNordisk, Pfizer, Roche, Sanofi Aventis, Servier, and Takeda. As of June 2019, M.I.Mc. is an employee of Genentech, and a holder of Roche stock. |

## ADDITIONAL INFORMATION

**Supplementary Information** is available for this paper.

Correspondence and requests for materials should be addressed to Jennifer Wessel and/or Alisa K Manning.

## Supplementary Note

### Supplementary Authors and Affiliations

NHLBI Trans-Omics for Precision Medicine (TOPMed) Consortium Banner Authors:

Namiko Abe^1^, Gonçalo Abecasis^2^, Francois Aguet^3^, Christine Albert^4^, Laura Almasy^5^, Alvaro Alonso^6^, Seth Ament^7^, Peter Anderson^8^, Pramod Anugu^9^, Deborah Applebaum-Bowden^10^, Kristin Ardlie^3^, Dan Arking^11^, Donna K Arnett^12^, Allison Ashley-Koch^13^, Stella Aslibekyan^14^, Tim Assimes^15^, Paul Auer^16^, Dimitrios Avramopoulos^11^, John Barnard^17^, Kathleen Barnes^18^, R Graham Barr^19^, Emily Barron-Casella^11^, Lucas Barwick^20^, Terri Beaty^11^, Gerald Beck^21^, Diane Becker^22^, Lewis Becker^11^, Rebecca Beer^23^, Amber Beitelshees^7^, Emelia Benjamin^24^, Takis Benos^25^, Marcos Bezerra^26^, Larry Bielak^2^, Joshua Bis^27^, Thomas Blackwell^2^, John Blangero^28^, Eric Boerwinkle^29^, Donald W Bowden^30^, Russell Bowler^31^, Jennifer Brody^8^, Ulrich Broeckel^32^, Jai Broome^8^, Karen Bunting^1^, Esteban Burchard^33^, Carlos Bustamante^34^, Erin Buth^35^, Brian Cade^36^, Jonathan Cardwell^37^, Vincent Carey^38^, Cara Carty^39^, Richard Casaburi^40^, James Casella^11^, Peter Castaldi^41^, Mark Chaffin^3^, Christy Chang^7^, Yi-Cheng Chang^42^, Daniel Chasman^43^, Sameer Chavan^37^, Bo-Juen Chen^1^, Wei-Min Chen^44^, Yii-Der Ida Chen^45^, Michael Cho^38^, Seung Hoan Choi^3^, Lee-Ming Chuang^46^, Mina Chung^47^, Ren-Hua Chung^48^, Clary Clish^49^, Suzy Comhair^50^, Matthew Conomos^35^, Elaine Cornell^51^, Adolfo Correa^52^, Carolyn Crandall^40^, James Crapo^53^, L Adrienne Cupples^54^, Joanne Curran^55^, Jeffrey Curtis^2^, Brian Custer^56^, Coleen Damcott^7^, Dawood Darbar^57^, Sayantan Das^2^, Sean David^58^, Colleen Davis^8^, Michelle Daya^37^, Mariza de Andrade^59^, Lisa de las Fuentes^60^, Michael DeBaun^61^, Ranjan Deka^62,63^, Dawn DeMeo^38^, Scott Devine^7^, Qing Duan^64^, Ravi Duggirala^65^, Jon Peter Durda^51^, Susan Dutcher^66^, Charles Eaton^67^, Lynette Ekunwe^9^, Adel El Boueiz^68^, Patrick Ellinor^69^, Leslie Emery^8^, Serpil Erzurum^17^, Charles Farber^44^, Tasha Fingerlin^70^, Matthew Flickinger^2^, Myriam Fornage^29^, Nora Franceschini^71^, Chris Frazar^8^, Mao Fu^7^, Stephanie M Fullerton^8^, Lucinda Fulton^66^, Stacey Gabriel^3^, Weiniu Gan^23^, Shanshan Gao^37^, Yan Gao^9^, Margery Gass^72^, Bruce Gelb^73^, Xiaoqi (Priscilla) Geng^2^, Mark Geraci^74^, Soren Germer^1^, Robert Gerszten^75^, Auyon Ghosh^38^, Richard Gibbs^76^, Chris Gignoux^15^, Mark Gladwin^25^, David Glahn^77^, Stephanie Gogarten^8^, Da-Wei Gong^7^, Harald Goring^78^, Sharon Graw^18^, Daniel Grine^37^, C Charles Gu^66^, Yue Guan^7^, Xiuqing Guo^45^, Namrata Gupta^3^, Jeff Haessler^72^, Michael Hall^9^, Daniel Harris^7^, Nicola L Hawley^79,63^, Jiang He^80^, Ben Heavner^35^, Susan Heckbert^8^, Ryan Hernandez^81^, David Herrington^82^, Craig Hersh^83^, Bertha Hidalgo^14^, James Hixson^29^, Brian Hobbs^38^, John Hokanson^37^, Elliott Hong^7^, Karin Hoth^84^, Chao (Agnes) Hsiung^85^, Yi-Jen Hung^86^, Haley Huston^87^, Chii Min Hwu^88^, Marguerite Ryan Irvin^14^, Rebecca Jackson^89^, Deepti Jain^8^, Cashell Jaquish^23^, Min A Jhun^2^, Jill Johnsen^90^, Andrew Johnson^23^, Craig Johnson^8^, Rich Johnston^6^, Kimberly Jones^11^, Hyun Min Kang^91^, Robert Kaplan^92^, Sharon Kardia^2^, Sekar Kathiresan^3^, Shannon Kelly^56^, Eimear Kenny^73^, Michael Kessler^7^, Alyna Khan^8^, Wonji Kim^93^, Greg Kinney^37^, Barbara Konkle^87^, Charles Kooperberg^72^, Holly Kramer^94^, Christoph Lange^95^, Ethan Lange^37^, Leslie Lange^37^, Cathy Laurie^8^, Cecelia Laurie^8^, Meryl LeBoff^38^, Jiwon Lee^38^, Seunggeun Shawn Lee^2^, Wen-Jane Lee^88^, Jonathon LeFaive^2^, David Levine^8^, Dan Levy^23^, Joshua Lewis^7^, Xiaohui Li^45^, Yun Li^64^, Henry Lin^45^, Honghuang Lin^96^, Keng Han Lin^2^, Xihong Lin^97^, Simin Liu^98^, Yongmei Liu^99^, Yu Liu^100^, Ruth J. F. Loos^101^, Steven Lubitz^69^, Kathryn Lunetta^96^, James Luo^23^, Michael Mahaney^55^, Barry Make^11^, Ani Manichaikul^44^, JoAnn Manson^38^, Lauren Margolin^3^, Lisa Martin^102^, Susan Mathai^37^, Rasika Mathias^11^, Susanne May^35^, Patrick McArdle^7^, Merry-Lynn McDonald^14^, Sean McFarland^93^, Stephen McGarvey^67,63^, Daniel McGoldrick^8^, Caitlin McHugh^35^, Hao Mei^9^, Luisa Mestroni^18^, Deborah A Meyers^103^, Julie Mikulla^23^, Nancy Min^9^, Mollie Minear^23^, Ryan L Minster^25,63^, Braxton D Mitchell^7^, Matt Moll^41^, May E Montasser^7^, Courtney Montgomery^104^, Arden Moscati^73^, Solomon Musani^52^, Stanford Mwasongwe^9^, Josyf C Mychaleckyj^44^, Girish Nadkarni^73^, Rakhi Naik^11^, Take Naseri^105,63^, Pradeep Natarajan^106^, Sergei Nekhai^107^, Sarah C Nelson^35^, Bonnie Neltner^37^, Deborah Nickerson^8^, Kari North^64^, Jeff O’Connell^108^, Tim O’Connor^7^, Heather Ochs-Balcom^109^, David T Paik^110^, Nicholette Palmer^111^, James Pankow^112^, George Papanicolaou^23^, Afshin Parsa^7^, Juan Manuel Peralta^65^, Marco Perez^15^, James Perry^7^, Ulrike Peters^113^, Patricia Peyser^2^, Lawrence S Phillips^6^, Toni Pollin^7^, Wendy Post^114^, Julia Powers Becker^115^, Meher Preethi Boorgula^37^, Michael Preuss^73^, Bruce Psaty^8^, Pankaj Qasba^23^, Dandi Qiao^38^, Zhaohui Qin^6^, Nicholas Rafaels^116^, Laura Raffield^117^, Vasan S Ramachandran^96^, D C Rao^66^, Laura Rasmussen-Torvik^118^, Aakrosh Ratan^44^, Susan Redline^38^, Robert Reed^7^, Elizabeth Regan^53^, Alex Reiner^119^, Muagututi’a Sefuiva Reupena^120^, Ken Rice^8^, Stephen Rich^44^, Dan Roden^121^, Carolina Roselli^3^, Jerome Rotter^45^, Ingo Ruczinski^11^, Pamela Russell^37^, Sarah Ruuska^87^, Kathleen Ryan^7^, Ester Cerdeira Sabino^122^, Danish Saleheen^19^, Shabnam Salimi^7^, Steven Salzberg^11^, Kevin Sandow^123^, Vijay G Sankaran^124^, Christopher Scheller^2^, Ellen Schmidt^2^, Karen Schwander^66^, David Schwartz^37^, Frank Sciurba^25^, Christine Seidman^125^, Jonathan Seidman^126^, Vivien Sheehan^127^, Stephanie L Sherman^128^, Amol Shetty^7^, Aniket Shetty^37^, Wayne Hui-Heng Sheu^88^, M Benjamin Shoemaker^129^, Brian Silver^130^, Edwin Silverman^38^, Jennifer Smith^2^, Josh Smith^8^, Nicholas Smith^131^, Tanja Smith^1^, Sylvia Smoller^92^, Beverly Snively^132^, Michael Snyder^15^, Tamar Sofer^38^, Nona Sotoodehnia^8^, Adrienne M Stilp^8^, Garrett Storm^37^, Elizabeth Streeten^7^, Jessica Lasky Su^38^, Yun Ju Sung^66^, Jody Sylvia^38^, Adam Szpiro^8^, Carole Sztalryd^7^, Daniel Taliun^2^, Hua Tang^133^, Margaret Taub^11^, Kent D Taylor^134^, Matthew Taylor^18^, Simeon Taylor^7^, Marilyn Telen^13^, Timothy A Thornton^8^, Machiko Threlkeld^135^, Lesley Tinker^72^, David Tirschwell^8^, Sarah Tishkoff^136^, Hemant Tiwari^137^, Catherine Tong^138^, Russell Tracy^139^, Michael Tsai^112^, Dhananjay Vaidya^11^, David Van Den Berg^140^, Peter VandeHaar^2^, Scott Vrieze^141^, Tarik Walker^37^, Robert Wallace^84^, Avram Walts^37^, Fei Fei Wang^8^, Heming Wang^142^, Karol Watson^40^, Daniel E Weeks^25,63^, Bruce Weir^8^, Scott Weiss^38^, Lu-Chen Weng^69^, Jennifer Wessel^143^, Cristen Willer^144^, Kayleen Williams^35^, L Keoki Williams^145^, Carla Wilson^38^, James Wilson^146^, Quenna Wong^8^, Joseph Wu^110^, Huichun Xu^7^, Lisa Yanek^11^, Ivana Yang^37^, Rongze Yang^7^, Norann Zaghloul^7^, Maryam Zekavat^3^, Yingze Zhang^147^, Snow Xueyan Zhao^53^, Wei Zhao^148^, Degui Zhi^29^, Xiang Zhou^2^, Xiaofeng Zhu^149^, Michael Zody^1^, Sebastian Zoellner^2^

^1^New York Genome Center, New York, New York, 10013, US, ^2^University of Michigan, Ann Arbor, Michigan, 48109, US, ^3^Broad Institute, Cambridge, Massachusetts, 2142, US, ^4^Brigham & Women’s Hospital, Cedars Sinai, Boston, Massachusetts, 2114, US, ^5^Children’s Hospital of Philadelphia, University of Pennsylvania, Philadelphia, Pennsylvania, 19104, US, ^6^Emory University, Atlanta, Georgia, 30322, US, ^7^University of Maryland, Baltimore, Maryland, 21201, US, ^8^University of Washington, Seattle, Washington, 98195, US, ^9^University of Mississippi, Jackson, Mississippi, 38677, US, ^10^National Institutes of Health, Bethesda, Maryland, 20892, US, ^11^Johns Hopkins University, Baltimore, Maryland, 21218, US, ^12^University of Kentucky, Lexington, Kentucky, 40506, US, ^13^Duke University, Durham, North Carolina, 27708, US, ^14^University of Alabama, Birmingham, Alabama, 35487, US, ^15^Stanford University, Stanford, California, 94305, US, ^16^University of Wisconsin Milwaukee, Milwaukee, Wisconsin, 53211, US, ^17^Cleveland Clinic, Cleveland, Ohio, 44195, US, ^18^University of Colorado Anschutz Medical Campus, Aurora, Colorado, 80045, US, ^19^Columbia University, New York, New York, 10027, US, ^20^LTRC, The Emmes Corporation, Rockville, Maryland, 20850, US, ^21^Quantitative Health Sciences, Cleveland Clinic, Cleveland, Ohio, 44195, US, ^22^Medicine, Johns Hopkins University, Baltimore, Maryland, 21218, US, ^23^National Heart, Lung, and Blood Institute, National Institutes of Health, Bethesda, Maryland, 20892, US, ^24^Boston University School of Medicine, Boston University, Massachusetts General Hospital, Boston, Massachusetts, 2114, US, ^25^University of Pittsburgh, Pittsburgh, Pennsylvania, 15260, US, ^26^Fundação de Hematologia e Hemoterapia de Pernambuco - Hemope, Recife, 52011-000, BR, ^27^Cardiovascular Health Research Unit, Department of Medicine, University of Washington, Seattle, Washington, 98195, US, ^28^Human Genetics, University of Texas Rio Grande Valley School of Medicine, Brownsville, Texas, 78520, US, ^29^University of Texas Health at Houston, Houston, Texas, 77225, US, ^30^Department of Biochemistry, Wake Forest Baptist Health, Winston-Salem, North Carolina, 27157, US, ^31^National Jewish Health, National Jewish Health, Denver, Colorado, 80206, US, ^32^Medical College of Wisconsin, Milwaukee, Wisconsin, 53226, US, ^33^University of California, San Francisco, San Francisco, California, 94143, US, ^34^Biomedical Data Science, Stanford University, Stanford, California, 94305, US, ^35^Biostatistics, University of Washington, Seattle, Washington, 98195, US, ^36^Brigham and Women’s Hospital, Brigham & Women’s Hospital, Boston, Massachusetts, 2115, US, ^37^University of Colorado at Denver, Denver, Colorado, 80204, US, ^38^Brigham & Women’s Hospital, Boston, Massachusetts, 2115, US, ^39^Washington State University, Pullman, Washington, 99164, US, ^40^University of California, Los Angeles, Los Angeles, California, 90095, US, ^41^Medicine, Brigham & Women’s Hospital, Boston, Massachusetts, 2115, US, ^42^National Taiwan University, Taipei, 10617, TW, ^43^Division of Preventive Medicine, Brigham & Women’s Hospital, Boston, Massachusetts, 2215, US, ^44^University of Virginia, Charlottesville, Virginia, 22903, US, ^45^Lundquist Institute, Torrance, California, 90502, US, ^46^National Taiwan University Hospital, National Taiwan University, Taipei, 10617, TW, ^47^Cleveland Clinic, Cleveland Clinic, Cleveland, Ohio, 44195, US, ^48^National Health Research Institute Taiwan, Miaoli County, 350, TW, ^49^Metabolomics Platform, Broad Institute, Cambridge, Massachusetts, 2142, US, ^50^Immunity and Immunology, Cleveland Clinic, Cleveland, Ohio, 44195, US, ^51^University of Vermont, Burlington, Vermont, 5405, US, ^52^Medicine, University of Mississippi, Jackson, Mississippi, 38677, US, ^53^National Jewish Health, Denver, Colorado, 80206, US, ^54^Biostatistics, Boston University, Boston, Massachusetts, 2115, US, ^55^University of Texas Rio Grande Valley School of Medicine, Brownsville, Texas, 78520, US, ^56^Vitalant Research Institute, San Francisco, California, 94118, US, ^57^University of Illinois at Chicago, Chicago, Illinois, 60607, US, ^58^University of Chicago, Chicago, Illinois, 60637, US, ^59^Health Sciences Research, Mayo Clinic, Rochester, Minnesota, 55905, US, ^60^Department of Medicine, Cardiovascular Division, Washington University in St Louis, St. Louis, Missouri, 63110, US, ^61^Vanderbilt University, Nashville, Tennessee, 37235, US, ^62^University of Cincinnati, Cincinnati, Ohio, 45220, US, ^63^The Samoan Obesity, Lifestyle and Genetic Adaptations Study (OLaGA) Group, ^64^University of North Carolina, Chapel Hill, North Carolina, 27599, US, ^65^University of Texas Rio Grande Valley School of Medicine, Edinburg, Texas, 78539, US, ^66^Washington University in St Louis, St Louis, Missouri, 63130, US, ^67^Brown University, Providence, Rhode Island, 2912, US, ^68^Channing Division of Network Medicine, Harvard University, Cambridge, Massachusetts, 2138, US, ^69^Massachusetts General Hospital, Boston, Massachusetts, 2114, US, ^70^Center for Genes, Environment and Health, National Jewish Health, Denver, Colorado, 80206, US, ^71^Epidemiology, University of North Carolina, Chapel Hill, North Carolina, 27599, US, ^72^Fred Hutchinson Cancer Research Center, Seattle, Washington, 98109, US, ^73^Icahn School of Medicine at Mount Sinai, New York, New York, 10029, US, ^74^Medicine, Indiana University, Indianapolis, Indiana, 46202, US, ^75^Beth Israel Deaconess Medical Center, Boston, Massachusetts, 2215, US, ^76^Baylor College of Medicine Human Genome Sequencing Center, Houston, Texas, 77030, US, ^77^Yale University, New Haven, Connecticut, 6520, US, ^78^University of Texas Rio Grande Valley School of Medicine, San Antonio, Texas, 78229, US, ^79^Department of Chronic Disease Epidemiology, Yale University, New Haven, Connecticut, 6520, US, ^80^Tulane University, New Orleans, Louisiana, 70118, US, ^81^University of California, San Francisco, Montreal, H3A 0G4, CA, ^82^Wake Forest Baptist Health, Winston-Salem, North Carolina, 27157, US, ^83^Channing Division of Network Medicine, Brigham & Women’s Hospital, Boston, Massachusetts, 2115, US, ^84^University of Iowa, Iowa City, Iowa, 52242, US, ^85^Institute of Population Health Sciences, NHRI, National Health Research Institute Taiwan, Miaoli County, 350, TW, ^86^Tri-Service General Hospital National Defense Medical Center, TW, ^87^Blood Works Northwest, Seattle, Washington, 98104, US, ^88^Taichung Veterans General Hospital Taiwan, Taichung City, 407, TW, ^89^Internal Medicine, DIvision of Endocrinology, Diabetes and Metabolism, Ohio State University Wexner Medical Center, Columbus, Ohio, 43210, US, ^90^Blood Works Northwest, University of Washington, Seattle, Washington, 98104, US, ^91^Biostatistics, University of Michigan, Ann Arbor, Michigan, 48109, US, ^92^Albert Einstein College of Medicine, New York, New York, 10461, US, ^93^Harvard University, Cambridge, Massachusetts, 2138, US, ^94^Public Health Sciences, Loyola University, Maywood, Illinois, 60153, US, ^95^Biostats, Harvard School of Public Health, Boston, Massachusetts, 2115, US, ^96^Boston University, Boston, Massachusetts, 2215, US, ^97^Harvard School of Public Health, Boston, Massachusetts, 2115, US, ^98^Epidemiology and Medicine, Brown University, Providence, Rhode Island, 2912, US, ^99^Cardiology, Duke University, Durham, North Carolina, 27708, US, ^100^Cardiovascular Institute, Stanford University, Stanford, California, 94305, US, ^101^The Charles Bronfman Institute for Personalized Medicine, Icahn School of Medicine at Mount Sinai, New York, New York, 10029, US, ^102^George Washington University, Washington, District of Columbia, 20052, US, ^103^University of Arizona, Tucson, Arizona, 85721, US, ^104^Genes and Human Disease, Oklahoma Medical Research Foundation, Oklahoma City, Oklahoma, 73104, US, ^105^Ministry of Health, Government of Samoa, Apia, WS, ^106^Broad Institute, Harvard University, Massachusetts General Hospital, Boston, Massachusetts, 2114, US, ^107^Howard University, Washington, District of Columbia, 20059, US, ^108^University of Maryland, Baltimore, Maryland, 21201, US, ^109^University at Buffalo, Buffalo, New York, 14260, US, ^110^Stanford Cardiovascular Institute, Stanford University, Stanford, California, 94305, US, ^111^Biochemistry, Wake Forest Baptist Health, Winston-Salem, North Carolina, 27157, US, ^112^University of Minnesota, Minneapolis, Minnesota, 55455, US, ^113^Fred Hutchinson Cancer Research Center, University of Washington, Seattle, Washington, 98195, US, ^114^Cardiology/Medicine, Johns Hopkins University, Baltimore, Maryland, 21218, US, ^115^Medicine, University of Colorado at Denver, Denver, Colorado, 80204, US, ^116^University of Colorado at Denver, Denver, Colorado, 80045, US, ^117^Genetics, University of North Carolina, Chapel Hill, North Carolina, 27599, US, ^118^Northwestern University, Chicago, Illinois, 60208, US, ^119^Fred Hutchinson Cancer Research Center, University of Washington, Seattle, Washington, 98109, US, ^120^Lutia I Puava Ae Mapu I Fagalele, Apia, WS, ^121^Medicine, Pharmacology, Biomedicla Informatics, Vanderbilt University, Nashville, Tennessee, 37235, US, ^122^Faculdade de Medicina, Universidade de Sao Paulo, Sao Paulo, 1310000, BR, ^123^TGPS, Lundquist Institute, Torrance, California, 90502, US, ^124^Division of Hematology/Oncology, Broad Institute, Harvard University, Cambridge, Massachusetts, 2142, US, ^125^Genetics, Harvard Medical School, Boston, Massachusetts, 2115, US, ^126^Harvard Medical School, Boston, Massachusetts, 2115, US, ^127^Pediatrics, Baylor College of Medicine, Houston, Texas, 77030, US, ^128^Human Genetics, Emory University, Atlanta, Georgia, 30322, US, ^129^Medicine/Cardiology, Vanderbilt University, Nashville, Tennessee, 37235, US, ^130^UMass Memorial Medical Center, Worcester, Massachusetts, 1655, US, ^131^Epidemiology, University of Washington, Seattle, Washington, 98195, US, ^132^Biostatistical Sciences, Wake Forest Baptist Health, Winston-Salem, North Carolina, 27157, US, ^133^Genetics, Stanford University, Stanford, California, 94305, US, ^134^Institute for Translational Genomics and Populations Sciences, Lundquist Institute, Torrance, California, 90502, US, ^135^University of Washington, Department of Genome Sciences, University of Washington, Seattle, Washington, 98195, US, ^136^Genetics, University of Pennsylvania, Philadelphia, Pennsylvania, 19104, US, ^137^Biostatistics, University of Alabama, Birmingham, Alabama, 35487, US, ^138^Department of Biostatistics, University of Washington, Seattle, Washington, 98195, US, ^139^Pathology & Laboratory Medicine, University of Vermont, Burlington, Vermont, 5405, US, ^140^USC Methylation Characterization Center, University of Southern California, University of Southern California, California, 90033, US, ^141^University of Colorado at Boulder, University of Minnesota, Boulder, Colorado, 80309, US, ^142^Brigham & Women’s Hospital, Partners.org, Boston, Massachusetts, 2115, US, ^143^Epidemiology, Indiana University, Indianapolis, Indiana, 46202, US, ^144^Internal Medicine, University of Michigan, Ann Arbor, Michigan, 48109, US, ^145^Henry Ford Health System, Detroit, Michigan, 48202, US, ^146^Cardiology, Beth Israel Deaconess Medical Center, Boston, Massachusetts, 2215, US, ^147^Medicine, University of Pittsburgh, Pittsburgh, Pennsylvania, 15260, US, ^148^Department of Epidemiology, University of Michigan, Ann Arbor, Michigan, 48109, US, ^149^Department of Population and Quantitative Health Sciences, Case Western Reserve University, Cleveland, Ohio, 44106, US

### Supplementary Results

#### Enrichment of rare variant signals in islet enriched genes

We also tested the coding genome, to understand if islet expressed genes were enriched for rare variant associations with T2D. We gathered results from the three coding variant aggregation strategies (all missense, deleterious missense, and loss of function) and performed two sample Kolmogorov-Smirnov (KS) tests for each analysis. Association P values were compared between islet expressed genes (fragments per kilobase of exon per million reads mapped, FPKM > 2) and genes not expressed in islets with a single tailed KS-test to determine if their distributions differed. Correcting for the four models tested in each ancestry group or study population and aggregation strategy pair, the loss of function aggregation tests from islet expressed genes were enriched for low P values within the Hispanic/Latinx ancestry group (P = 0.0054; burden test, unadjusted for BMI; **Supplementary Table 11**). No other results show significant deviation between the two groups of genes.

#### GWAS level of significance for T2D outcome

We tabulated loci passing GWAS level of significance (i.e. p<5×10^−8^, **Supplementary Table 13**) and identified an additional 8 loci (9 variants) of which six variants have not been previously reported. Of the 6 variants not previously identified, 2 were rare or low-frequency (MAF<5%) and the other four were identified in historically underrepresented populations (i.e. non-European). For variants that are in regions not previously identified we explored whether they could have been missed by GWAS or imputation. Using LDlink (https://ldlink.nci.nih.gov/?tab=home) we found five of the six variants are not on any of the arrays; rs78479678 is available on the exome chip and newer arrays. Imputation quality was good (>0.90) for only two of the variants (chr1:86325383:T:A and rs78479678) and associations were not observed in the DIAMANTE EU study (**Supplementary Table 17**).

Pooled analyses yielded more WGS-wide significant results than meta-analyses; and results from meta-analyses were all identified by pooled analyses (**Supplementary Tables 21-22**).

#### Associations with related cardiometabolic traits and by carrier status

We examined whether our novel variants were also associated with related cardiometabolic traits – FG, FI, HbA1c and BMI in other TOPMed analysis samples. Several rare T2D risk-raising alleles showed nominally significant association with these traits: rs145197571 at the *MRPL46*/*MRPS11* locus was associated with higher BMI in African-ancestry individuals (P=8.1×10^−3^); rs200622604 at the *NR4A2*/*GPD2* locus was associated with higher FI in European ancestry (P=6.1×10^−3^). The low-frequency allele at the *CCND2* locus, rs76895963, was protective for T2D risk and was also associated with lower FG (P1×10^−7^), lower FI (P=4×10^−3^), lower HbA1c (2.8×10^−4^) and higher BMI in the pooled sample (P=4.7×10^−4^; **Supplementary Table 23**).

We next compared carriers versus non-carriers of T2D-associated alleles with regard to the last available FG, HbA1C and BMI values for non-diabetic individuals and the age and BMI at T2D diagnosis for individuals with T2D, stratified by cohort and ancestry (**Supplementary Table 24)**. We observed that European-ancestry carriers of the T2D-risk decreasing allele of rs76895963 at the *CCND2* locus had decreased last-available HbA1c levels (HbA1c_carriers_=5.4; HbA1c_non-carriers_=5.5; P=0.3.8×10^−4^), increased last-available BMI levels in individuals without T2D (BMI_carriers_ = 28.7; BMI_non-carrier_=28.1; P=0.002) and also in individuals with T2D (BMI_carriers_ = 33.3; BMI_noncarrier_=31.2; P=0.004). In the Samoan Study, individuals with T2D who were carriers of the T2D risk increasing allele of driver variant rs929186279 from the *PSD4*/*PAX8-AS1* locus had marginally significant earlier onset of T2D (38±9.6 vs 49±8.2, P=0.06) than non-carriers. Interestingly, Hispanic non-diabetic carriers of the T2D risk increasing allele of driver variant rs200945165 at the *NR4A2*/*GPD2* locus had lower BMI than non-carriers (N_carriers_=22 BMI_carriers_ = 27.3; BMI_noncarrier_=30.1; P=0.008). Additionally, Hispanic-ancestry, non-diabetic carriers of the rs11992463 variant at the KCNV1 locus had higher FG (5.5±0.6 vs 5.2±0.6, p=0.03) than non-carriers.

### T2D+, an expanded T2D outcome

#### Heritability Analysis

We also conducted heritability analyses with the T2D+ outcome using the same methods (assuming a prevalence of 11%, **Supplementary Table 6 and Supplementary Figure 2)**. The contribution of rare, non-coding variants in the 2^nd^ LD score quartile had the largest proportional heritability, estimated to be 35% (95% confidence interval [CI] 0.11-0.59, P=1.6×10^−3^), with additional contributions from common variants (9%, 95%CI 0.04-0.13, P=1.7×10^−4^, **Supplementary Table 7**). Furthermore, we found results were similar to T2D, except we identified additional contributions to T2D+ heritability from low-frequency variants annotated as ‘*islet interaction and expression promoter’* in the 1^st^ LD quartile (2%, 95%CI 0-0.04, P=7.1×10^−3^), and ultra-rare variants in the 3^rd^ LD quartile annotated as ‘*islet interaction and expression enhancer’* (11%, 95%CI 0.03-0.18, P=4.2×10^−3^) and annotated as ‘*islet interaction and chromatin structure enhancer’* (7%, 95%CI 0.02-0.12, P=1.7×10^−3^).

#### Single Variant Analyses

In single variant analyses of T2D+, we identified 10 variants at 9 loci in either pooled or ancestry-specific analyses at WGS-wide level of significance (P<4×10^−9^) or locus-wide significance (P<1×10^−5^; **Supplementary Table 14, Supplementary Figures 5 and 10**). We also looked at sub-significant associations, variants passing GWAS level of significance (i.e. P<5×10^−8^, **Supplementary Table 25**) and identified an additional 7 loci (9 variants). Four of these loci have been previously reported. The other 4 loci are either from diverse ancestry groups (i.e. AF or HSL) or were rare (MAF<.01). Using the T2D+ definition identified the known glycemia loci, MTNRB1 and GCK, suggesting a role for glycemia. Furthermore, NWD2 has not previously been reported and the variant, rs10028027, is available on commercial SNP arrays.

Credible set analyses of whole genome sequencing and T2D+ identified one locus, MTNR1B, where the 95% posterior probability exceeds 0.9 for only one variant, rs10830963 (**Supplementary Table 26**). For the remaining five loci, multiple variants within a locus demonstrated moderate 95%PP (posterior probability) (i.e. <0.90). Of note, at the established GCK locus, rs2300584 has been reported as the index variant multiple times by previous GWAS^1^ shows low 95%PP=0.03 while other variants in our dataset have higher 95%PP (∼0.20).

We found more associations with the T2D+ outcome, and associations across the 2 outcomes were 90% concordant (*DUSP9*, a known locus, was not significant with T2D+). For those variants uniquely associated with the T2D+ outcome (i.e. not passing WGS level of significance for association with the T2D outcome) results with the T2D outcome are summarized in **Supplementary Table 27**; and P values are significant to1×10^−3^ suggesting these may also be T2D or glycemia risk variants.

### Supplementary Methods

#### Trait Harmonization Algorithm

We developed an algorithm to remove duplicate samples in the data set based on sequencing quality, study type and availability of phenotype data. By using each individual’s phenotype data from participating studies, we determined whether they represented monozygotic twins or an individual participating in multiple studies. For these individuals we made decisions about which phenotype data to keep for analyses by giving preference to studies with longitudinal data and/or by availability of type 2 diabetes (T2D) status. Scripts for harmonizing study data and creating a final dataset for analysis are available https://github.com/manning-lab/topmed-t2d-wg-trait-harmonization.

#### LD Score Regression from previously published GWAS summary statistics

Enrichment analyses from previously performed GWAS^2-5^ were performed using LD score regression (LDSC), which can be used to partition heritability by functional annotation and identify those annotations explaining an outsized proportion of heritability than expected by chance^6^. LD was estimated using the 1000 Genomes Phase 1 data from the CEU population for European ancestry GWAS studies (MAGIC and DIAGRAM) and from the ASW population for African ancestry GWAS studies (AAGILE and MEDIA).

The LDSC authors provided a full baseline model – a set of 53 genomic annotations curated from several sources (Table 3 of their publication) which are not specific to any cell types^6^. Redundant annotations contained in the baseline model, generated by extending regional boundaries by 500bp, were removed from primary analyses to remove redundant annotations. These extended annotations were, however, included in subsequent sensitivity analysis.

In addition to the baseline model annotations from LDSC, we included 68 tissue-specific annotations based on GenoSkyline Plus scores (**Supplementary Figure 11**)^7^. GenoSkyline Plus scores, interpreted as the posterior probability of a variant being functional in a tissue based on publicly available epigenomic data, range between 0 and 1 for all variants. We defined variants as belonging to a tissue-specific annotation if the GenoSkyline Plus score was greater than 0.5. Our results should not be sensitive to the choice of threshold, as GenoSkyline Plus scores generally have a bimodal distribution^8^.

#### Heritability Analysis

A major advantage of using WGS data to estimate variant-based heritability is that causal variants are directly ascertained in the sample. Evans *et al*. carefully considered heritability estimation methods for WGS data and describe a bias in estimates when stratification exists within samples and the MAF and linkage disequilibrium (LD) patterns of all variants do not match the MAF and LD patterns of the causal variants^9,10^. Multi-component methods implemented in GCTA correct this bias by binning variants by MAF and LD score, a metric of the amount of LD between variants, and jointly estimating the heritability of each component. This allows assessment of the contribution of variants that have lower LD scores (the 1^st^ and 2^nd^ LD score quartiles) and therefore may not have been captured by prior array based methods that depended on imputation from reference panels available at the time^1^.

#### Rare variant aggregation and association analysis

We developed a rare variant aggregation strategy for association testing built from the previously described *Gene-centric* aggregation. To further restrict aggregated variants to those likely functional within regulatory regions, we integrated predicted transcription factor binding sites (TFBS)^11^ and position frequency matrices (PFM)^12^. We created a set of predicted TFBS that fall within ATAC-seq peaks in pancreatic islets and also are within enhancer and promoter regions. We considered only TFBS where corresponding transcription factors were likely present within pancreatic islet tissue, determined by islet gene expression with average fragments per kilobase of exon per million reads mapped (FPKM) greater than 2^13-15^. Within each TFBS, base pair positions were filtered by the corresponding PFM^12^ to sites with information content > 1 in order to focus on regions vital in transcription factor binding efficacy^12^. Association analysis was performed in an identical manner to other aggregation strategies. No significant associations were observed in any ancestry groups.

Enrichment of exonic, rare-variant association signal within islet-expressed genes was determined by performing Kolmogov-Smirnov (KS) tests, comparing association P values between genes expressed in pancreatic islets and all other genes. There were 60 total sets of association results after considering coding variant aggregation strategy (all missense, deleterious missense, and loss of function), each ancestry or study population, and statistical model (SKAT/Burden and BMI adjustment). Within each set of results, genes were grouped by islet expression (FPKM > 2 or FPKM < 2). The distribution of association P values were compared between the two groups using a two sample, single tailed KS-test with the alternative hypothesis of lower P values within islet-expressed genes.

#### Association analyses with related cardiometabolic traits

Novel T2D variants were also considered for association with fasting glucose (FG), fasting insulin (FI), hemoglobin A1c (HbA1c) and body mass index (BMI), in collaboration with the TOPMed Anthropometric working group (**Supplementary Table 23**). Analyses of FG and log-transformed FI were examined in individuals without T2D; and adjusted for age, age-squared, BMI, sex, and study-ancestry (study and ancestry combined into a single variable), and accounting for relatedness using a genetic relatedness matrix (GRM). Association analysis with HbA1c was stratified by ancestry and meta-analyzed for pooled estimates across ancestries, adjusting for age, sex, and study, and included a random effect for study and a GRM to account for relatedness^16^. Association analysis with BMI was performed by creating BMI residuals, adjusted for age, age squared, study and 10 PCs; and were created within ancestry and sex strata, then rank-normal transformed and rescaled by strata variance. Pooled residuals were analyzed with linear mixed models including a variance component associated with the GRM plus separate residual variance components for each sex-ancestry group. Analyses were performed pooled across ancestries and additionally stratified by ancestry.

## REFERENCES

1. Collaboration, N. C. D. R. F. Worldwide trends in diabetes since 1980: a pooled analysis of 751 population-based studies with 4.4 million participants. Lancet 387, 1513–1530, doi:10.1016/S0140-6736(16)00618-8 (2016).

2. Taliun, D. et al. Sequencing of 53,831 diverse genomes from the NHLBI TOPMed Program. Nature Accepted (2020).

3. Diabetes Epigenome Atlas, <https://www.t2depigenome.org/> (Accessed September 11, 2018).

4. Fadista, J. et al. Global genomic and transcriptomic analysis of human pancreatic islets reveals novel genes influencing glucose metabolism. Proc Natl Acad Sci U S A 111, 13924–13929, doi:10.1073/pnas.1402665111 (2014).

5. Varshney, A. et al. Genetic regulatory signatures underlying islet gene expression and type 2 diabetes. Proc Natl Acad Sci U S A 114, 2301–2306, doi:10.1073/pnas.1621192114 (2017).

6. Miguel-Escalada, I. et al. Human pancreatic islet three-dimensional chromatin architecture provides insights into the genetics of type 2 diabetes. Nat Genet 51, 1137–1148, doi:10.1038/s41588-019-0457-0 (2019).

7. Mahajan, A. et al. Trans-ancestry genetic study of type 2 diabetes highlights the power of diverse populations for discovery and translation. medRxiv, 2020.2009.2022.20198937, doi:10.1101/2020.09.22.20198937 (2020).

8. Vujkovic, M. et al. Discovery of 318 new risk loci for type 2 diabetes and related vascular outcomes among 1.4 million participants in a multi-ancestry meta-analysis. Nat Genet 52, 680–691, doi:10.1038/s41588-020-0637-y (2020).

9. Willemsen, G. et al. The Concordance and Heritability of Type 2 Diabetes in 34,166 Twin Pairs From International Twin Registers: The Discordant Twin (DISCOTWIN) Consortium. Twin Res Hum Genet 18, 762–771, doi:10.1017/thg.2015.83 (2015).

10. Mahajan, A. et al. Fine-mapping type 2 diabetes loci to single-variant resolution using high-density imputation and islet-specific epigenome maps. Nat Genet 50, 1505–1513, doi:10.1038/s41588-018-0241-6 (2018).

11. Wessel, J. et al. Low-frequency and rare exome chip variants associate with fasting glucose and type 2 diabetes susceptibility. Nat Commun 6, 5897, doi:10.1038/ncomms6897 (2015).

12. Mahajan, A. et al. Identification and functional characterization of G6PC2 coding variants influencing glycemic traits define an effector transcript at the G6PC2-ABCB11 locus. PLoS Genet 11, e1004876, doi:10.1371/journal.pgen.1004876 (2015).

13. Fuchsberger, C. et al. The genetic architecture of type 2 diabetes. Nature 536, 41–47, doi:10.1038/nature18642 (2016).

14. Jun, G. et al. Evaluating the contribution of rare variants to type 2 diabetes and related traits using pedigrees. Proc Natl Acad Sci U S A 115, 379–384, doi:10.1073/pnas.1705859115 (2018).

15. Flannick, J. et al. Exome sequencing of 20,791 cases of type 2 diabetes and 24,440 controls. Nature 570, 71–76, doi:10.1038/s41586-019-1231-2 (2019).

16. Mahajan, A. et al. Refining the accuracy of validated target identification through coding variant fine-mapping in type 2 diabetes. Nat Genet 50, 559–571, doi:10.1038/s41588-018-0084-1 (2018).

17. Thurner, M. et al. Integration of human pancreatic islet genomic data refines regulatory mechanisms at Type 2 Diabetes susceptibility loci. Elife 7, doi:10.7554/eLife.31977 (2018).

18. Kryukov, G. V., Pennacchio, L. A. & Sunyaev, S. R. Most rare missense alleles are deleterious in humans: implications for complex disease and association studies. Am J Hum Genet 80, 727–739, doi:10.1086/513473 (2007).

19. Bonas-Guarch, S. et al. Re-analysis of public genetic data reveals a rare X-chromosomal variant associated with type 2 diabetes. Nat Commun 9, 321, doi:10.1038/s41467-017-02380-9 (2018).

20. Fishilevich, S. et al. GeneHancer: genome-wide integration of enhancers and target genes in GeneCards. Database (Oxford) 2017, doi:10.1093/database/bax028 (2017).

21. Leong, A. et al. Prediction of Type 2 Diabetes by Hemoglobin A1c in Two Community-Based Cohorts. Diabetes Care 41, 60–68, doi:10.2337/dc17-0607 (2018).

22. Chen, H. et al. Control for Population Structure and Relatedness for Binary Traits in Genetic Association Studies via Logistic Mixed Models. Am J Hum Genet 98, 653–666, doi:10.1016/j.ajhg.2016.02.012 (2016).

23. Zhou, W. et al. Efficiently controlling for case-control imbalance and sample relatedness in large-scale genetic association studies. Nat Genet 50, 1335–1341, doi:10.1038/s41588-018-0184-y (2018).

24. Gogarten, S. M. et al. Genetic association testing using the GENESIS R/Bioconductor package. Bioinformatics 35, 5346–5348, doi:10.1093/bioinformatics/btz567 (2019).

25. Willer, C. J., Li, Y. & Abecasis, G. R. METAL: fast and efficient meta-analysis of genomewide association scans. Bioinformatics 26, 2190–2191, doi:10.1093/bioinformatics/btq340 (2010).

26. Gaulton, K. J. et al. A map of open chromatin in human pancreatic islets. Nat Genet 42, 255–259, doi:10.1038/ng.530 (2010).

27. Wainschtein, P. et al. Recovery of trait heritability from whole genome sequence data. https://www.biorxiv.org/content/10.1101/588020v1.abstract, doi:https://doi.org/10.1101/588020(2019).

## METHODS REFERENCES

28. Brody, J. A. et al. Analysis commons, a team approach to discovery in a big-data environment for genetic epidemiology. Nat Genet 49, 1560–1563, doi:10.1038/ng.3968 (2017).

29. Conomos, M. P., Miller, M. B. & Thornton, T. A. Robust inference of population structure for ancestry prediction and correction of stratification in the presence of relatedness. Genet Epidemiol 39, 276–293, doi:10.1002/gepi.21896 (2015).

30. (Hail Program Team).

31. Liu, X. et al. WGSA: an annotation pipeline for human genome sequencing studies. J Med Genet 53, 111–112, doi:10.1136/jmedgenet-2015-103423 (2016).

32. GeneHancer database (v4.6), < https://genecards.weizmann.ac.il/geneloc_prev/index.shtml> (Accessed September 2018).

33. Javierre, B. M. et al. Lineage-Specific Genome Architecture Links Enhancers and Non-coding Disease Variants to Target Gene Promoters. Cell 167, 1369–1384 e1319, doi:10.1016/j.cell.2016.09.037 (2016).

34. Buenrostro, J. D., Giresi, P. G., Zaba, L. C., Chang, H. Y. & Greenleaf, W. J. Transposition of native chromatin for fast and sensitive epigenomic profiling of open chromatin, DNA-binding proteins and nucleosome position. Nat Methods 10, 1213–1218, doi:10.1038/nmeth.2688 (2013).

35. Johnson, D. S., Mortazavi, A., Myers, R. M. & Wold, B. Genome-wide mapping of in vivo protein-DNA interactions. Science 316, 1497–1502, doi:10.1126/science.1141319 (2007).

36. McLaren, W. et al. The Ensembl Variant Effect Predictor. Genome Biol 17, 122, doi:10.1186/s13059-016-0974-4 (2016).

37. Chun, S. & Fay, J. C. Identification of deleterious mutations within three human genomes. Genome Res 19, 1553–1561, doi:10.1101/gr.092619.109 (2009).

38. Schwarz, J. M., Cooper, D. N., Schuelke, M. & Seelow, D. MutationTaster2: mutation prediction for the deep-sequencing age. Nat Methods 11, 361–362, doi:10.1038/nmeth.2890 (2014).

39. Adzhubei, I. A. et al. A method and server for predicting damaging missense mutations. Nat Methods 7, 248–249, doi:10.1038/nmeth0410-248 (2010).

40. Kumar, P., Henikoff, S. & Ng, P. C. Predicting the effects of coding non-synonymous variants on protein function using the SIFT algorithm. Nat Protoc 4, 1073–1081, doi:10.1038/nprot.2009.86 (2009).

41. Yang, J. et al. Genetic variance estimation with imputed variants finds negligible missing heritability for human height and body mass index. Nat Genet 47, 1114–1120, doi:10.1038/ng.3390 (2015).

42. Evans, L. M. et al. Comparison of methods that use whole genome data to estimate the heritability and genetic architecture of complex traits. Nat Genet 50, 737–745, doi:10.1038/s41588-018-0108-x (2018).

43. Bulik-Sullivan, B. K. et al. LD Score regression distinguishes confounding from polygenicity in genome-wide association studies. Nat Genet 47, 291–295, doi:10.1038/ng.3211 (2015).

44. Lee, S. H., Wray, N. R., Goddard, M. E. & Visscher, P. M. Estimating missing heritability for disease from genome-wide association studies. Am J Hum Genet 88, 294–305, doi:10.1016/j.ajhg.2011.02.002 (2011).

45. Browning, S. R. & Browning, B. L. Population structure can inflate SNP-based heritability estimates. Am J Hum Genet 89, 191-193; author reply 193-195, doi:10.1016/j.ajhg.2011.05.025 (2011).

46. Xu, C. et al. Estimating genome-wide significance for whole-genome sequencing studies. Genet Epidemiol 38, 281–290, doi:10.1002/gepi.21797 (2014).

47. Fadista, J., Manning, A. K., Florez, J. C. & Groop, L. The (in)famous GWAS P-value threshold revisited and updated for low-frequency variants. Eur J Hum Genet 24, 1202–1205, doi:10.1038/ejhg.2015.269 (2016).

48. Wakefield, J. A Bayesian measure of the probability of false discovery in genetic epidemiology studies. Am J Hum Genet 81, 208–227, doi:10.1086/519024 (2007).

49. Johnson, J. L. & Abecasis, G. R. Genetic Association Study (GAS) Power Calculator, <https://csg.sph.umich.edu/abecasis/gas_power_calculator/reference.html#> (https://csg.sph.umich.edu/abecasis/gas_power_calculator/index.html).

## REFERENCES

1. Mahajan, A. et al. Fine-mapping type 2 diabetes loci to single-variant resolution using high-density imputation and islet-specific epigenome maps. Nat Genet 50, 1505–1513 (2018).

2. Liu, C.T. et al. Trans-ethnic Meta-analysis and Functional Annotation Illuminates the Genetic Architecture of Fasting Glucose and Insulin. Am J Hum Genet 99, 56–75 (2016).

3. Ng, M.C. et al. Meta-analysis of genome-wide association studies in African Americans provides insights into the genetic architecture of type 2 diabetes. PLoS Genet 10, e1004517 (2014).

4. Morris, A.P. et al. Large-scale association analysis provides insights into the genetic architecture and pathophysiology of type 2 diabetes. Nat Genet 44, 981–90 (2012).

5. Scott, R.A. et al. An Expanded Genome-Wide Association Study of Type 2 Diabetes in Europeans. Diabetes 66, 2888–2902 (2017).

6. Finucane, H.K. et al. Partitioning heritability by functional annotation using genome-wide association summary statistics. Nat Genet 47, 1228–35 (2015).

7. Lu, Q. et al. Systematic tissue-specific functional annotation of the human genome highlights immune-related DNA elements for late-onset Alzheimer’s disease. PLoS Genet 13, e1006933 (2017).

8. Lu, Q., Powles, R.L., Wang, Q., He, B.J. & Zhao, H. Integrative Tissue-Specific Functional Annotations in the Human Genome Provide Novel Insights on Many Complex Traits and Improve Signal Prioritization in Genome Wide Association Studies. PLoS Genet 12, e1005947 (2016).

9. Yang, J. et al. Genetic variance estimation with imputed variants finds negligible missing heritability for human height and body mass index. Nat Genet 47, 1114–20 (2015).

10. Evans, L.M. et al. Comparison of methods that use whole genome data to estimate the heritability and genetic architecture of complex traits. Nat Genet 50, 737–745 (2018).

11. Varshney, A. et al. Genetic regulatory signatures underlying islet gene expression and type 2 diabetes. Proc Natl Acad Sci U S A 114, 2301–2306 (2017).

12. Kheradpour, P. & Kellis, M. Systematic discovery and characterization of regulatory motifs in ENCODE TF binding experiments. Nucleic Acids Res 42, 2976–87 (2014).

13. Diabetes Epigenome Atlas. https://www.t2depigenome.org/ (Accessed September 11, 2018).

14. Lawlor, N. et al. Single-cell transcriptomes identify human islet cell signatures and reveal cell-type-specific expression changes in type 2 diabetes. Genome Res 27, 208–222 (2017).

15. Fadista, J. et al. Global genomic and transcriptomic analysis of human pancreatic islets reveals novel genes influencing glucose metabolism. Proc Natl Acad Sci U S A 111, 13924–9 (2014).

16. Sarnowski, C. et al. Impact of Rare and Common Genetic Variants on Diabetes Diagnosis by Hemoglobin A1c in Multi-Ancestry Cohorts: The Trans-Omics for Precision Medicine Program. Am J Hum Genet 105, 706–718 (2019).

