## Supplemental Figures for "Rare Non-coding Variation Identified by Large Scale Whole Genome Sequencing Reveals Unexplained Heritability of Type 2 Diabetes": SUPPL.FIGURES.29OCT2020.pdf

### SUPPLEMENTARY FIGURES

**Supplementary Figure 1. Variant frequency spectrum from whole genome sequencing for (A) SNVs and (B) insertions and deletions.** Variants were aggregated by MAC and MAF into six, non-overlapping ranges depicted by bar color. Pancreatic islet-specific, non-exonic, functional annotations and protein-coding annotations were used to further subdivide variant classes. Annotations were broadly grouped into three types. *All genes* annotations relate to variants falling within protein coding, exonic regions, partitioned by predicted effect on protein function. '*Islet regulation and expression'* annotations describe genes expressed within pancreatic islet tissue and active regulatory regions within pancreatic islets. '*Islet interaction and chromatin structure'* annotations relate to active regulatory regions participating in 3D chromatin interactions within islet cells.

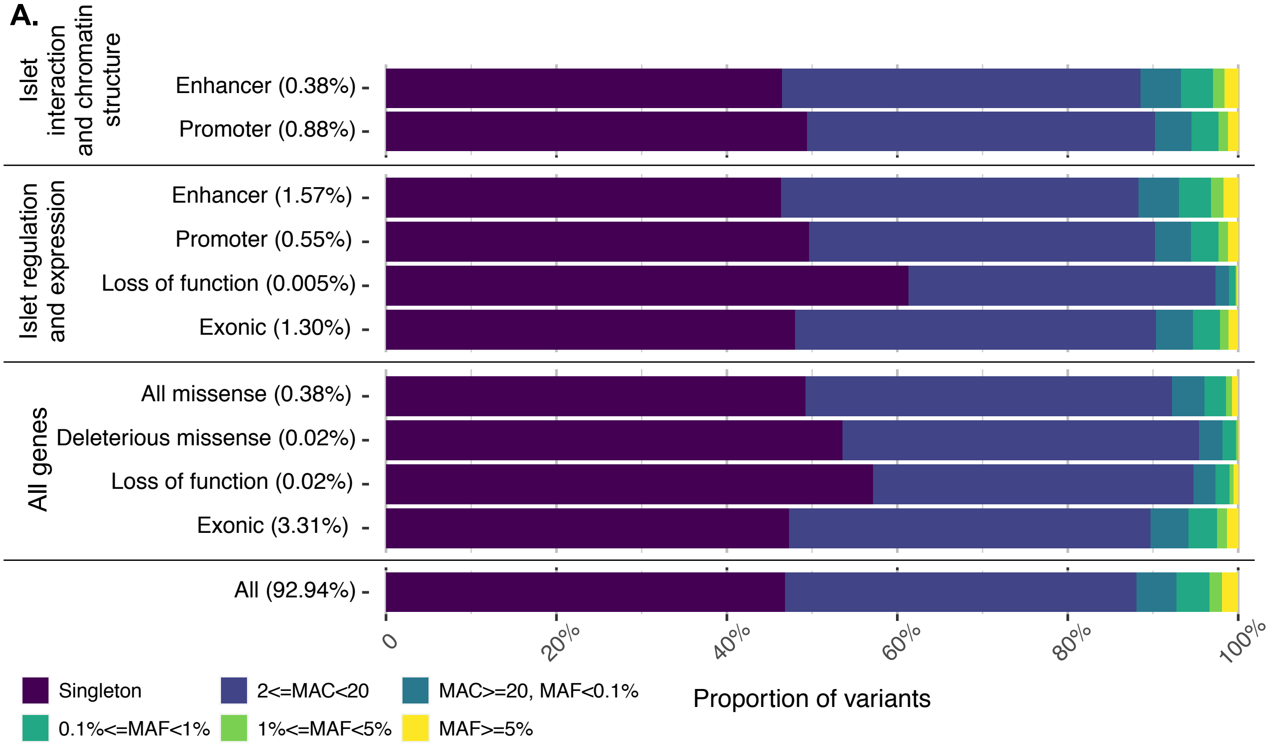

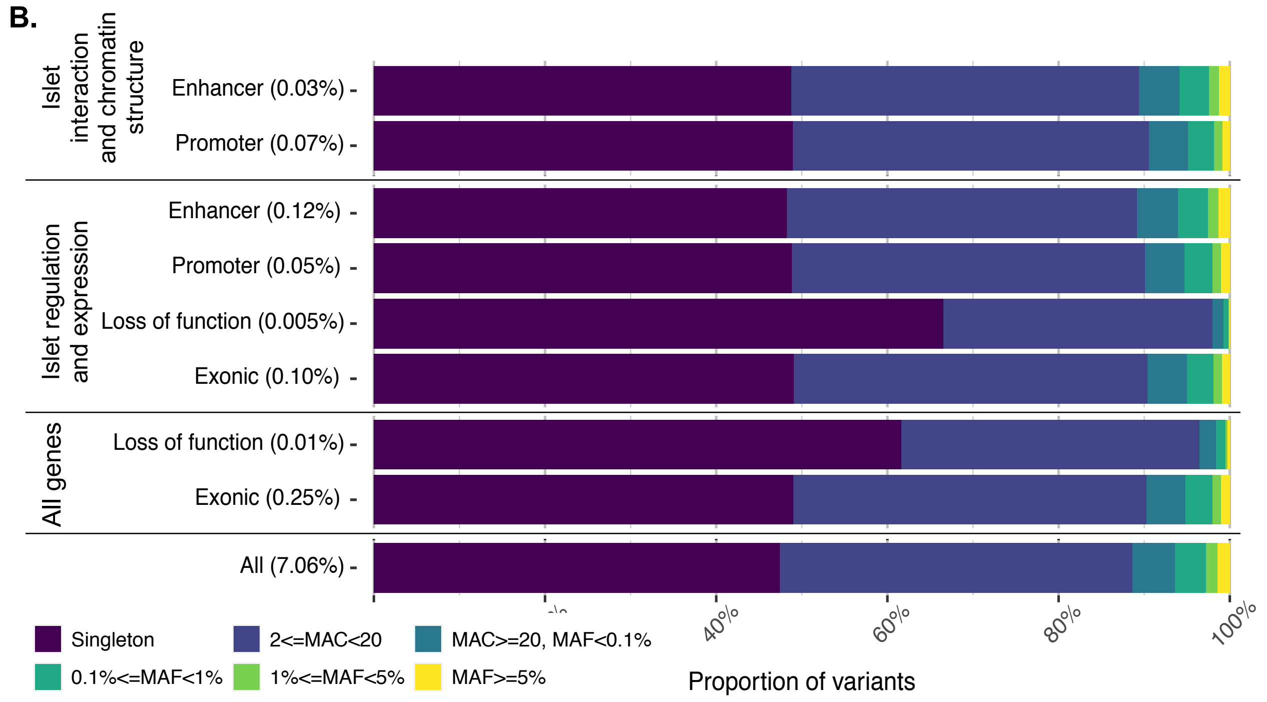

**Supplementary Figure 2. Distribution of LD Scores and minor allele frequency of WGS variants observed in the subset of unrelated individuals in the PC-derived European ancestry sample cluster.** In total, 36,915,823 variants had a minor allele count greater than 5 in the subset of 21,406 individuals from 10 cohorts. Of these, 22,831,695 variants had 0.0001<=MAF<0.001 (mean LD Score = 40.78; SD = 79.17, Median LD Score = 16.98), 5,583,774 variants had 0.001<=MAF<0.01 (mean LD Score = 52.74; SD = 84.22, Median LD Score = 26.69), 2,558,934 had 0.01<=MAF<0.05 (mean LD Score = 96.23; SD = 121.22, Median LD Score = 60.7737), and 5,941,420 variants had 0.05<=MAF<=0.5 (mean LD Score = 181.53; SD = 260.37, Median LD Score = 126.21). In subsequent heritability analyses, individuals from the Amish cohort and the WHI cohort were removed from the sample subset, n=15,109.

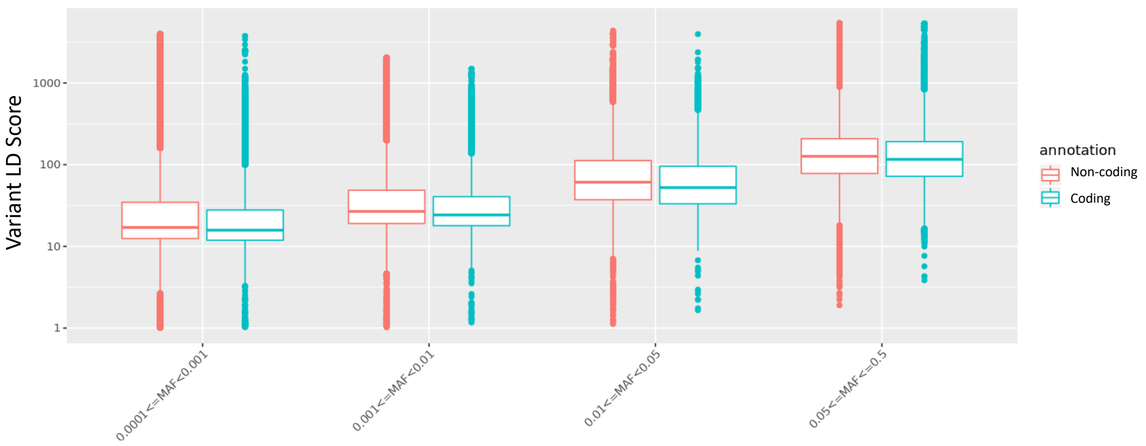

**Supplementary Figure 3. Aggregate variant association results for seven aggregation types: (a,b) Islet regulation and expression (+TFBS), (c,d) Islet regulation and expression, (e,f) Whole hub, (g,h) Hub-components, (i,j) Loss of function, (k,l) Deleterious missense, and (m,n) All missense.** For each aggregate type, Manhattan-style and quantile-quantile plots are shown per ancestry/population-specific and meta-analysis. Each plot combines results from four models: Burden test, adjusted for BMI (purple); Burden test, unadjusted for BMI (blue); SKAT, adjusted for BMI (green); SKAT, unadjusted for BMI (yellow). Bonferroni corrected significance thresholds are represented by a dashed black line in the Manhattan-style plots. Significance thresholds were defined per ancestry by dividing 0.05 by the product of the number of tests and the number of models. Genomic inflation factors are shown for each model in each quantile-quantile plot at lower right.

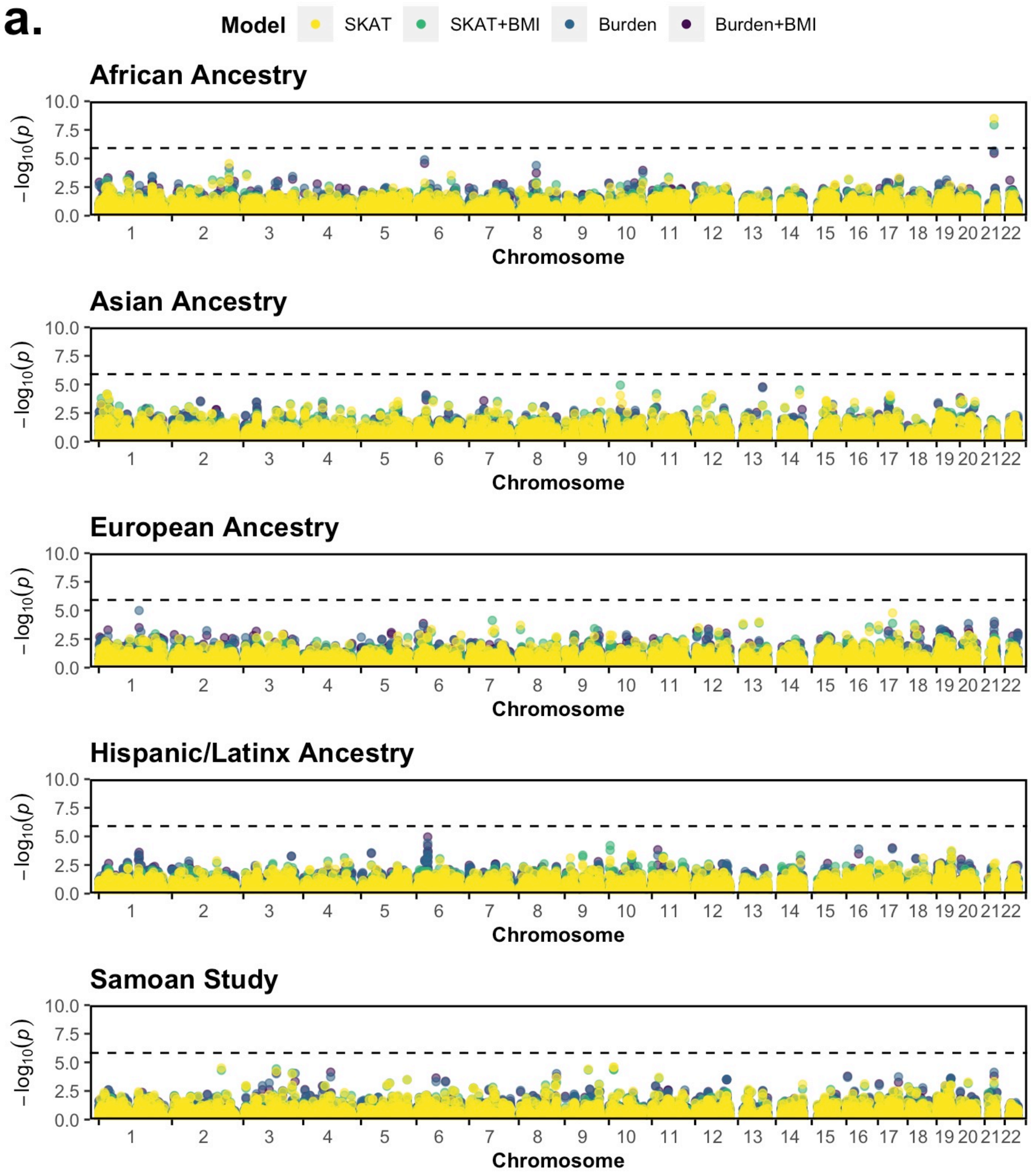
a.
Model
SKAT
SKAT+BMI
Burden
Burden+BMI
African Ancestry
Asian Ancestry
European Ancestry
Hispanic/Latinx Ancestry
Samoan Study
−log10(p)
0.0
2.5
5.0
7.5
10.0
1 2 3 4 5 6 7 8 9 10 11 12 13 14 15 16 17 18 19 20 21 22
Chromosome

b.

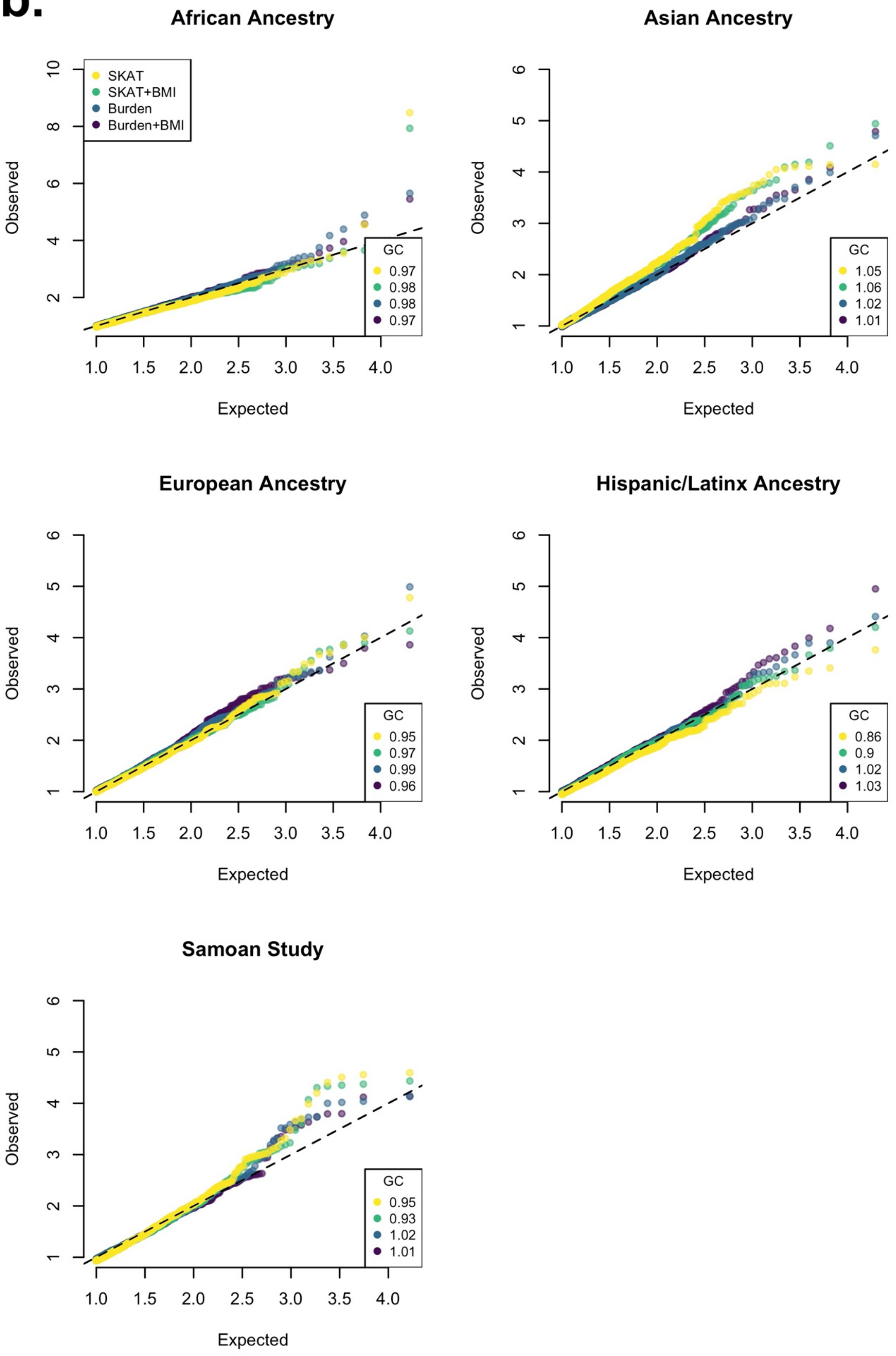

African Ancestry
Asian Ancestry
European Ancestry
Hispanic/Latinx Ancestry
Samoan Study
SKAT
SKAT+BMI
Burden
Burden+BMI
Observed
Expected
GC
0.97
0.98
0.98
0.97
1.05
1.06
1.02
1.01
0.95
0.97
0.99
0.96
0.86
0.9
1.02
1.03
0.95
0.93
1.02
1.01

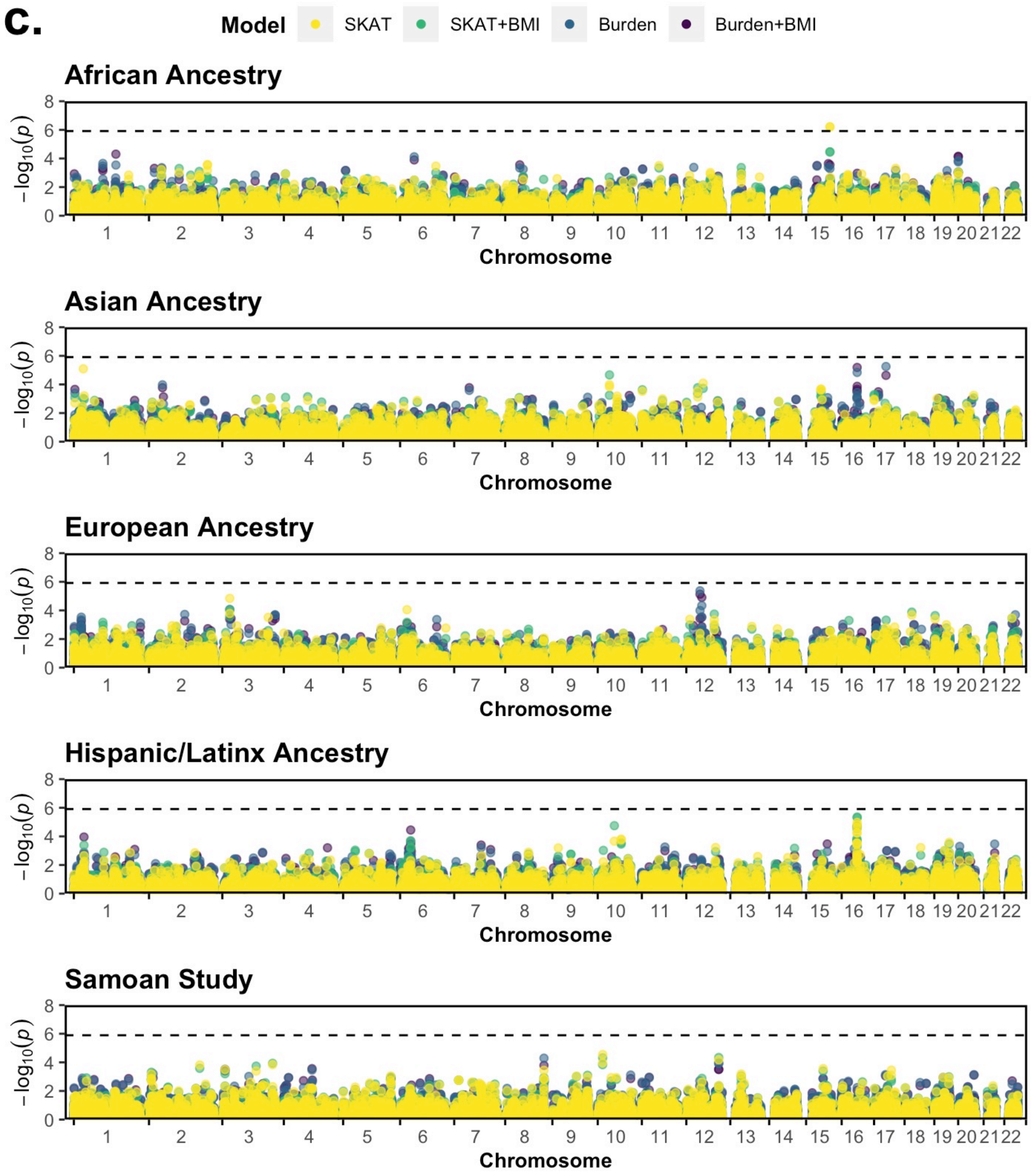
c.
Model
SKAT
SKAT+BMI
Burden
Burden+BMI
African Ancestry
Asian Ancestry
European Ancestry
Hispanic/Latinx Ancestry
Samoan Study
−log10(p)
Chromosome
0
2
4
6
8
1 2 3 4 5 6 7 8 9 10 11 12 13 14 15 16 17 18 19 20 21 22

d.

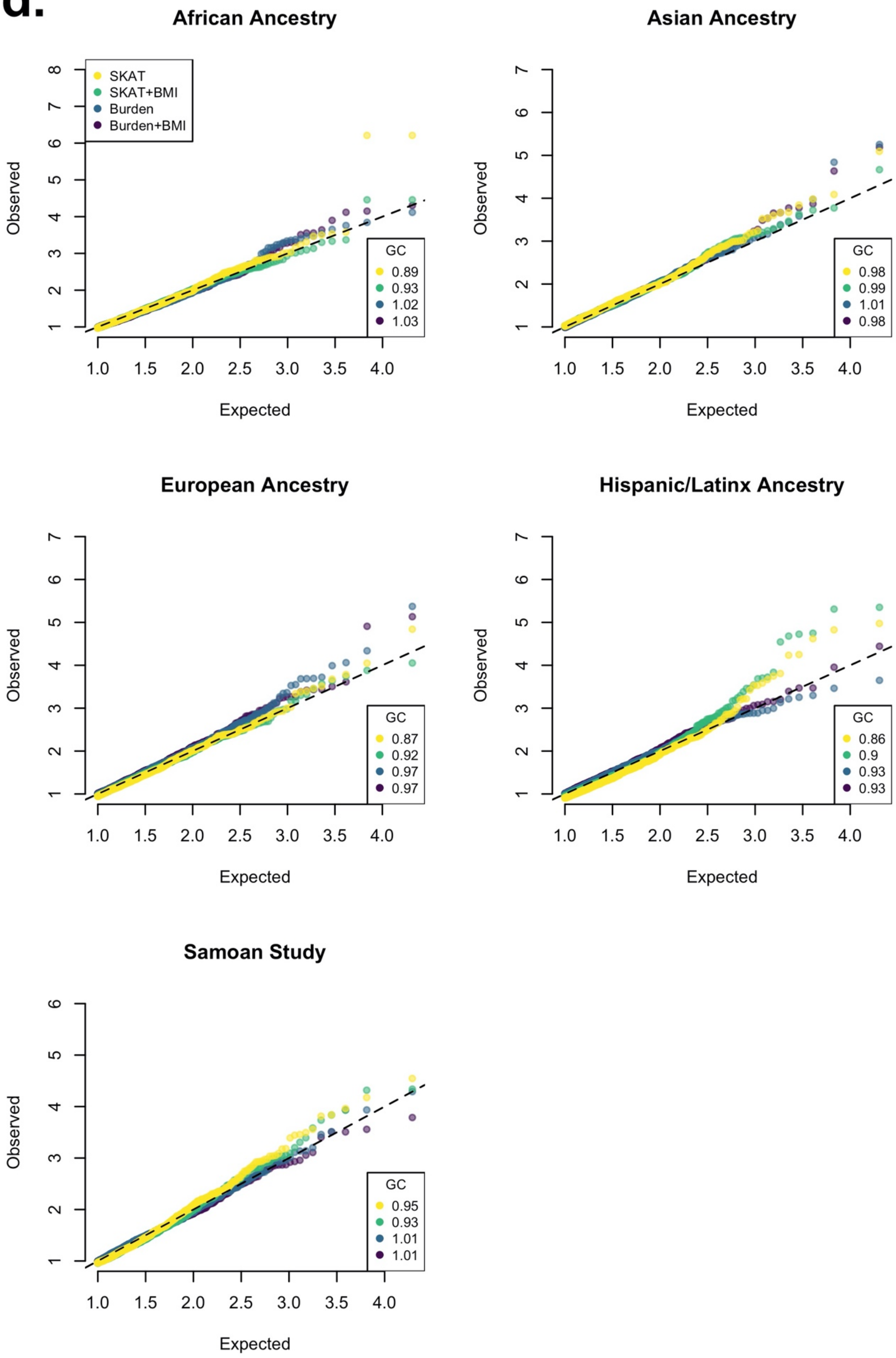
African Ancestry
SKAT
SKAT+BMI
Burden
Burden+BMI
GC
0.89
0.93
1.02
1.03
Asian Ancestry
GC
0.98
0.99
1.01
0.98
European Ancestry
GC
0.87
0.92
0.97
0.97
Hispanic/Latinx Ancestry
GC
0.86
0.9
0.93
0.93
Samoan Study
GC
0.95
0.93
1.01
1.01
Observed
Expected

e.

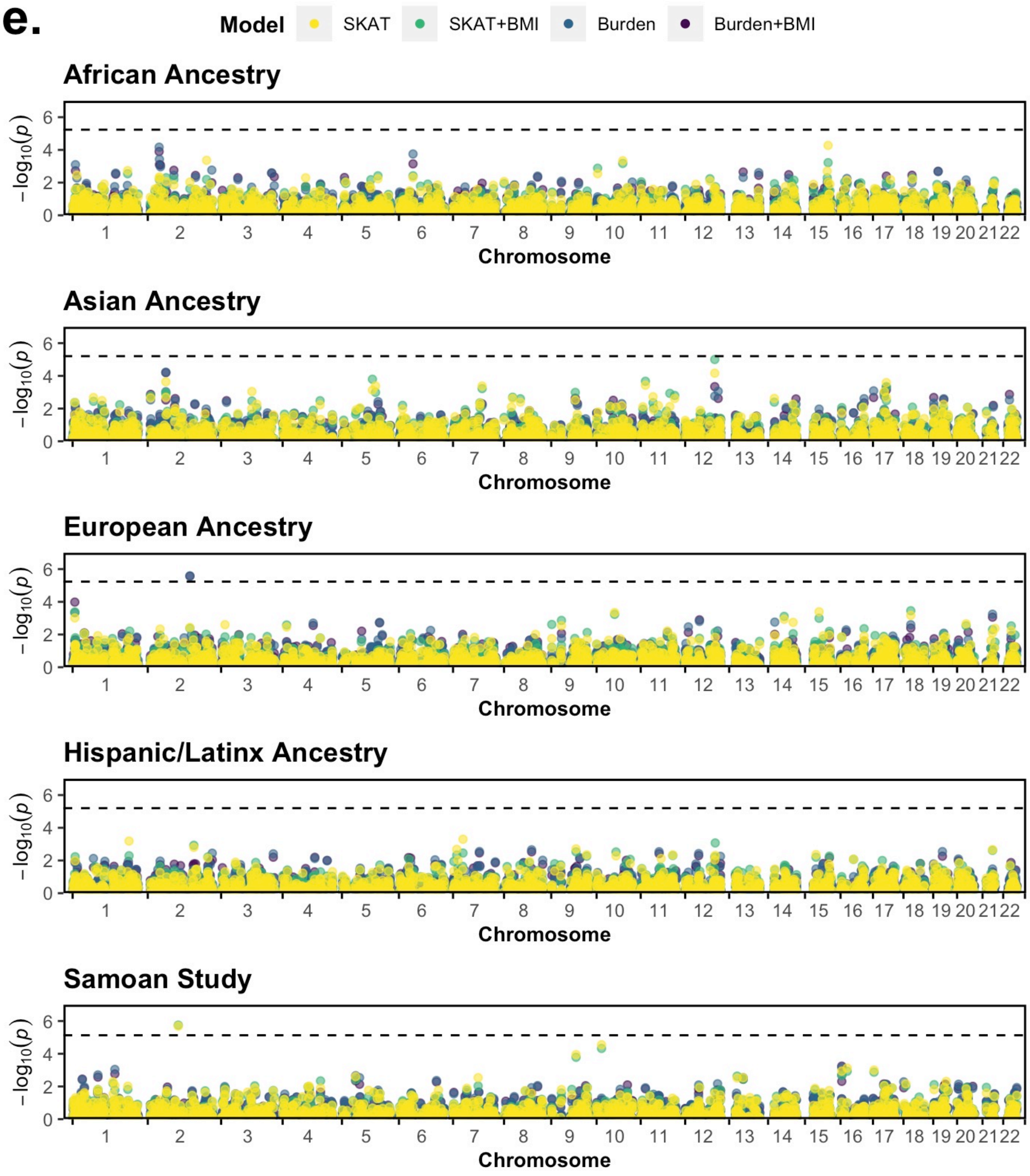

Model
SKAT
SKAT+BMI
Burden
Burden+BMI
African Ancestry
Asian Ancestry
European Ancestry
Hispanic/Latinx Ancestry
Samoan Study
−log10(p)
Chromosome

f.

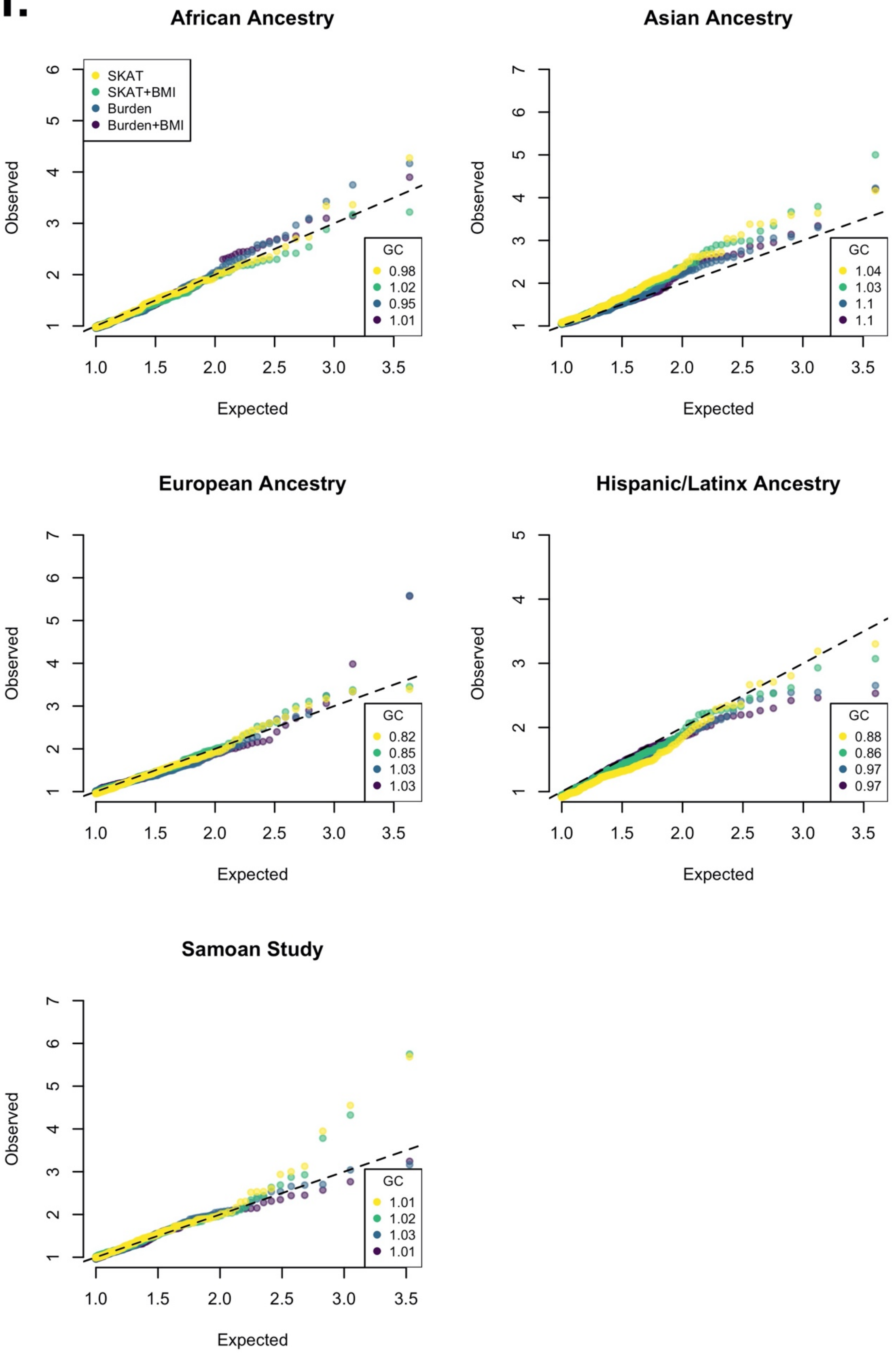
African Ancestry
SKAT
SKAT+BMI
Burden
Burden+BMI
GC
0.98
1.02
0.95
1.01
Observed
Expected
Asian Ancestry
GC
1.04
1.03
1.1
1.1
Observed
Expected
European Ancestry
GC
0.82
0.85
1.03
1.03
Observed
Expected
Hispanic/Latinx Ancestry
GC
0.88
0.86
0.97
0.97
Observed
Expected
Samoan Study
GC
1.01
1.02
1.03
1.01
Observed
Expected

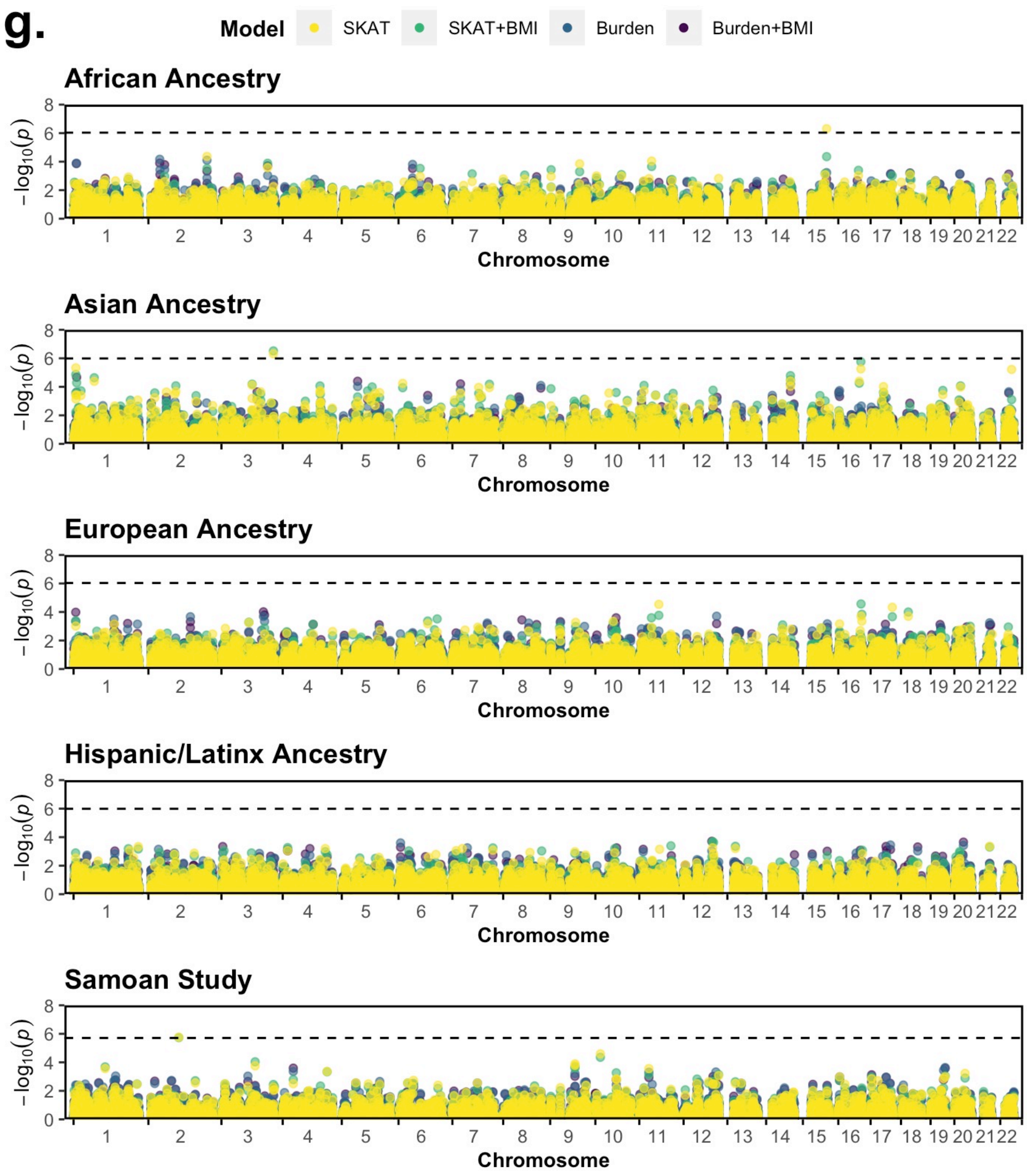
g.
Model
SKAT
SKAT+BMI
Burden
Burden+BMI
African Ancestry
Asian Ancestry
European Ancestry
Hispanic/Latinx Ancestry
Samoan Study
$-\log_{10}(p)$
Chromosome
1 2 3 4 5 6 7 8 9 10 11 12 13 14 15 16 17 18 19 20 21 22

**h.**

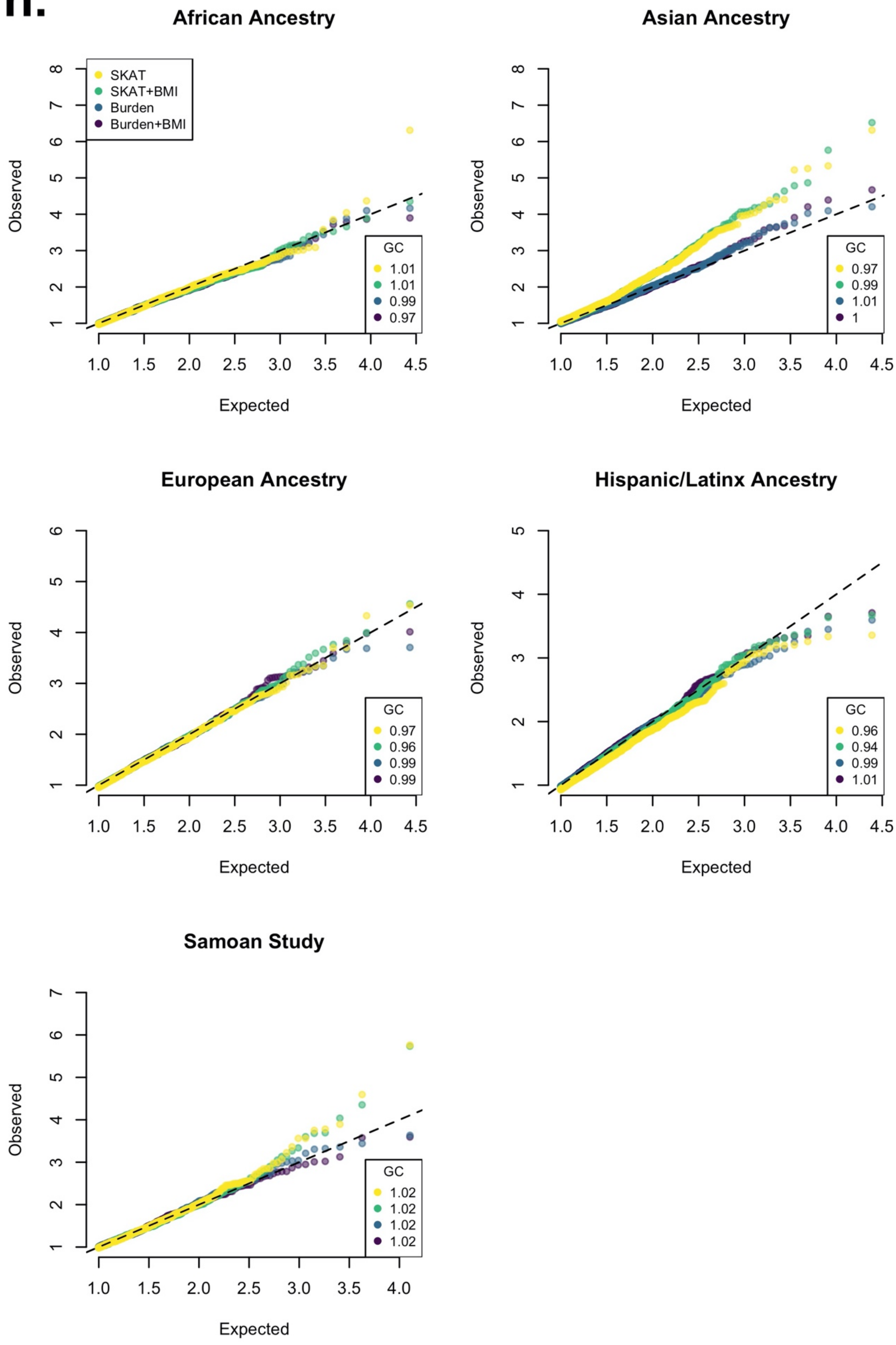
African Ancestry
SKAT
SKAT+BMI
Burden
Burden+BMI
GC
1.01
1.01
0.99
0.97
Observed
Expected
Asian Ancestry
GC
0.97
0.99
1.01
1
Observed
Expected
European Ancestry
GC
0.97
0.96
0.99
0.99
Observed
Expected
Hispanic/Latinx Ancestry
GC
0.96
0.94
0.99
1.01
Observed
Expected
Samoan Study
GC
1.02
1.02
1.02
1.02
Observed
Expected

i.

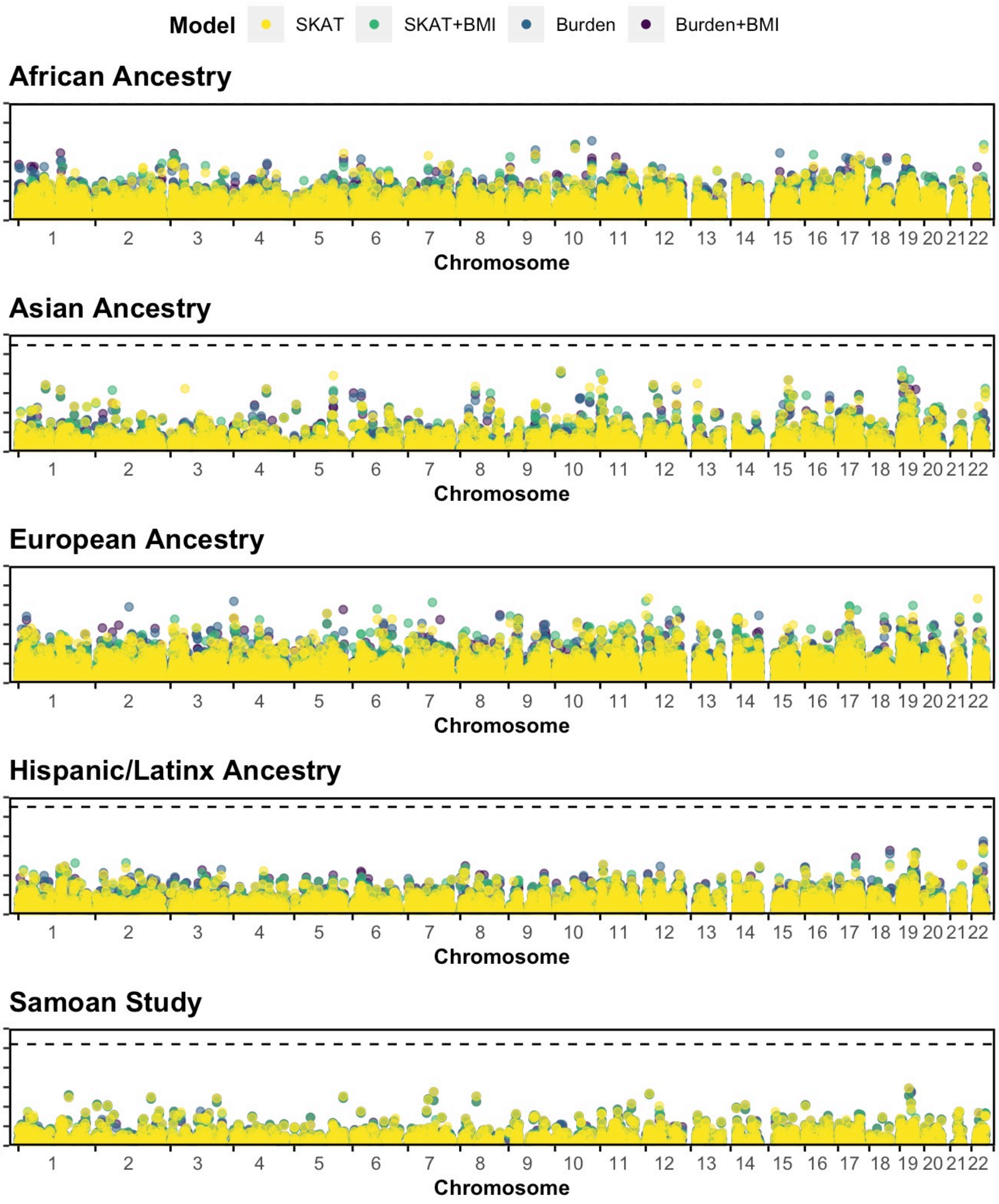
Model
SKAT
SKAT+BMI
Burden
Burden+BMI
African Ancestry
Asian Ancestry
European Ancestry
Hispanic/Latinx Ancestry
Samoan Study
$-\log_{10}(p)$
Chromosome
1 2 3 4 5 6 7 8 9 10 11 12 13 14 15 16 17 18 19 20 21 22

j.

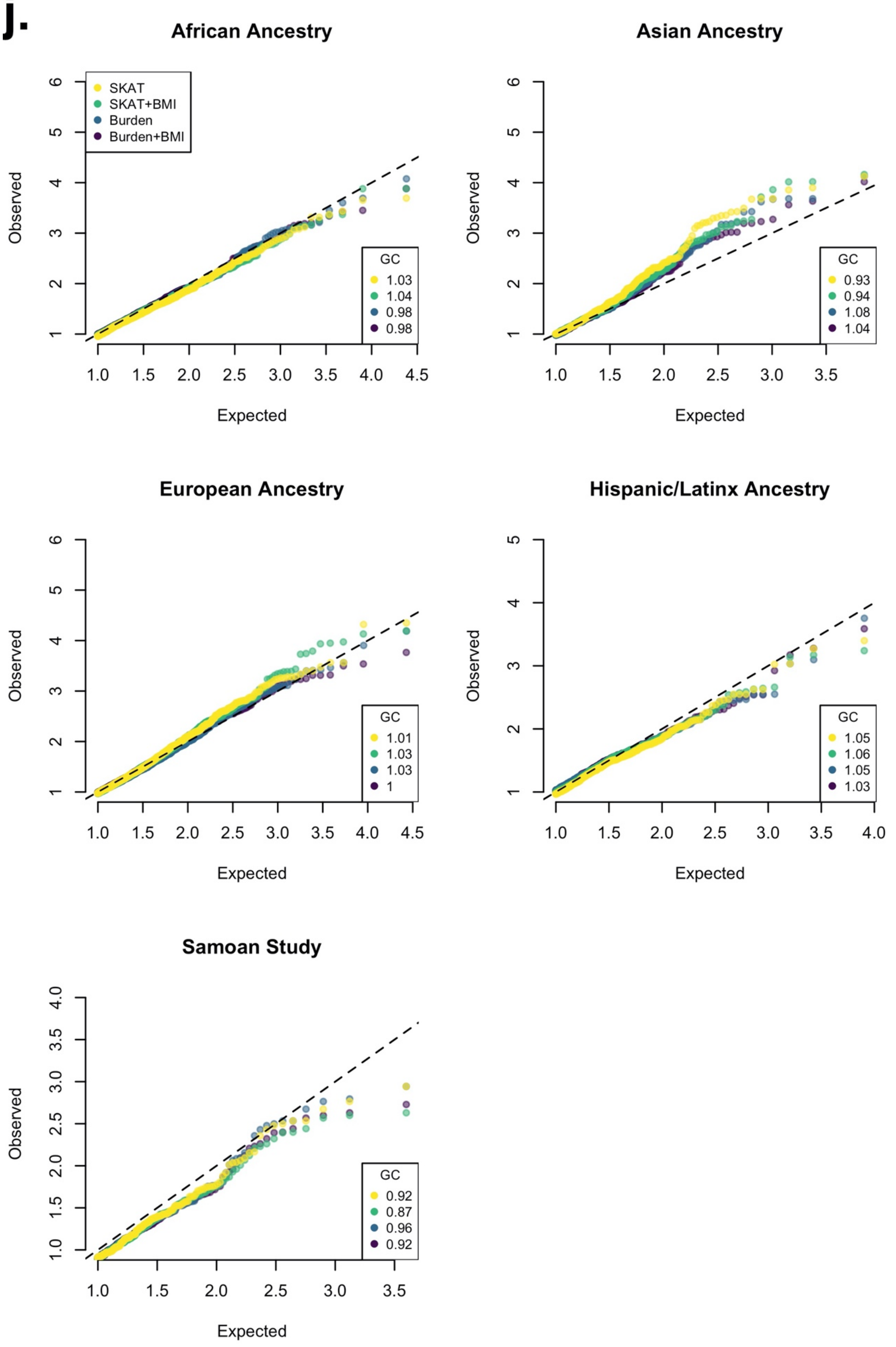
African Ancestry
SKAT
SKAT+BMI
Burden
Burden+BMI
GC
1.03
1.04
0.98
0.98
Observed
Expected
Asian Ancestry
GC
0.93
0.94
1.08
1.04
Observed
Expected
European Ancestry
GC
1.01
1.03
1.03
1
Observed
Expected
Hispanic/Latinx Ancestry
GC
1.05
1.06
1.05
1.03
Observed
Expected
Samoan Study
GC
0.92
0.87
0.96
0.92
Observed
Expected

**k.**

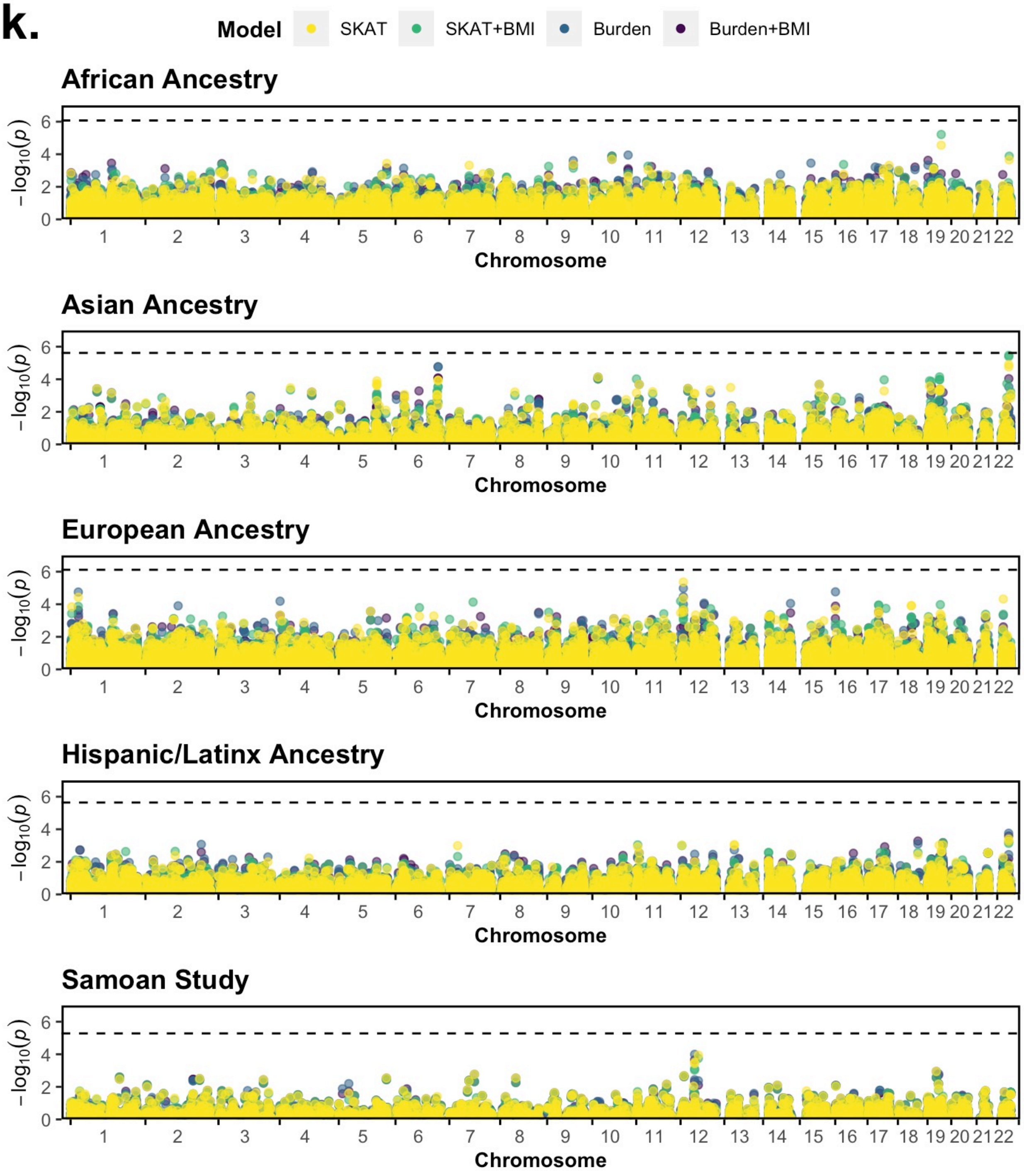

Model
SKAT
SKAT+BMI
Burden
Burden+BMI
African Ancestry
Asian Ancestry
European Ancestry
Hispanic/Latinx Ancestry
Samoan Study
−log10(p)
0
2
4
6
Chromosome

I.

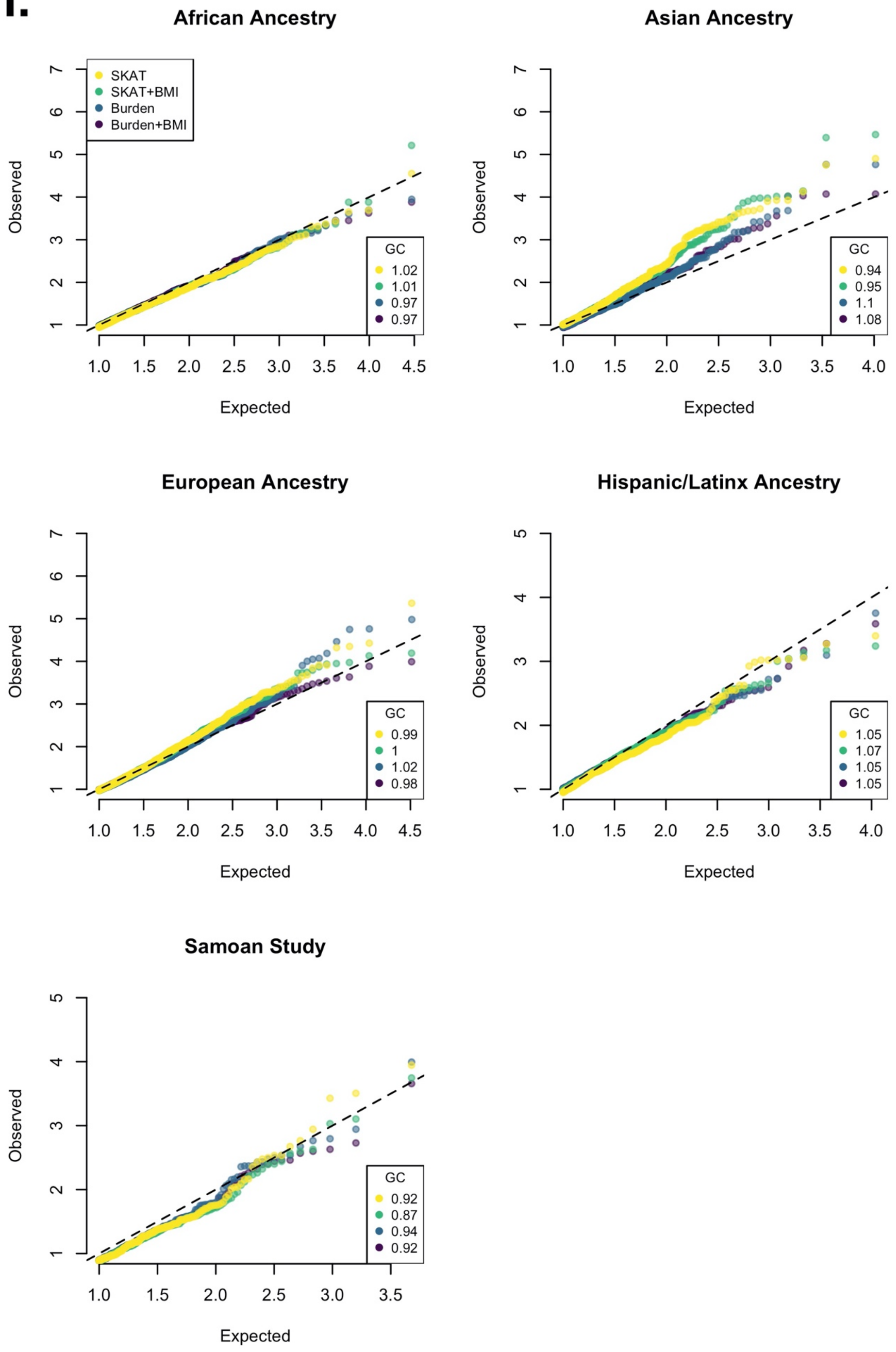
African Ancestry
Asian Ancestry
European Ancestry
Hispanic/Latinx Ancestry
Samoan Study
SKAT
SKAT+BMI
Burden
Burden+BMI
GC
1.02
1.01
0.97
0.97
GC
0.94
0.95
1.1
1.08
GC
0.99
1
1.02
0.98
GC
1.05
1.07
1.05
1.05
GC
0.92
0.87
0.94
0.92
Observed
Expected

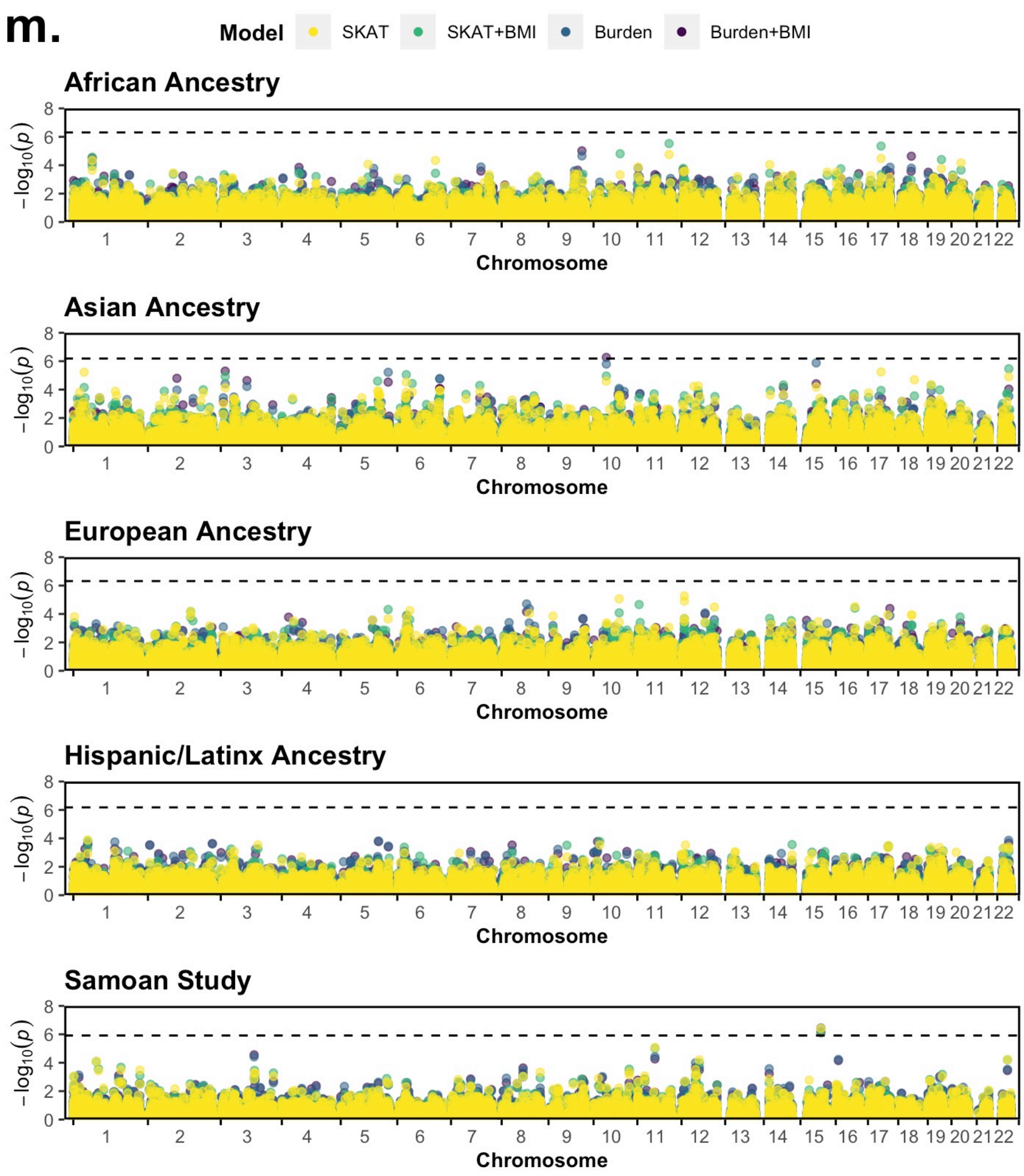

m.
Model
SKAT
SKAT+BMI
Burden
Burden+BMI
African Ancestry
Asian Ancestry
European Ancestry
Hispanic/Latinx Ancestry
Samoan Study
$-\log_{10}(p)$
0
2
4
6
8
Chromosome

n.

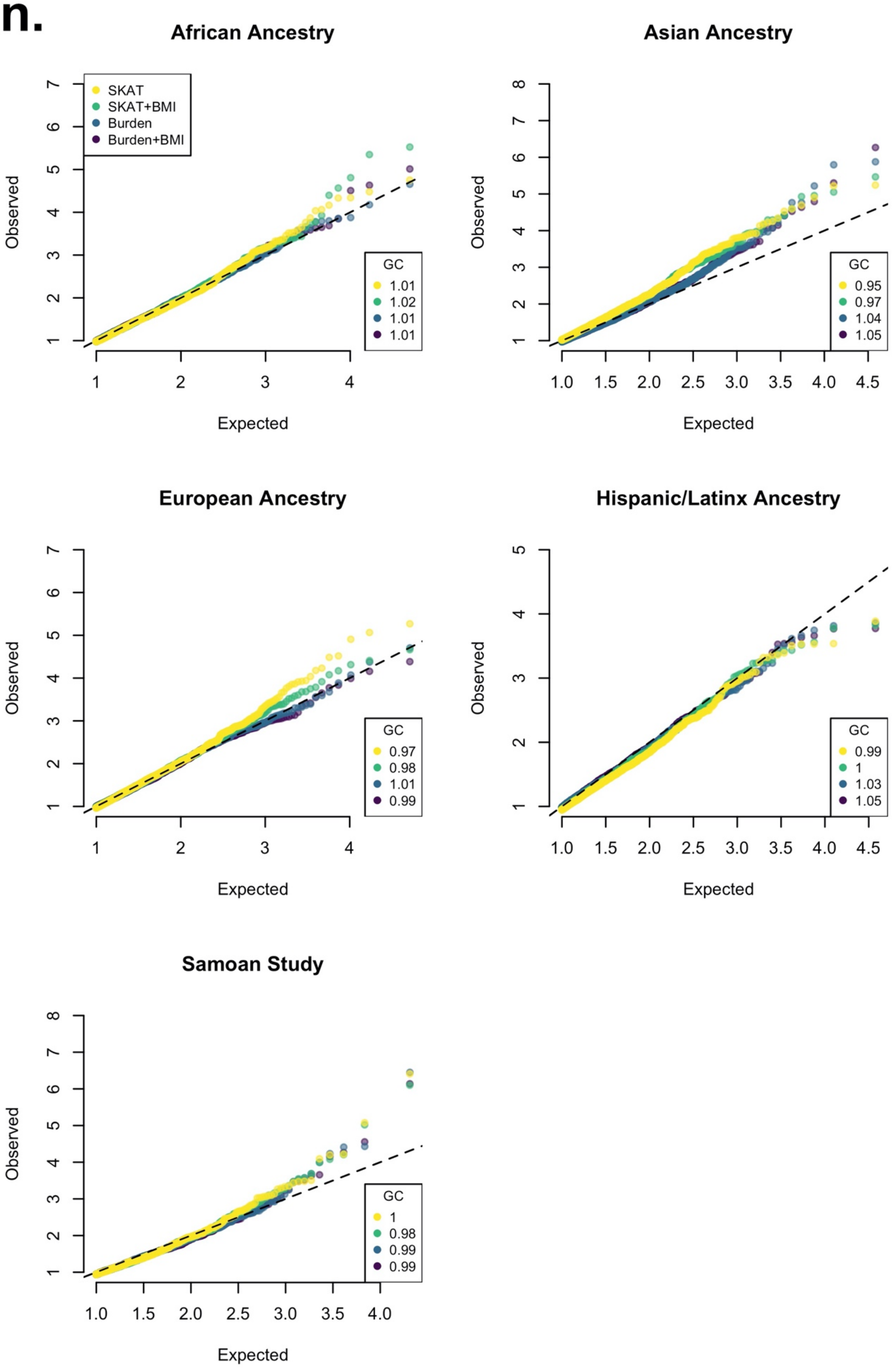
African Ancestry
SKAT
SKAT+BMI
Burden
Burden+BMI
GC
1.01
1.02
1.01
1.01
Observed
Expected
Asian Ancestry
GC
0.95
0.97
1.04
1.05
Observed
Expected
European Ancestry
GC
0.97
0.98
1.01
0.99
Observed
Expected
Hispanic/Latinx Ancestry
GC
0.99
1
1.03
1.05
Observed
Expected
Samoan Study
GC
1
0.98
0.99
0.99
Observed
Expected

**Supplementary Figure 4. Manhattan, quantile-quantile, and P value-effect size scatter plots for single variant association analyses with the T2D outcome.** For each ancestry or population group, meta-analyses, or pooled analyses, a Manhattan style plot was generated using results from both BMI adjusted (purple) and BMI unadjusted (green) models. Quantile-quantile plots are in the center with genomic inflation for each model at the top left of each plot. Furthest to the right, scatter plots comparing BMI adjusted and BMI unadjusted P values are shown. Points in each plot are colored by the model (BMI adj. or BMI unadj.) in which the smaller P value was observed. The size of each point is proportional to the effect size estimate in the more significant model. Horizontal and vertical dashed lines are placed at $P = 5x10^{-8}$. For some analyses, see the inset for the full distribution of P values and effect sizes as the plotting ranges have been limited to $P < 1x10^{-15}$.

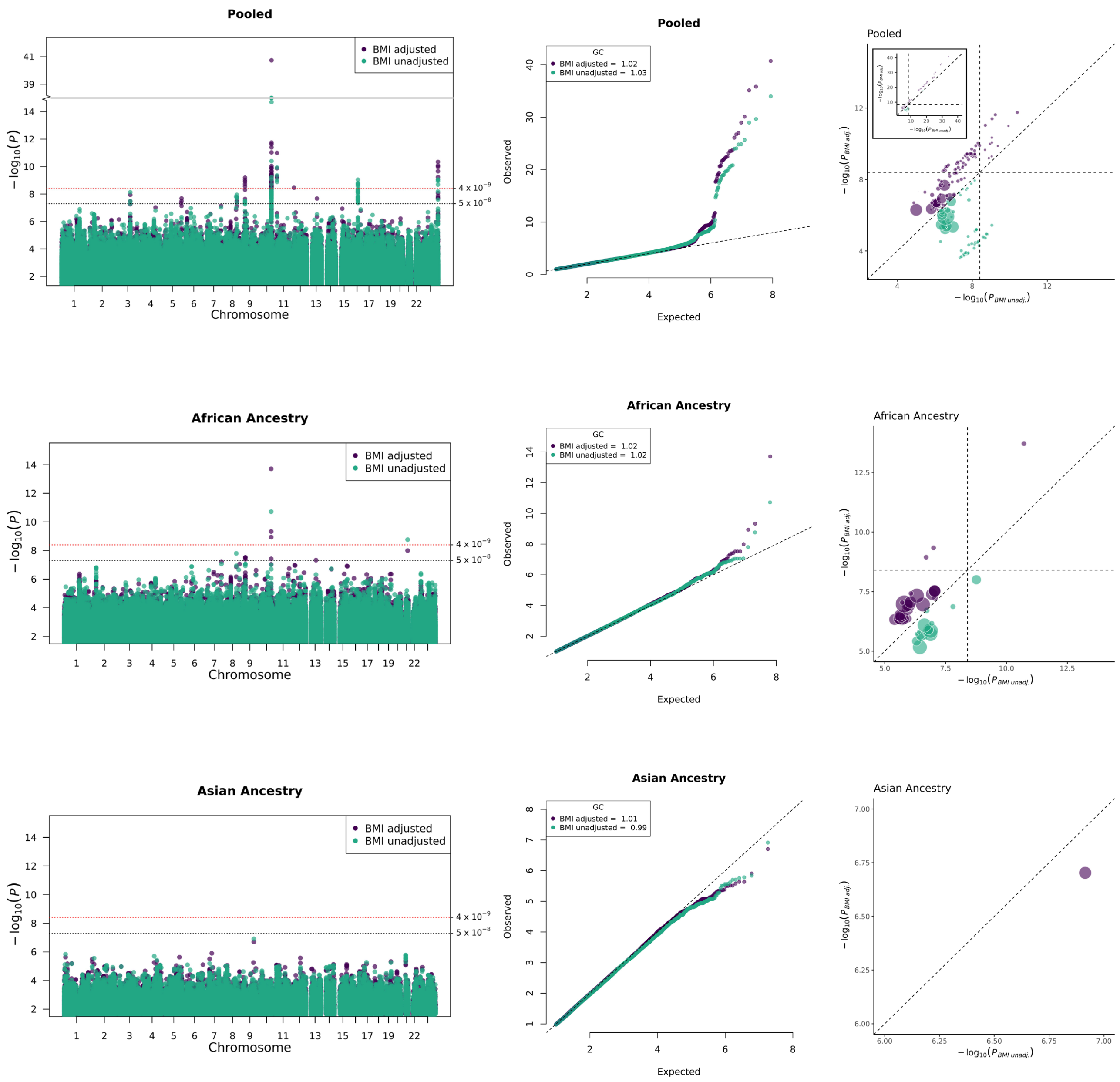
Pooled
BMI adjusted
BMI unadjusted
4 x 10^-9
5 x 10^-8
− log10(P)
Chromosome
GC
BMI adjusted = 1.02
BMI unadjusted = 1.03
Observed
Expected
− log10(P BMI adj.)
− log10(P BMI unadj.)
African Ancestry
BMI adjusted
BMI unadjusted
4 x 10^-9
5 x 10^-8
− log10(P)
Chromosome
GC
BMI adjusted = 1.02
BMI unadjusted = 1.02
Observed
Expected
− log10(P BMI adj.)
− log10(P BMI unadj.)
Asian Ancestry
BMI adjusted
BMI unadjusted
4 x 10^-9
5 x 10^-8
− log10(P)
Chromosome
GC
BMI adjusted = 1.01
BMI unadjusted = 0.99
Observed
Expected
− log10(P BMI adj.)
− log10(P BMI unadj.)

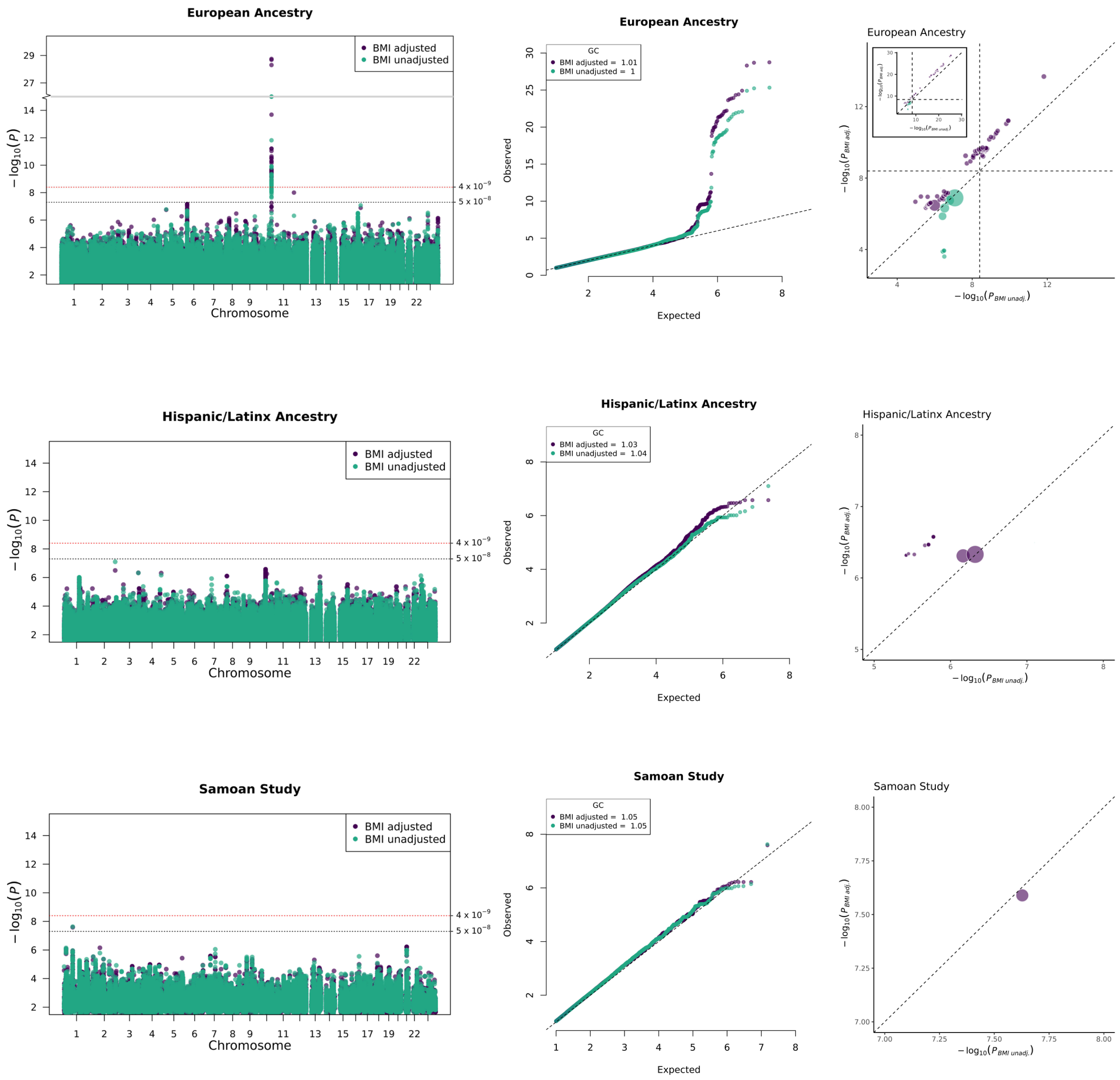
European Ancestry
BMI adjusted
BMI unadjusted
4 x 10^-9
5 x 10^-8
− log10(P)
Chromosome
GC
BMI adjusted = 1.01
BMI unadjusted = 1
Observed
Expected
− log10(P BMI adj.)
− log10(P BMI unadj.)
Hispanic/Latinx Ancestry
BMI adjusted
BMI unadjusted
GC
BMI adjusted = 1.03
BMI unadjusted = 1.04
Samoan Study
BMI adjusted
BMI unadjusted
GC
BMI adjusted = 1.05
BMI unadjusted = 1.05

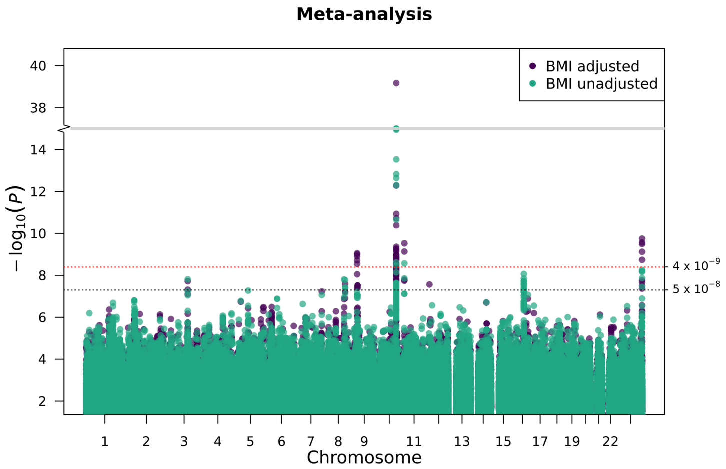
Meta-analysis
BMI adjusted
BMI unadjusted
$-\log_{10}(P)$
4 x 10$^{-9}$
5 x 10$^{-8}$
Chromosome

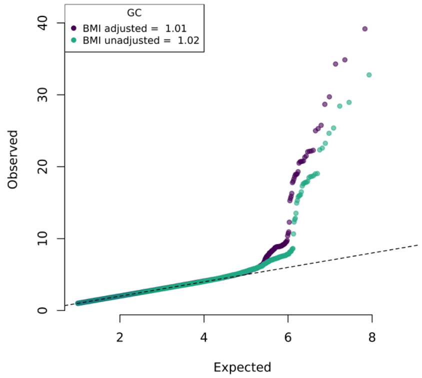
GC
BMI adjusted = 1.01
BMI unadjusted = 1.02
Observed
Expected

**Supplementary Figure 5. Locus zoom plots of regions with secondary signals observed in conditional analyses.** a. CDKN2B-AS1; b. NWD2; c. SLC30A8; d. NFIC; (See **Supplementary Tables 11, 21 and 22**). Plots order: marginal results, condition on most significant (index) variant, condition on secondary signal.

**a. CDKN2B-AS1.** Associated with T2D in pooled analyses, BMIunadj.

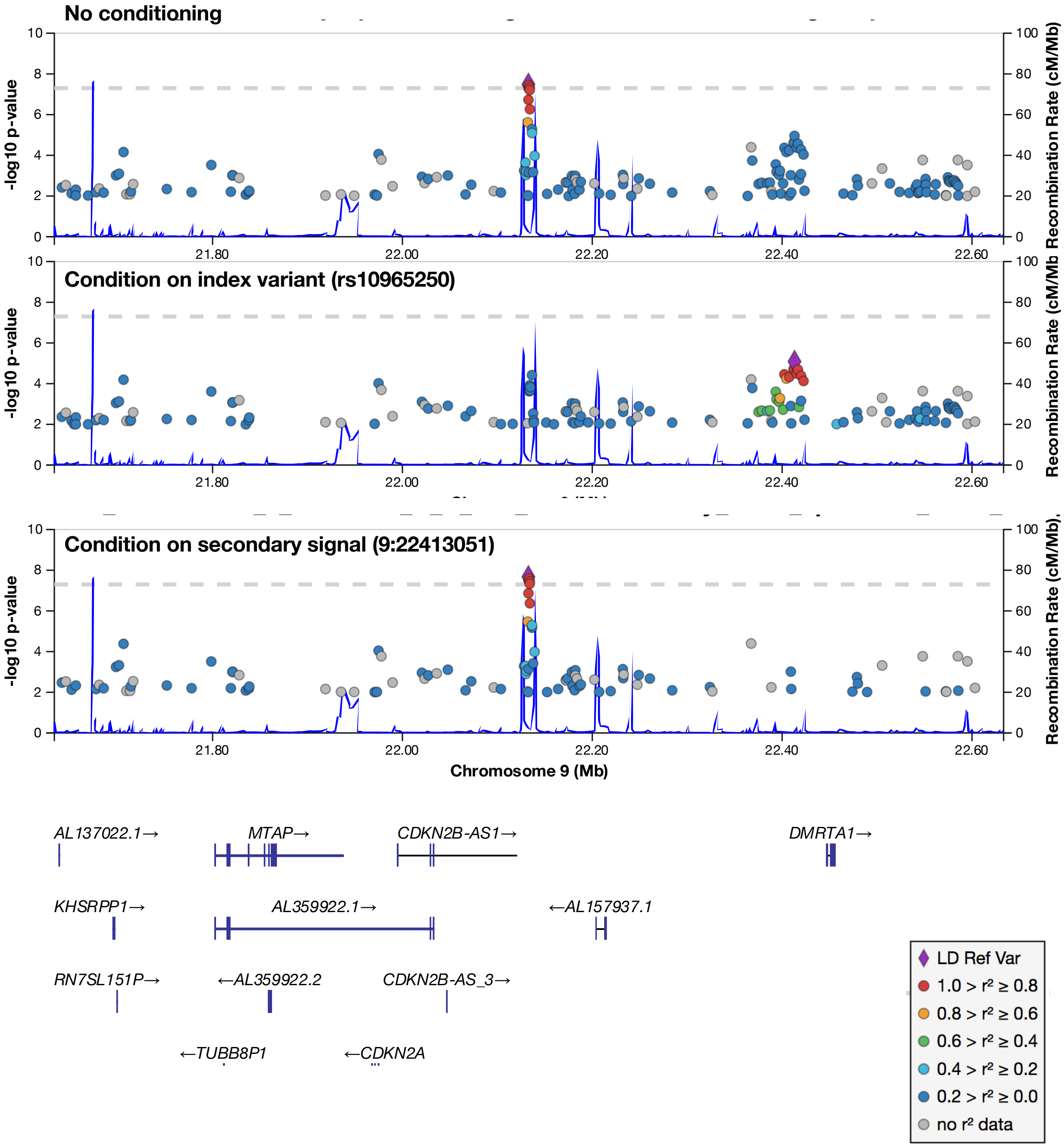

**b. NWD2.** Associated with T2D+ in pooled analyses, BMIunadj.

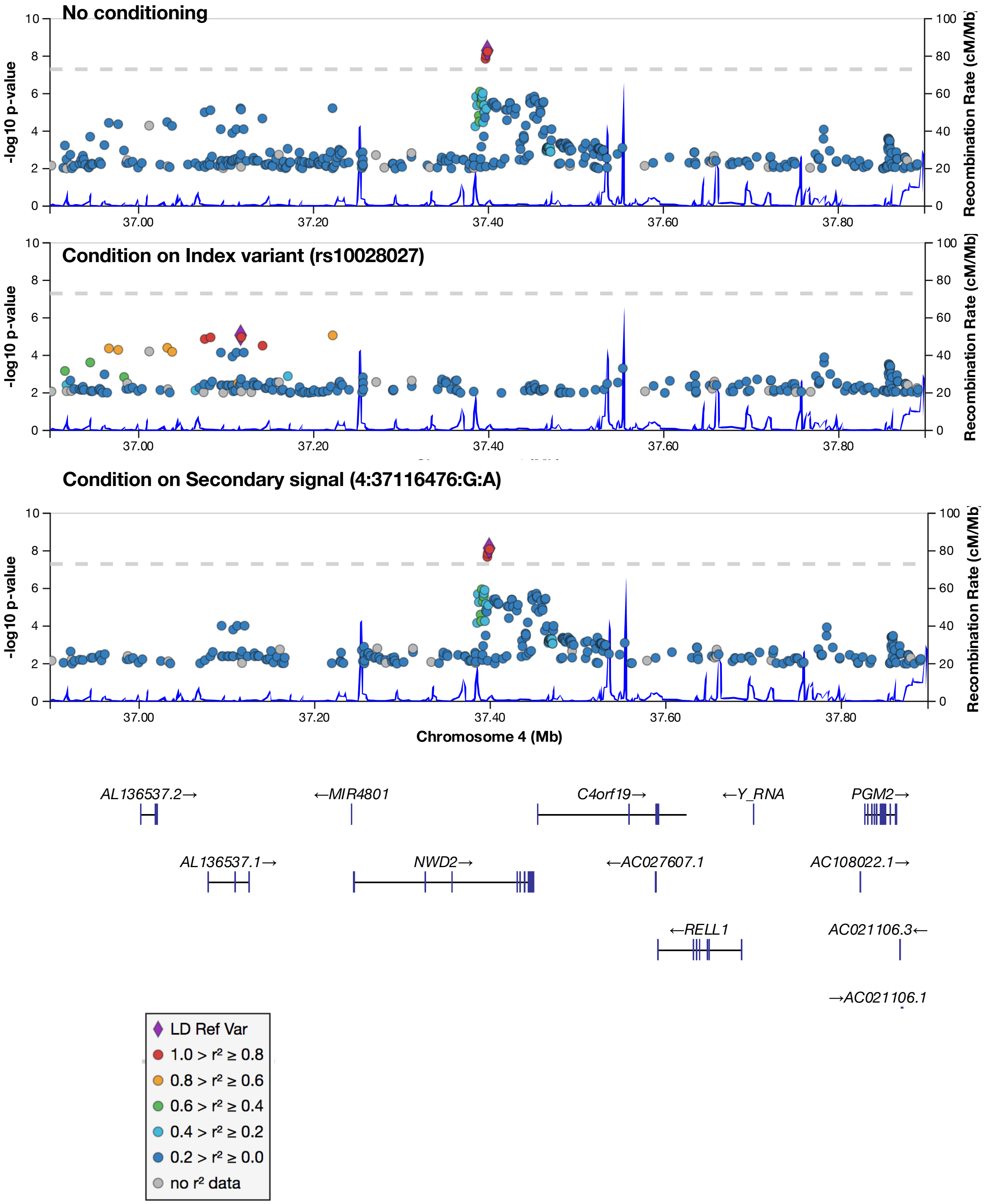

**c. SLC30A8.** Associated with T2D+ in pooled analyses, BMIunadj.

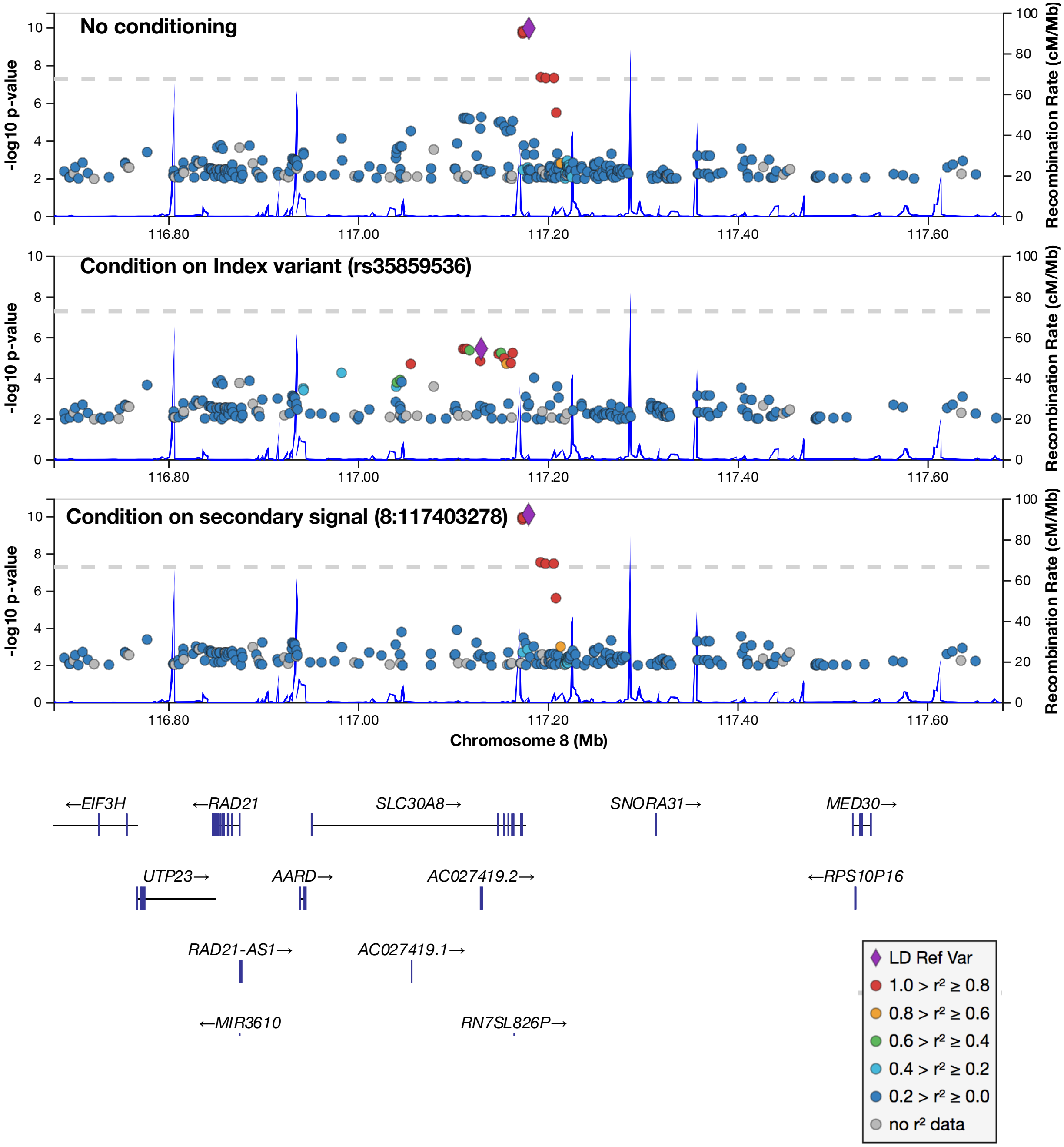

**d. NFIC.** Associated with+ T2D in African ancestry analyses, BMIunadj.

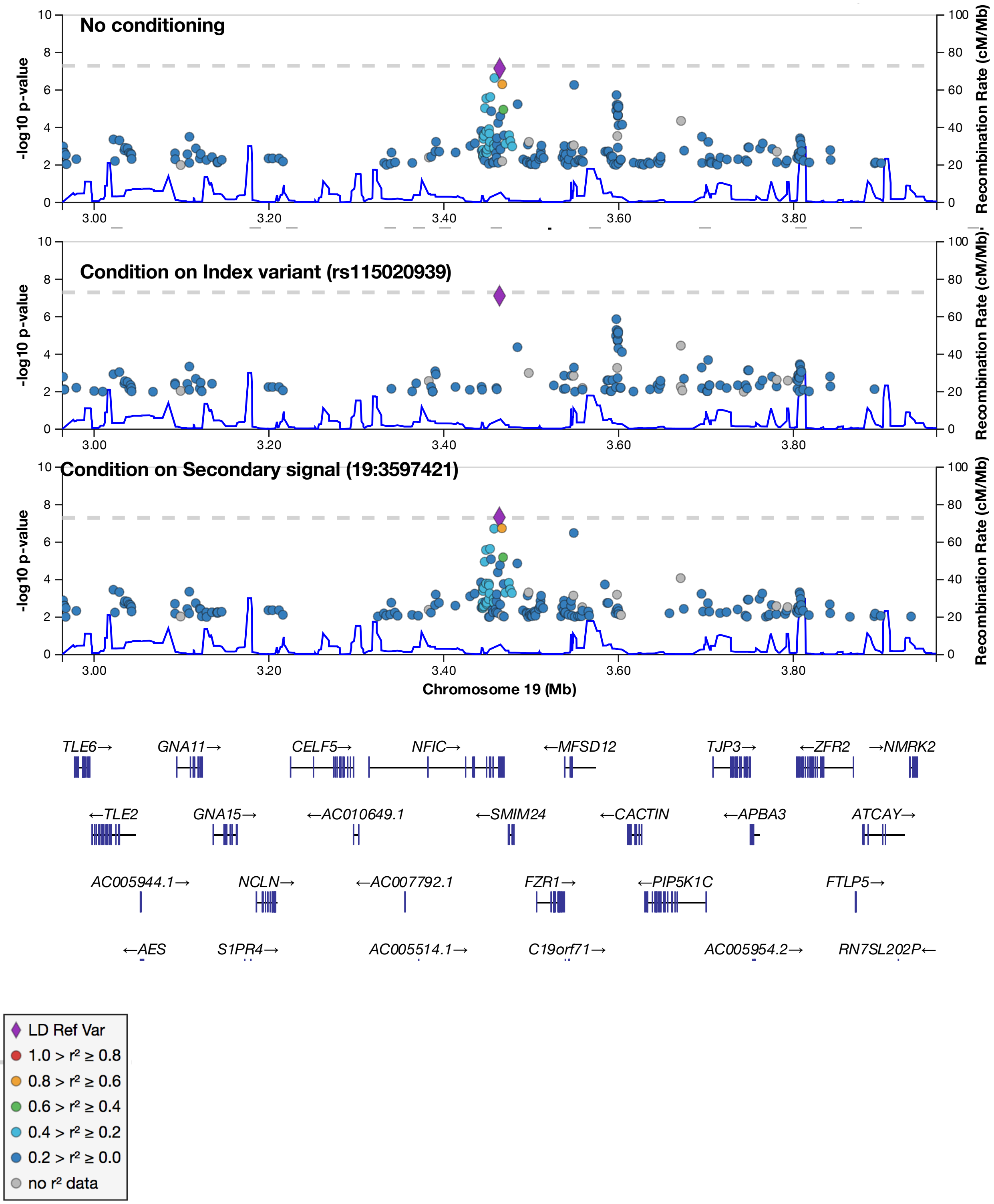

**Supplementary Figure 6. Credible set posterior probability comparison to Mahajan et al.** The posterior probability of variants in overlapping credible sets between the two analyses. Each locus is represented by a different color, with one point for each variant. Open circles indicate no posterior probability was generated for one dataset.

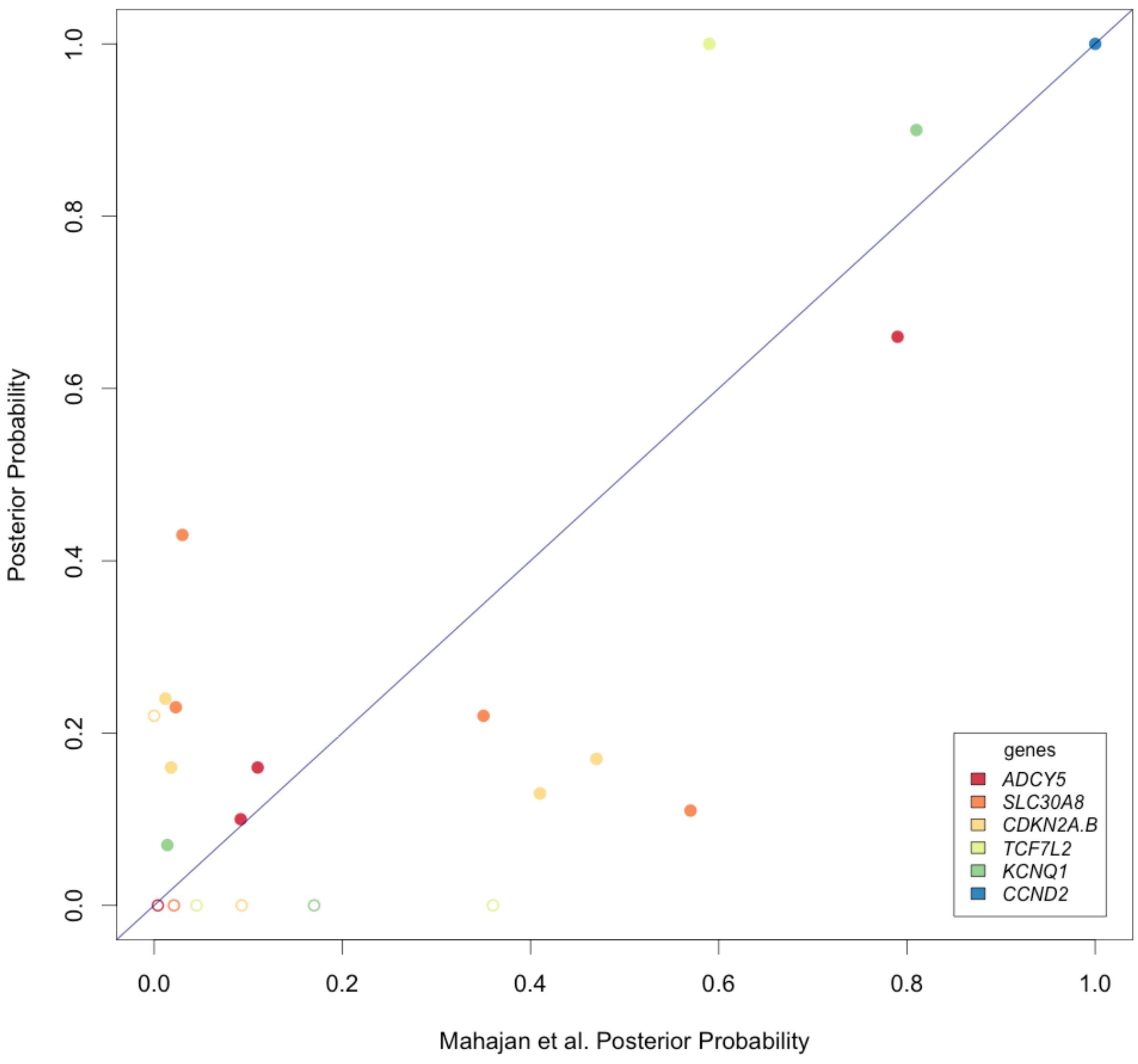

**Supplementary Figure 7. Power to detect an association by odds ratio, at different minor allele frequencies.** Assuming an additive model, T2D prevalence of 8%, and P value<4x$10^{-9}$.

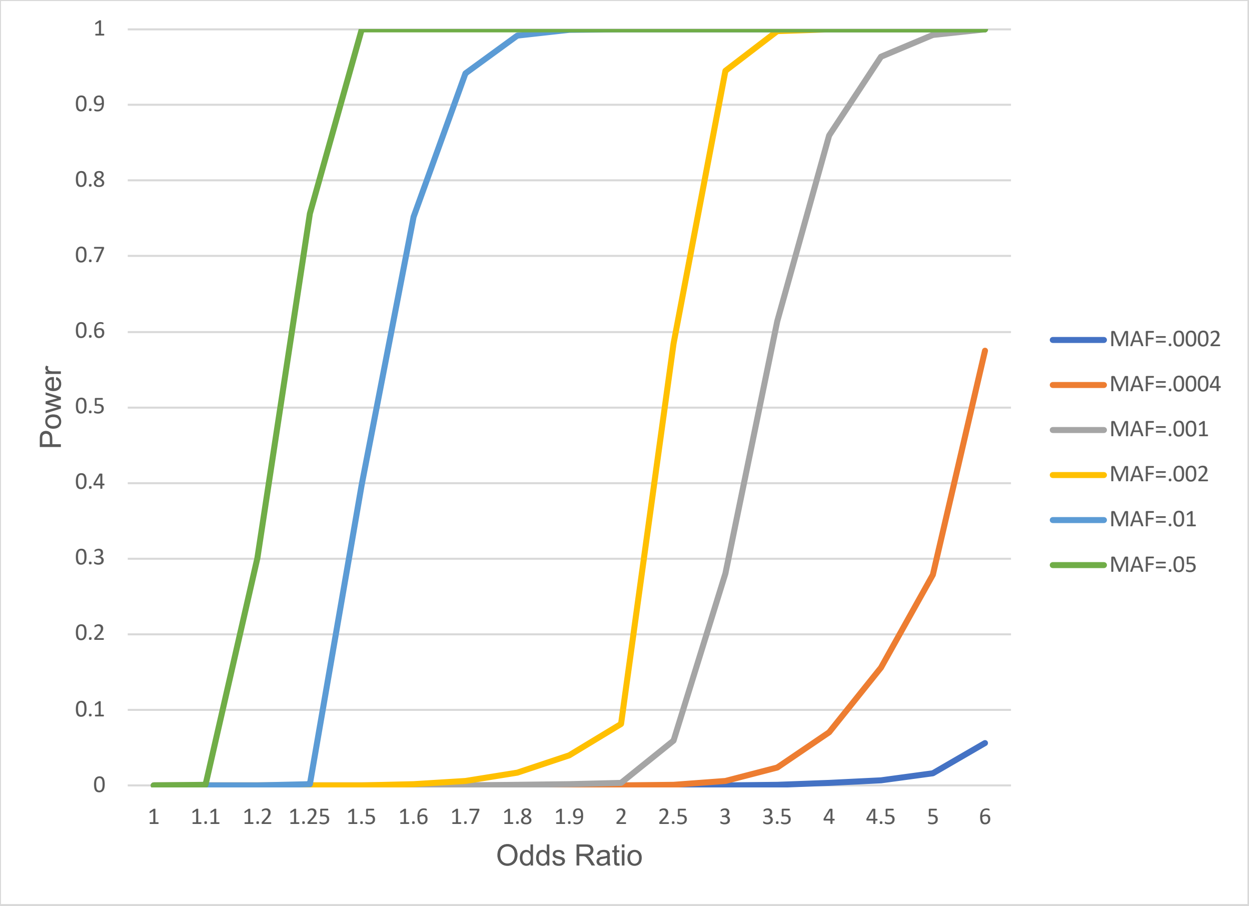

**Supplementary Figure 8:** Quantile-Quantile plots of P values from single variant, pooled analysis. P values for variants within known T2D associated loci are shown in (A). P values for all variants not within T2D associated loci are shown in (B). The genomic inflation factor (lambda GC) was calculated for each set of variants, subset by minor allele frequency, and depicted at top left in each plot. Within known loci, P values were inflated among variants with MAF ≥ 1%. Substantial inflation was not observed within regions not previously implicated in T2D or among variants with MAF < 1%.

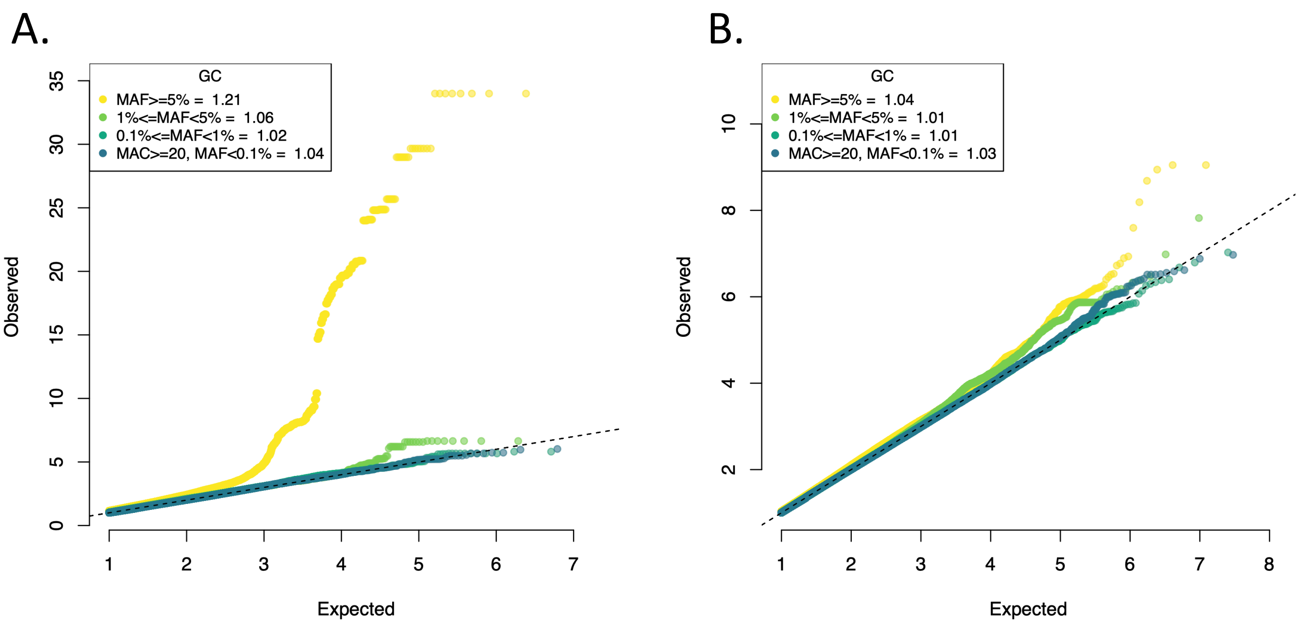

**Supplementary Figure 9. Relationship of effect size with allele frequency among diabetes-associated variants with P value<5x10$^{-5}$.** Odds ratio (OR) of T2D plotted against minor allele frequency for all variants with P value<5x10$^{-5}$ and minor allele count >20. Variants falling within gene boundaries (left) were classified as exonic, missense, missense in genes expressed in islets, or none of the above. All variants (right) were classified as *'Islet interaction and chromatin structure'* or '*Islet regulation and expression'* or neither based on islet annotation as defined for aggregate variant association tests.

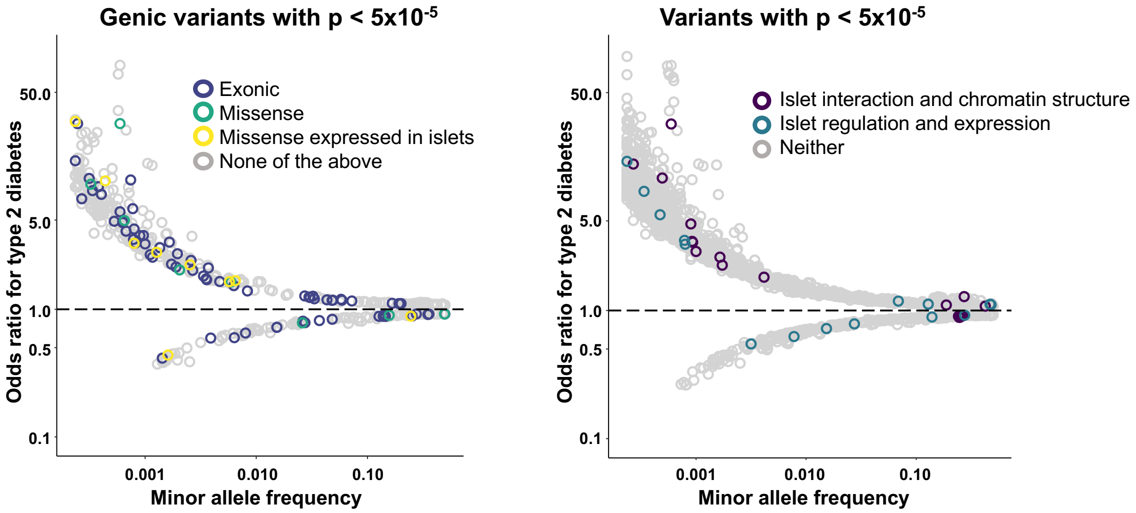

**Supplementary Figure 10. Manhattan, quantile-quantile, and P value-effect size scatter plots for single variant association analyses with the T2D+ outcome.** For each ancestry/population group, meta-analyses, or pooled analyses, a Manhattan style plot was generated using results from both BMI adjusted (purple) and BMI unadjusted (green) models. Quantile-quantile plots are in the center with genomic inflation for each model at the top left of each plot. Furthest to the right, scatter plots comparing BMI adjusted and BMI unadjusted P values are shown. Points in each plot are colored by the model (BMI adj. or BMI unadj.) in which the smaller P value was observed. The size of each point is proportional to the effect size estimate in the more significant model. Horizontal and vertical dashed lines are placed at P value = $5x10^{-8}$. For some analyses, see the inset for the full distribution of P values and effect sizes as the plotting ranges have been limited to P value < $1x10^{-15}$.

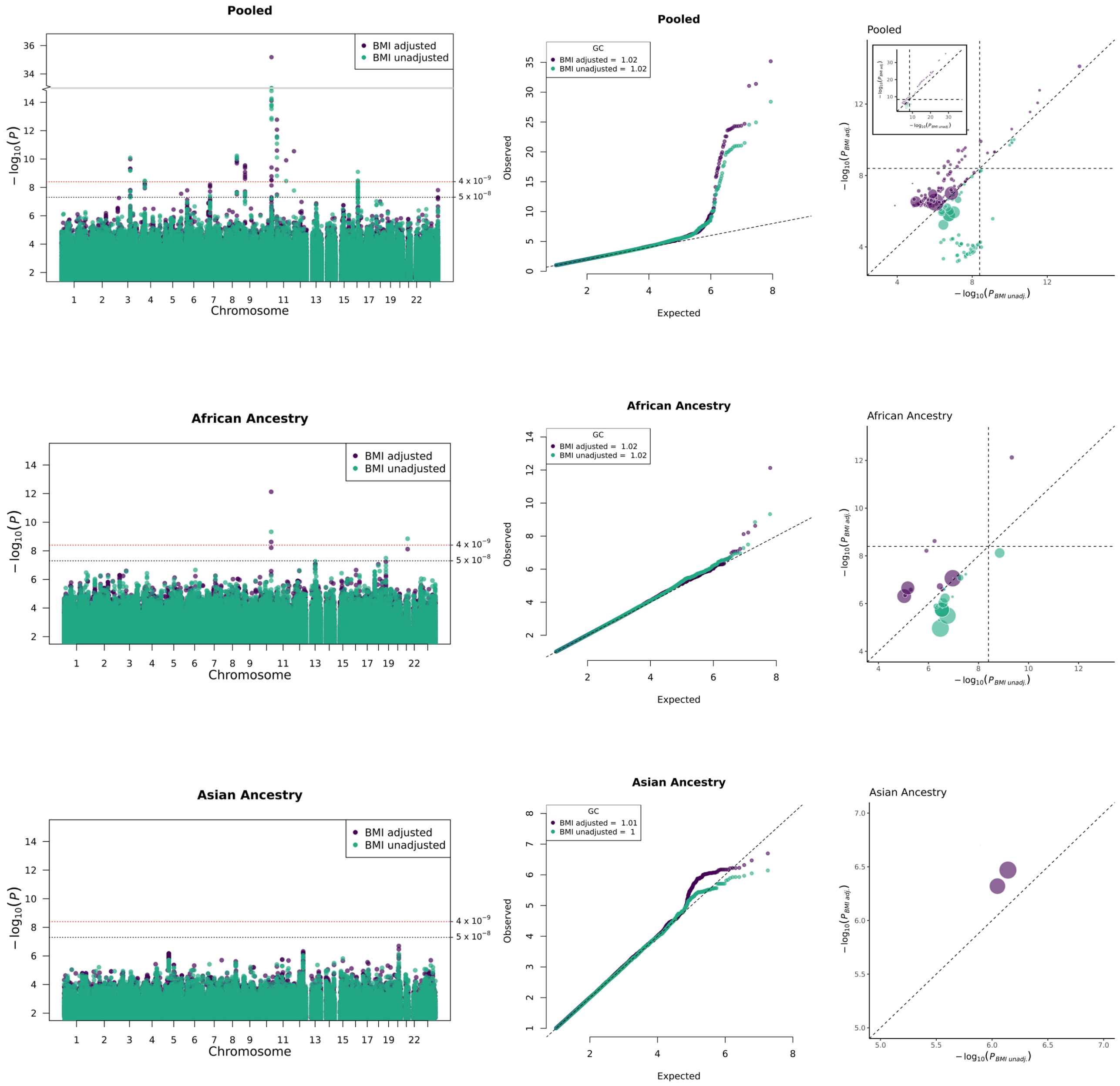
Pooled
BMI adjusted
BMI unadjusted
4 x 10^-9
5 x 10^-8
− log10(P)
Chromosome
GC
BMI adjusted = 1.02
BMI unadjusted = 1.02
Observed
Expected
− log10(P BMI adj.)
− log10(P BMI unadj.)
African Ancestry
GC
BMI adjusted = 1.02
BMI unadjusted = 1.02
Asian Ancestry
GC
BMI adjusted = 1.01
BMI unadjusted = 1

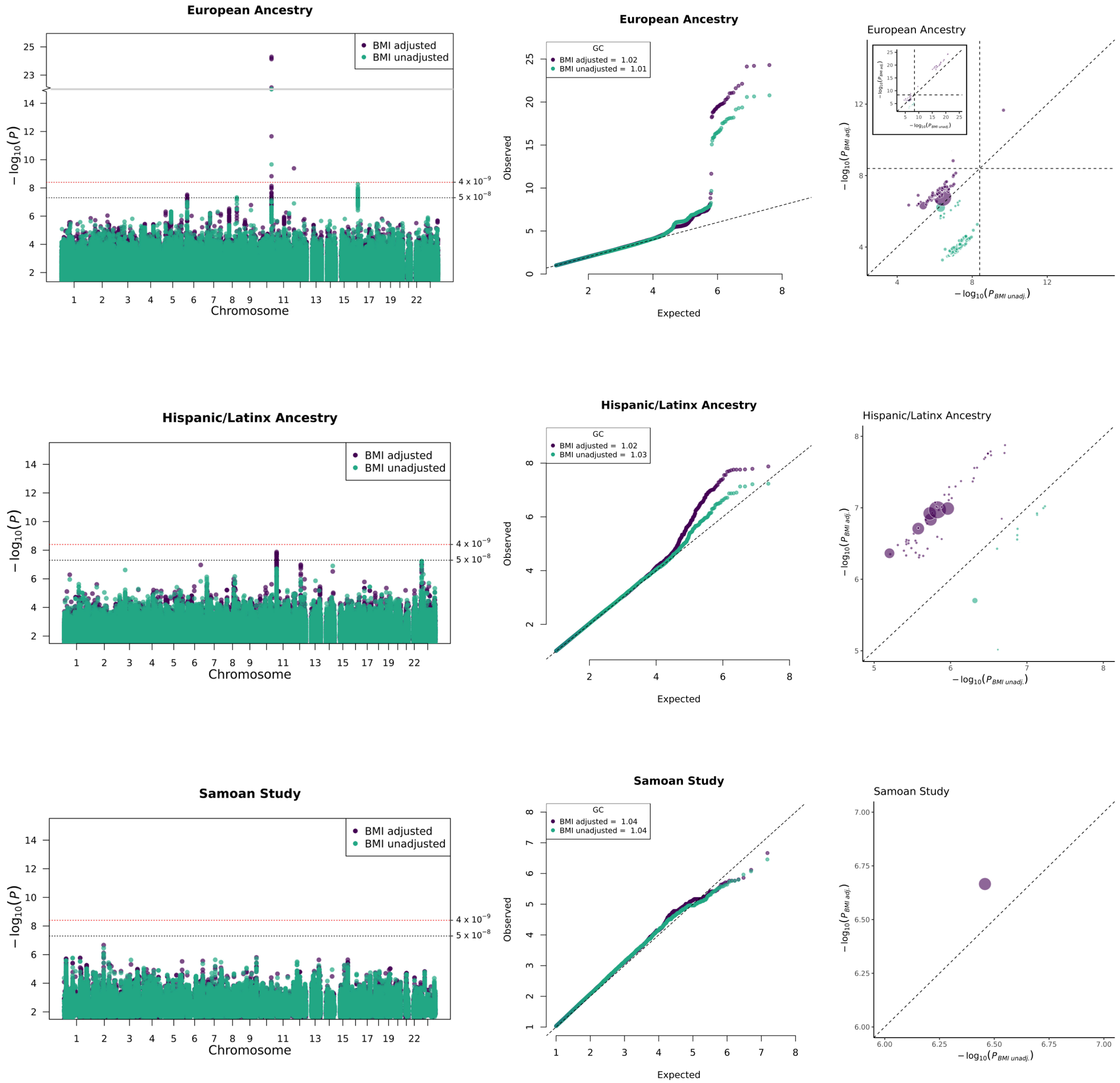
European Ancestry
BMI adjusted
BMI unadjusted
4 x 10$^{-9}$
5 x 10$^{-8}$
$-\log_{10}(P)$
Chromosome
GC
BMI adjusted = 1.02
BMI unadjusted = 1.01
Observed
Expected
$-\log_{10}(P_{BMI\ adj.})$
$-\log_{10}(P_{BMI\ unadj.})$
Hispanic/Latinx Ancestry
BMI adjusted
BMI unadjusted
4 x 10$^{-9}$
5 x 10$^{-8}$
$-\log_{10}(P)$
Chromosome
GC
BMI adjusted = 1.02
BMI unadjusted = 1.03
Observed
Expected
$-\log_{10}(P_{BMI\ adj.})$
$-\log_{10}(P_{BMI\ unadj.})$
Samoan Study
BMI adjusted
BMI unadjusted
4 x 10$^{-9}$
5 x 10$^{-8}$
$-\log_{10}(P)$
Chromosome
GC
BMI adjusted = 1.04
BMI unadjusted = 1.04
Observed
Expected
$-\log_{10}(P_{BMI\ adj.})$
$-\log_{10}(P_{BMI\ unadj.})$

**Supplementary Figure 11. Enrichment scores from LD Score Regression for type 2 diabetes and glycemic trait associations from the DIAGRAM, MEDIA, MAGIC and AAGILE consortia.** LD Score regression was performed with summary statistics from each GWAS with 53 genomic annotations from the LD Score Regression authors and 68 tissue-specific annotations based on GenoSkyline Plus scores. We present annotations with P values less than the Bonferroni corrected level of significance in at least one GWAS data set.

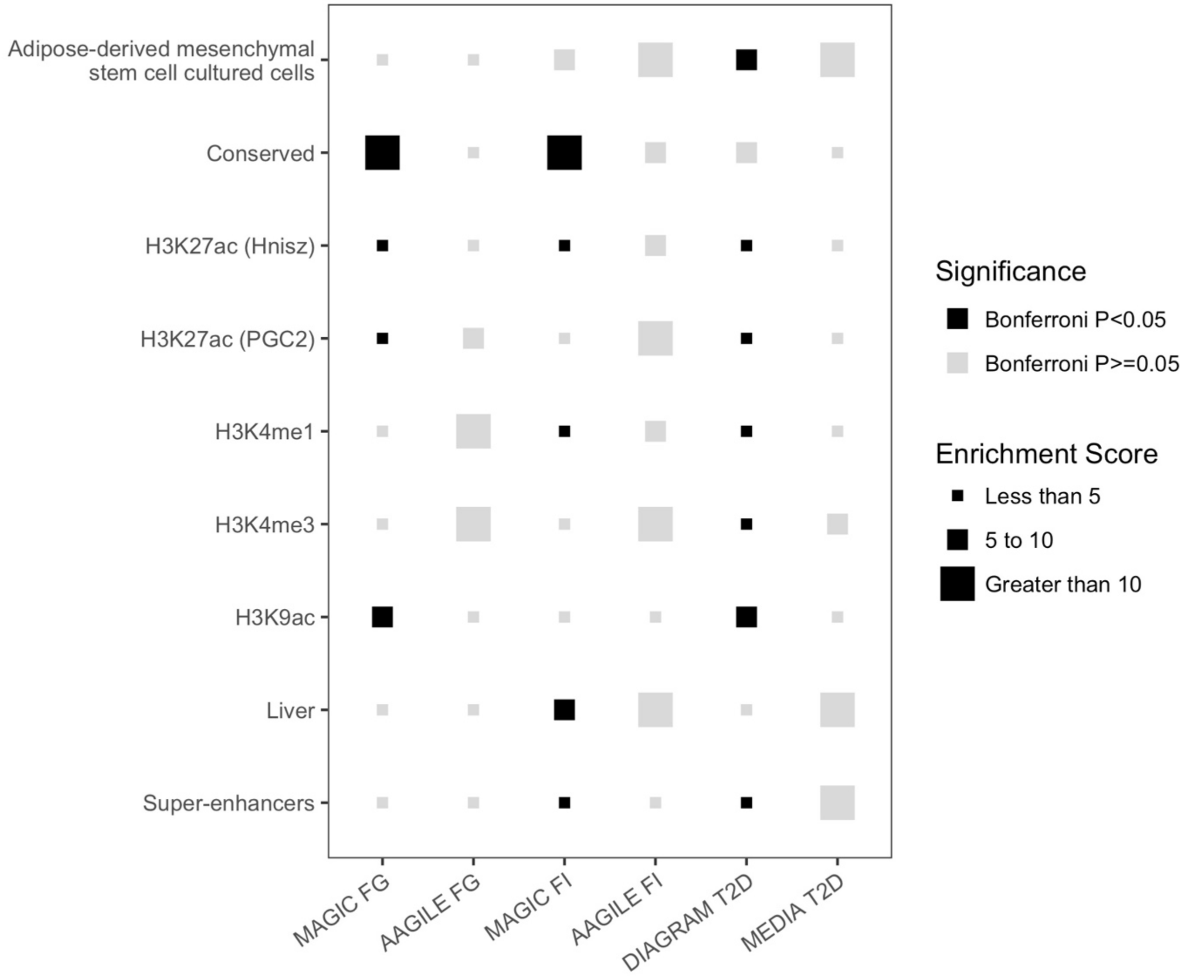
